# Risk Factor–Based Metabolomic Profiling Reveals Plasma Biomarkers of Hepatobiliary Cancer

**DOI:** 10.64898/2026.03.09.26347912

**Authors:** Felix Boekstegers, Vivian Viallon, Marie Breeur, Cosmin Voican, Gabriel Perlemutter, Chrysovalantou Chatziioannou, Pekka Keski-Rahkonen, Dominique Scherer, Mazda Jenab, Justo Lorenzo Bermejo

**Author notes:** **Correspondence:** Address correspondence to: Felix Boekstegers, PhD, Biostatistics and Data Integration Team, International Agency for Research on Cancer, 25 Avenue Tony Garnier, 69366 Lyon cedex 07, France. these authors contributed equally to this work (Mazda Jenab, Justo Lorenzo Bermejo).

## Abstract

**Background and Aims:** Highly aggressive hepatobiliary tumours include gallbladder cancer (GBC), hepatocellular carcinoma (HCC), intrahepatic and extrahepatic cholangiocarcinoma (iCCA, eCCA) and ampulla of Vater cancer (AoV). We aimed to identify plasma biomarkers for the early diagnosis of hepatobiliary cancer by leveraging the metabolomic signatures of established clinical risk factors.

**Method:** Based on 273,190 participants from the UK Biobank, we (1) identified metabolites associated with gallstone-related conditions (e.g. cholecystitis), primary sclerosing cholangitis (PSC) and metabolic liver diseases (e.g. cirrhosis), and (2) evaluated the relationship between the identified metabolites and the risk of GBC, HCC, iCCA, eCCA and AoV. Findings were validated in an independent group of 227,809 participants from the UK Biobank. We also derived metabolomic scores summarizing the three risk-factor signatures and evaluated their ability to stratify cancer risk.

**Results:** We identified 27 metabolites associated with gallstone-related conditions, 11 with PSC, and 34 with metabolic liver diseases, some of which showed associations with inconsistent directions across risk factors, suggesting distinct pathogenic processes. Several metabolites were associated with cancer risk in both the discovery and validation datasets, independently of established risk factors, predominantly for HCC (16 signals) and for iCCA (4), with one for GBC and none for eCCA and AoV. Metabolomic scores clearly distinguished individuals at high risk for HCC and iCCA.

**Conclusion:** The preselection of plasma metabolites associated with established risk factors facilitated the subsequent identification and validation of biomarkers for early cancer detection. The identified metabolites suggest specific pathogenic pathways for each type of hepatobiliary cancer. Wider replication is urgently needed to advance toward clinical implementation.

**What you need to know:** *BACKGROUND AND CONTEXT:* Clinical risk factors for hepatobiliary cancers often progress silently, making early identification of high-risk individuals difficult and highlighting the need for biological markers detectable before clinical diagnosis.

*NEW FINDINGS:* Risk-factor–based serum metabolomic profiling identified circulating metabolites that predict specific hepatobiliary cancers years before diagnosis, with strongest and most consistent signals for hepatocellular and intrahepatic cholangiocarcinoma.

*LIMITATIONS:* Clinical risk factors were assumed to be frequently underdiagnosed in UK Biobank, and event numbers were relatively small for some cancers, which may have reduced power and attenuated associations for less common endpoints.

*CLINICAL RESEARCH RELEVANCE:* This study shows that serum metabolic profiles can identify individuals at increased risk for hepatobiliary cancers long before symptoms appear, particularly for hepatocellular and intrahepatic cholangiocarcinoma. These findings support the development of precision risk-stratification strategies that may ultimately enable earlier surveillance.

*BASIC RESEARCH RELEVANCE:* By first identifying metabolites linked to specific liver and biliary clinical conditions, the study clarifies which metabolites are indirectly associated with hepatobiliary cancers through known disease pathways. Testing these metabolites again while adjusting for diagnoses of those conditions then reveals which ones also show direct, pathway-independent associations with individual hepatobiliary cancers, providing clearer insight into cancer-specific metabolic mechanisms.

## INTRODUCTION

Hepatobiliary cancers represent a group of anatomically adjacent but biologically heterogeneous malignancies that includes hepatocellular carcinoma (HCC), intrahepatic and extrahepatic cholangiocarcinoma (iCCA and eCCA), gallbladder cancer (GBC), and Ampulla of Vater cancer (AoV). They are among the most aggressive gastrointestinal cancers and are typically diagnosed at advanced stages, when curative treatment options are limited and prognosis is poor.^1^

Hepatobiliary cancers share key metabolic and inflammatory risk pathways as well as clinical risk factors: HCC accounts for the largest burden and typically arises from chronic liver diseases (e.g. cirrhosis) related to chronic hepatitis B or C infection, moderate to heavy alcohol consumption, or the increasingly prevalent metabolic dysfunction–associated steatotic liver disease (MASLD), which may progress to steatohepatitis (MASH) and then to cirrhosis.^2–4^ Major conditions predisposing to iCCA include cirrhosis of viral, alcoholic, or metabolic origin, and primary sclerosing cholangitis (PSC), which is also a main factor for eCCA risk.^5, 6^ For GBC, gallstone disease is the dominant causal risk factor, and PSC has also been associated with increased risk.^7, 8^ AoV is rarer and less well known, but the literature suggests that gallstone disease and PSC may contribute to its aetiology.^9, 10^ All of these clinical precursor conditions produce sustained inflammatory, oxidative, fibrotic or cholestatic stress and may progress silently of non-specifically to hepatobiliary cancer, and most people affected by them are not identified in routine medical care.^11–13^ As a result, even where structured hepatobiliary cancer screening or prevention programs exists, only a small fraction of high-risk individuals are recognized early enough for surveillance or intervention.^14^

Circulating metabolites provide an opportunity for early identification of individuals at high risk for hepatobiliary cancer. Disruptions in lipid, glucose, amino-acid, bile-acid, and inflammatory pathways are hallmarks of chronic liver and biliary conditions such as MASLD, MASH, cirrhosis, PSC and gallstones.^15–18^ Since these pathways also play a central role in hepatobiliary carcinogenesis, circulating metabolites may capture biologically relevant signals long before cancer becomes clinically detectable. Yet prior metabolomic studies of hepatobiliary cancers have been limited by small case numbers, strong correlations among metabolites, and analytical approaches that either prioritized a narrow set of molecules or aggregated distinct types of hepatobiliary cancer, limiting the biological specificity and reproducibility of results.^19–22^

These considerations motivated the present two-phase study. In the first phase, we identified plasma metabolites associated with major risk factors for hepatobiliary cancer (MASLD, MASH, cirrhosis, PSC, gallstone-related conditions). In phase two, we assessed whether metabolites associated with risk factors were also related to the development of specific types of hepatobiliary cancer. The adjustment for risk factors in the in the statistical analyses of the second phase of the study was essential for identifying those metabolites that influence the risk of hepatobiliary cancer independently of established risk factors, which may reflect downstream biological processes, subclinical disease progression, or early carcinogenic changes that are not currently considered in routine clinical diagnostics. Collectively, our results indicate that metabolic alterations related to risk factors and hepatobiliary carcinogenesis are detectable years before clinical symptoms and may inform early diagnosis beyond clinical records.

## MATERIALS AND METHODS

Using targeted plasma metabolomic data from the UK Biobank, we performed a two-phase biomarker discovery, followed by independent validation (**Figure 1**).^23^ In the first phase, we identified metabolites associated with key clinical risk factors for hepatobiliary cancer, including gallstones, cholecystitis, cholecystectomy, PSC, MASLD, MASH and cirrhosis. In phase two, we analysed the association between metabolites related to risk factors and the development of hepatobiliary cancers types (GBC, HCC, eCCA, iCCA, AoV), considering the risk factors as potential confounders and adjusting statistical analyses accordingly. The associations identified in phase two were subsequently evaluated in a separate cohort of participants from the UK Biobank.

**Figure 1.**
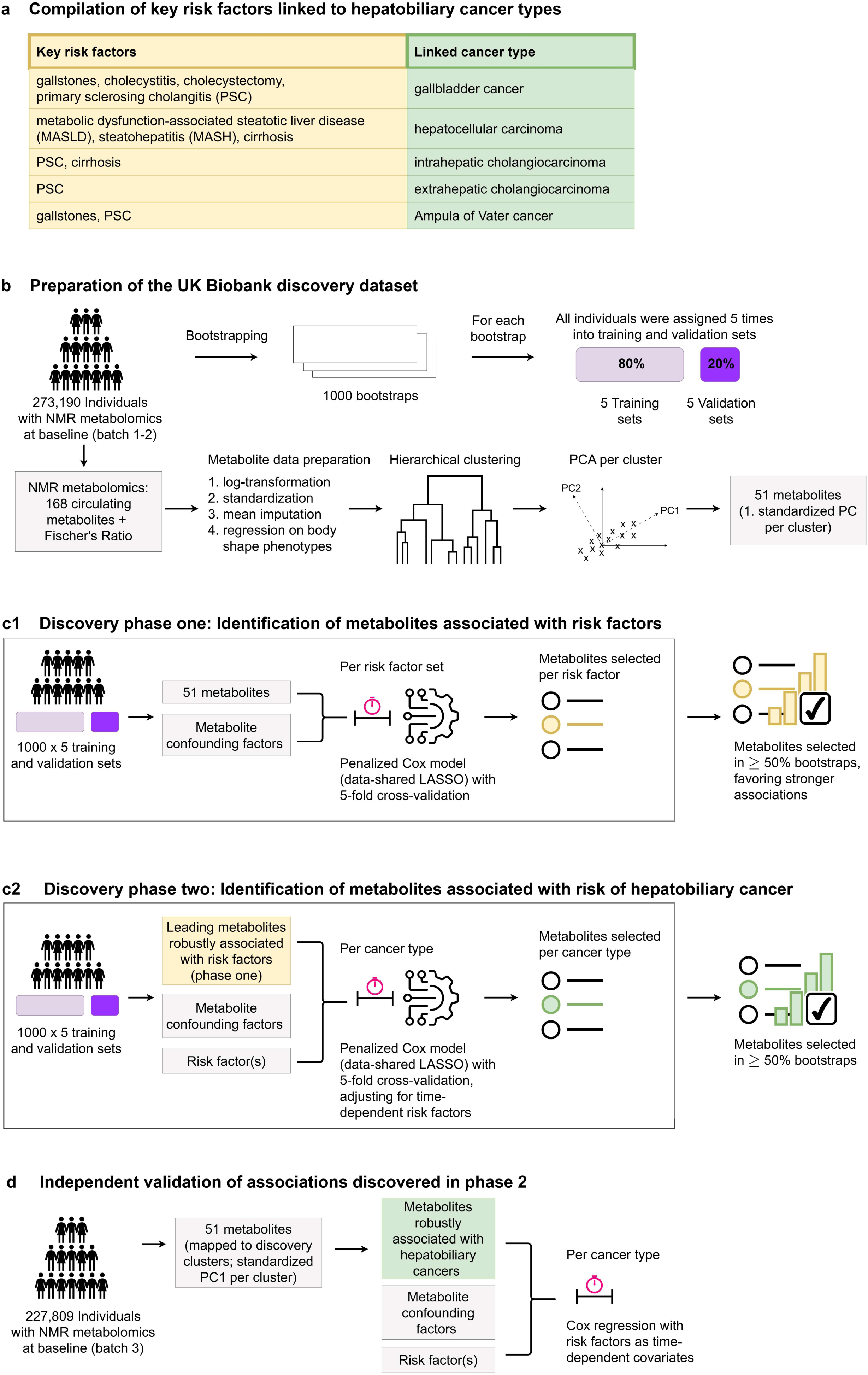
Study overview. **a,** Key clinical risk factors were compiled for each hepatobiliary cancer type. **b**, Targeted metabolomics data from UK Biobank were preprocessed for the discovery analysis. The eligible population (NMR blood metabolomics in batch1-2 and valid consent) was bootstrapped 1000 times and split into training and validation sets using 5-fold nested cross validation. Metabolite preprocessing included technical correction, log-transformation, standardization, adiposity-adjustment, and hierarchical clustering, yielding 51 standardized cluster representatives or isolated metabolites, hereafter referred to as *metabolites*. **c1,** In phase one, multivariate Cox models with a data-shared lasso penalty were fitted in each bootstrap to identify metabolites associated with three sets of clinical risk factors: (1) gallstone-related conditions (gallstones, cholcystitis, cholecystectomy), (2) PSC, and (3) metabolic liver diseases (MASLD, MASH, cirrhosis). Metabolites selected in ≥ 50% of bootstraps were considered robust associations, and those with the highest HRs were prioritized for phase two. **c2,** In phase two, Cox models were fitted to assess the association between risk factor-related metabolites and the risk of hepatobiliary cancer types, with robust associations again defined as those metabolites selected in ≥50% of bootstraps. **d,** In the independent validation, the associations between plasma metabolite levels and hepatobiliary cancer risk were re-evaluated in UK Biobank batch 3 using the same Cox model, with clinical risk factors included as time-dependent covariates. Discovery and validation HR were compared for consistency in effect direction and magnitude. AoV, Ampulla of Vater cancer; eCCA, extrahepatic cholangiocarcinoma; GBC, gallbladder cancer; iCCA, intrahepatic cholangiocarcinoma; HCC, hepatocellular carcinoma; MASH, Metabolic dysfunction-associated steatohepatitis; MASLD, Metabolic dysfunction-associated steatotic liver disease; PSC, primary sclerosing cholangitis.

### Study population and endpoint definition

Discovery and validation analyses were conducted within the UK Biobank cohort, which includes approximately 500,000 participants enrolled between 2006 and 2010 across the United Kingdom, with ongoing follow-up.^24^ EDTA plasma samples collected at UK Biobank’s baseline assessment were utilised for targeted nuclear magnetic resonance (NMR, Nightingale Health) metabolomics profiling (**Suppl. Table S1**) and released in three separate batches: **batches 1-2** (∼ 274,000 participants) served for discovery in this study, and **batch 3** (∼ 227,000 participants) served for validation. Ethics approval was granted by the North West Multi-centre Research Ethics Committee (Ref: 11/NW/0382). All participants provided written informed consent, and those who withdrew consent were not included in the analyses.

Seven established risk factors were considered for the five types of hepatobiliary cancer investigated (**Figure 1a; Suppl. Table S2** for the coding definitions). Gallstone-related conditions were not considered primary risk factors for HCC, iCCA or eCCA, because their associations were weaker compared with PSC, MASLD, MASH or cirrhosis.^25, 26^ The date of onset of clinical risk factors and hepatobiliary cancer types was defined as the earliest available data of diagnosis.

### Data preprocessing

We evaluated the 168 circulating biomarkers measured in plasma samples from the UK Biobank using the NMR assay and additionally calculated Fischer’s ratio (the molar ratio of branched-chain amino acids to aromatic amino acids, **Suppl Table S3**), a clinical indicator of liver damage and a potential prognostic marker for HCC.^27, 28^ Metabolitomic data were pre-processed using an established pipeline (**Figure 1b**): Technical variation was removed using the *ukbnmr* R package.^29^ Values were log-transformed and standardized, and missing values were replaced with the sample mean. To account for potential confounding by adiposity and body fat distribution, linear models were fitted for each metabolite with body shape phenotypes as covariates, using the residuals for further analysis.^30, 31^

Highly correlated metabolite residuals were grouped into clusters using ascendant hierarchical clustering (*ClustOfVar* R package).^32^ For each identified cluster, the first principal component (PC1) served as a surrogate metabolite, retaining the smallest number of clusters that cumulatively explained at least 80% of the metabolomic data variance. To ease interpretation, metabolite clusters were labelled according to their most recognized constituent metabolite, and cluster labels were used solely as descriptive abbreviations. Hereinafter, we refer to both single metabolites and cluster surrogate metabolites simply as ***metabolites***.

Metabolomic validation data (batch 3) were pre-processed using the same pipeline as for the discovery data. To ensure comparability and avoid overfitting, the standardization parameters, adiposity-adjustment coefficients and PC loads calculated in the discovery dataset were also used for validation. After adiposity adjustment, metabolites were mapped to the discovery-defined cluster structure by projecting batch 3 residuals onto the discovery PC1 loadings. These projections were used for the validation analyses.

### Survival modelling approach used for all analyses

The associations between plasma metabolite levels (per standard deviation) and clinical risk factors or the risk of hepatobiliary cancer types was assessed using multivariable Cox proportional hazard models with age as the underlying time scale. Participant follow-up started at the age of blood sampling and ended at diagnosis of the investigated event, or lost-to-follow up, death or the end of the follow-up period (right censoring). Individuals diagnosed with any type of cancer prior to blood sampling were excluded (left truncation). The hazard models included age at blood sampling, sex, fasting time, and, for women, menopausal status and use of exogenous hormones. Analyses on the association with the risk of hepatobiliary cancer types were further adjusted for clinical risk factors as time-dependent variables, and sensitivity analyses considered additional covariates such as alcohol consumption, and stratification by sex.

### Discovery phase one

In phase one, we investigated the association between the metabolite levels and the occurrence of three categories of established hepatobiliary cancer risk factors: gallstone-related clinical conditions (detection of gallstones, cholecystitis, cholecystectomy), PSC, and metabolic liver diseases (MASLD, MASH, cirrhosis), the latter category representing a progressive continuum from metabolic steatosis to advanced fibrosis and cirrhosis. Metabolites showing Benjamini–Hochberg–corrected association p-values < 0.05 with at least one risk factor were retained for further analyses. For each category of established clinical risk factors, the metabolites retained were analysed by data-shared lasso regularized Cox regression with five-fold cross-validation, defining adjustment factors as unpenalized variables (**Figure 1c**). The data-shared lasso extends the standard lasso by jointly modelling multiple related endpoints, leveraging shared metabolic patterns across risk factors while allowing associations specific to individual endpoints.^33, 34^ Compared to standard methods, this approach has demonstrated strong performance in identifying features with consistent non-null effects across correlated conditions.^35^ For PSC, models reduced to a standard lasso. Robustness was assessed using 1,000 bootstrap samples and metabolites selected in ≥ 50% of iterations were considered robustly associated with the corresponding endpoint. Following the rationale of the lasso ordinary least squares approach, and recognising that shrinkage bias may be more pronounced given the very different case numbers across the gallstone related clinical conditions and metabolic liver diseases, effect estimates for selected metabolites were obtained from unpenalized Cox models in each bootstrap to quantify their strength and assess potential heterogeneity beyond what was captured by the penalized models.^36^

### Discovery phase two

Metabolites identified in phase one were tested for association with the occurrence of hepatobiliary cancers (GBC, HCC, iCCA, eCCA, AoV) using lasso-penalized Cox models (**Figure 1d**). To avoid overfitting in cancer types with few cases, we limited the number of candidate metabolites to roughly one per ten cases, in line with recommendations for variable selection processes.^37^ Candidate metabolites from phase-one analyses were prioritised by jointly considering association strength and selection frequency: in each bootstrap sample, non-selected metabolites were assigned a hazard ratio (HR) = 1, and metabolites were then ranked according to the absolute median HR across all bootstraps.

Because metabolites were preselected based on their association with clinical risk factors, some associations with risk of hepatobiliary cancer types were probably due to the association with the risk factors. To determine which metabolites showed associations with cancer risk independent of clinical risk factors, we included the relevant risk factors as time-dependent covariates in the hazard models (i.e., covariate values were updated at the age when a diagnosis was first recorded, so the follow-up before and after diagnosis was correctly allocated).^38, 39^ Robustness of metabolite–cancer associations was assessed using 1,000 bootstrap samples, with metabolites selected in ≥50% of iterations considered robust. Effect estimates for selected metabolites were obtained from unpenalized Cox models in each bootstrap.

We conducted several sensitivity analyses on association with the risk of hepatobiliary cancer types. To address possible reverse causality, the first two and seven years of follow-up were excluded. To assess residual confounding, models were further adjusted for body mass index (BMI), educational level, pack years of adult smoking and alcohol consumption. Finally, given known sex differences in GBC and HCC incidence, sex-stratified Cox models were also fitted.^40^

### Independent validation

Metabolite–cancer associations identified in phase two were re-evaluated in an independent dataset (UK Biobank batch 3). We fitted multivariable Cox models identical to those fitted in the discovery phase, including the same covariates (age, sex, fasting time, menopause status and hormone use in women) and relevant clinical risk factors as time-dependent covariates. The robustness of the associations was assessed by comparing the magnitude and direction of the discovery and validation HRs.

### Evaluation of the predictive performance of metabolomic risk scores

We evaluated the predictive performance of risk-factor-based metabolomic risk scores in UK Biobank batch 3. First, metabolomic scores were calculated from the standardized HR estimates from the discovery dataset, which were subsequently dichotomized at the top decile, defining high-versus lower-cancer-risk groups (top 10% vs. bottom 90%). We then investigated the association between the dichotomized metabolomic risk scores and the risk of each type of hepatobiliary cancer using multivariable Cox regression. For HCC and iCCA, we also benchmarked the predictive performance of a publicly available, externally validated cirrhosis-related polygenic risk score (PGS), considering three mutually exclusive cancer-risk groups:

1. individuals with top 10% risk according to both the PGS and the metabolomic risk score,
2. individuals with top 10% risk according to the PGS only (metabolomic risk score in the bottom 90%), and
3. individuals with PGS values in the bottom 90%.^41^

For each type of hepatobiliary cancer, HRs were estimated for the risk groups and derived absolute age-specific risks using a competing-risks framework consistent with the iCARE approach.^42^ Our method combined the cancer-specific HRs with population age-specific cancer incidence and all-cause mortality rates (Office for National Statistics, 2008), and integrated these hazards over age to obtain the cumulative absolute risk.^43^

Because many of our investigated clinical risk factors were likely underdiagnosed in UK Biobank, we repeated the analyses among participants with diagnosed risk factors. This allowed us to assess whether the risk factor-based metabolomic score captures cancer risk primarily through undiagnosed risk-factor pathways or whether it reflects cancer risk beyond the presence of the clinical diagnoses.

## RESULTS

We investigated 168 metabolites and the Fischer’s ratio measured in plasma samples from 273,190 (discovery) and 227,809 (validation) participants in the UK Biobank, with comprehensive clinical data (**Table 1**). Statistical analysis focused on five types of hepatobiliary cancer (GBC, HCC, iCCA, eCCA, AoV), and seven established risk factors: gallstone-related factors (gallstones, cholecystitis and cholecystectomy), PSC, and metabolic liver diseases (MASLD, MASH and cirrhosis).

**Table 1.**
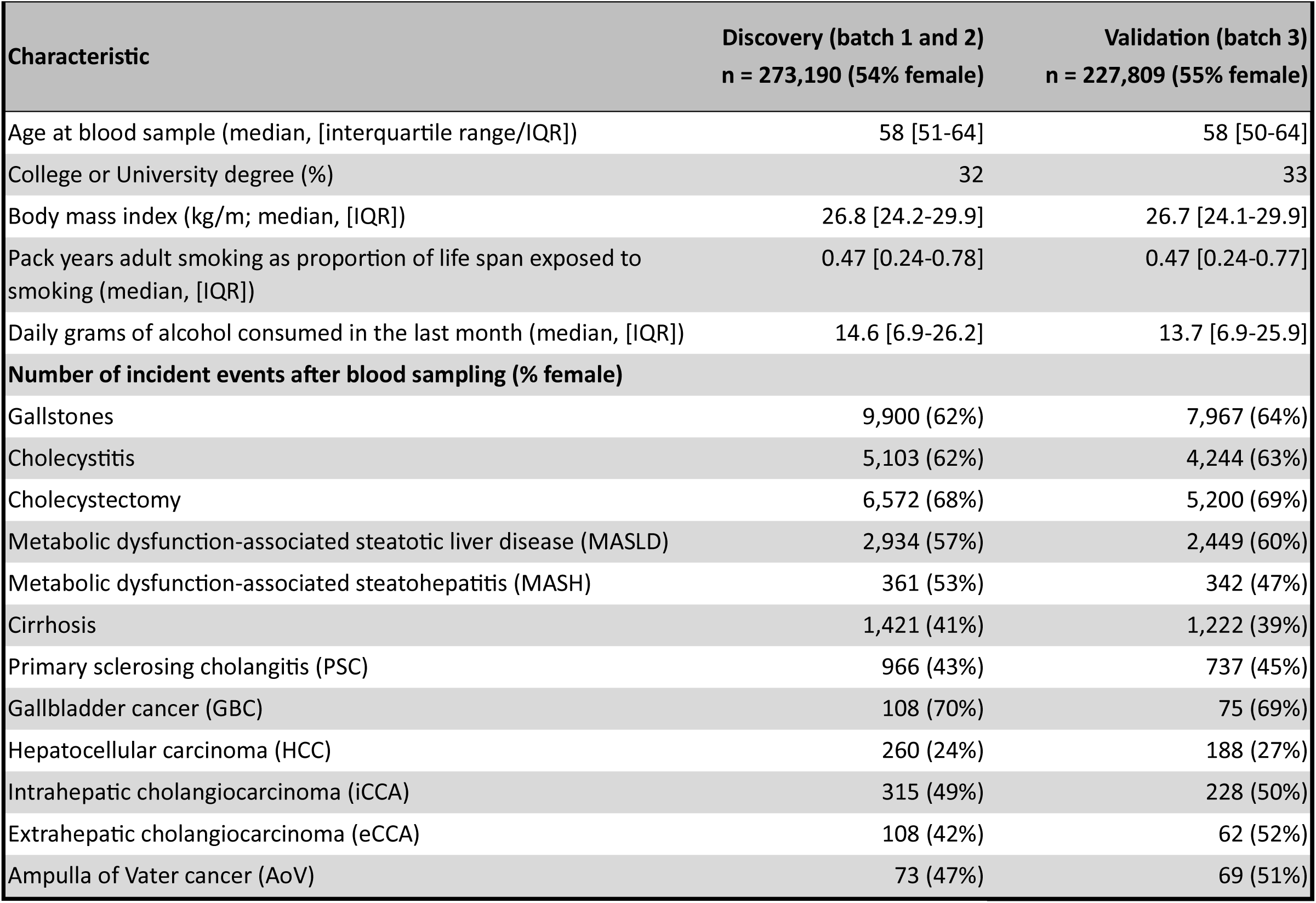
Main characteristics of the UK Biobank study population.

| Characteristic | Discovery (batch 1 and 2)<br>n = 273,190 (54% female) | Validation (batch 3)<br>n = 227,809 (55% female) |
| --- | --- | --- |
| Age at blood sample (median, [interquartile range/IQR]) | 58 [51-64] | 58 [50-64] |
| College or University degree (%) | 32 | 33 |
| Body mass index (kg/m; median, [IQR]) | 26.8 [24.2-29.9] | 26.7 [24.1-29.9] |
| Pack years adult smoking as proportion of life span exposed to smoking (median, [IQR]) | 0.47 [0.24-0.78] | 0.47 [0.24-0.77] |
| Daily grams of alcohol consumed in the last month (median, [IQR]) | 14.6 [6.9-26.2] | 13.7 [6.9-25.9] |
| **Number of incident events after blood sampling (% female)** |  |  |
| Gallstones | 9,900 (62%) | 7,967 (64%) |
| Cholecystitis | 5,103 (62%) | 4,244 (63%) |
| Cholecystectomy | 6,572 (68%) | 5,200 (69%) |
| Metabolic dysfunction-associated steatotic liver disease (MASLD) | 2,934 (57%) | 2,449 (60%) |
| Metabolic dysfunction-associated steatohepatitis (MASH) | 361 (53%) | 342 (47%) |
| Cirrhosis | 1,421 (41%) | 1,222 (39%) |
| Primary sclerosing cholangitis (PSC) | 966 (43%) | 737 (45%) |
| Gallbladder cancer (GBC) | 108 (70%) | 75 (69%) |
| Hepatocellular carcinoma (HCC) | 260 (24%) | 188 (27%) |
| Intrahepatic cholangiocarcinoma (iCCA) | 315 (49%) | 228 (50%) |
| Extrahepatic cholangiocarcinoma (eCCA) | 108 (42%) | 62 (52%) |
| Ampulla of Vater cancer (AoV) | 73 (47%) | 69 (51%) |

### Clustering of metabolites

Strong correlations were observed among most metabolites, particularly within fatty acids and lipoprotein subclasses, including High-Density Lipoprotein (HDL), Low-Density Lipoprotein (LDL), Very Low-Density Lipoprotein (VLDL) and Intermediate-Density Lipoprotein (IDL) (**Suppl. Figure S1**). Hierarchical clustering resulted in 144 metabolites grouped into 26 clusters, each cluster containing between 2 and 15 metabolites, while 25 metabolites remained isolated, primarily amino acids, glycolysis-related metabolites, and ketone bodies (**Suppl. Figure S2**). Clusters generally included metabolites from the same chemical classes, and correlations between metabolites in each cluster and the cluster representative were typically close to 1. On average, cluster representatives explained 98% of the within-cluster variance in plasma metabolite levels (range: 85–100%), and the combined set of 26+25 = 51 metabolites captured over 98% of the total variance. Nevertheless, residual correlations remained between certain cluster representatives (**Suppl. Figure S3**), underscoring the value of lasso regression combined with bootstrapping to identify robust associations.

### Discovery phase one: Metabolites associated with hepatobiliary cancer risk factors

In phase one, we identified 27 metabolites associated with gallstone-related conditions, 11 with PSC, and 34 with metabolic liver diseases (**Figure 2a, Suppl Tables S4-S13**). The strength of several metabolite associations (phosphatidylcholines cluster, XS-VLDL-P cluster, triglycerides in large HDL) increased along the sequence of MASLD to MASH and cirrhosis. Metabolites in the lipid-cluster, in particular sphingomyelins, phosphatidylcholines, and triglyceride-rich lipoproteins, were associated with hepatobiliary cancer risk factors, while omega-3 fatty acids and free cholesterol in small HDL showed a protective effect. It is noteworthy that some metabolites showed opposite associations across risk factors (e.g., rising plasma levels of XS-VLDL-P cluster metabolites were associated with a lower risk of gallstone-related conditions, but with an increased risk of metabolic liver diseases), suggesting distinct pathogenic processes.

**Figure 2.**
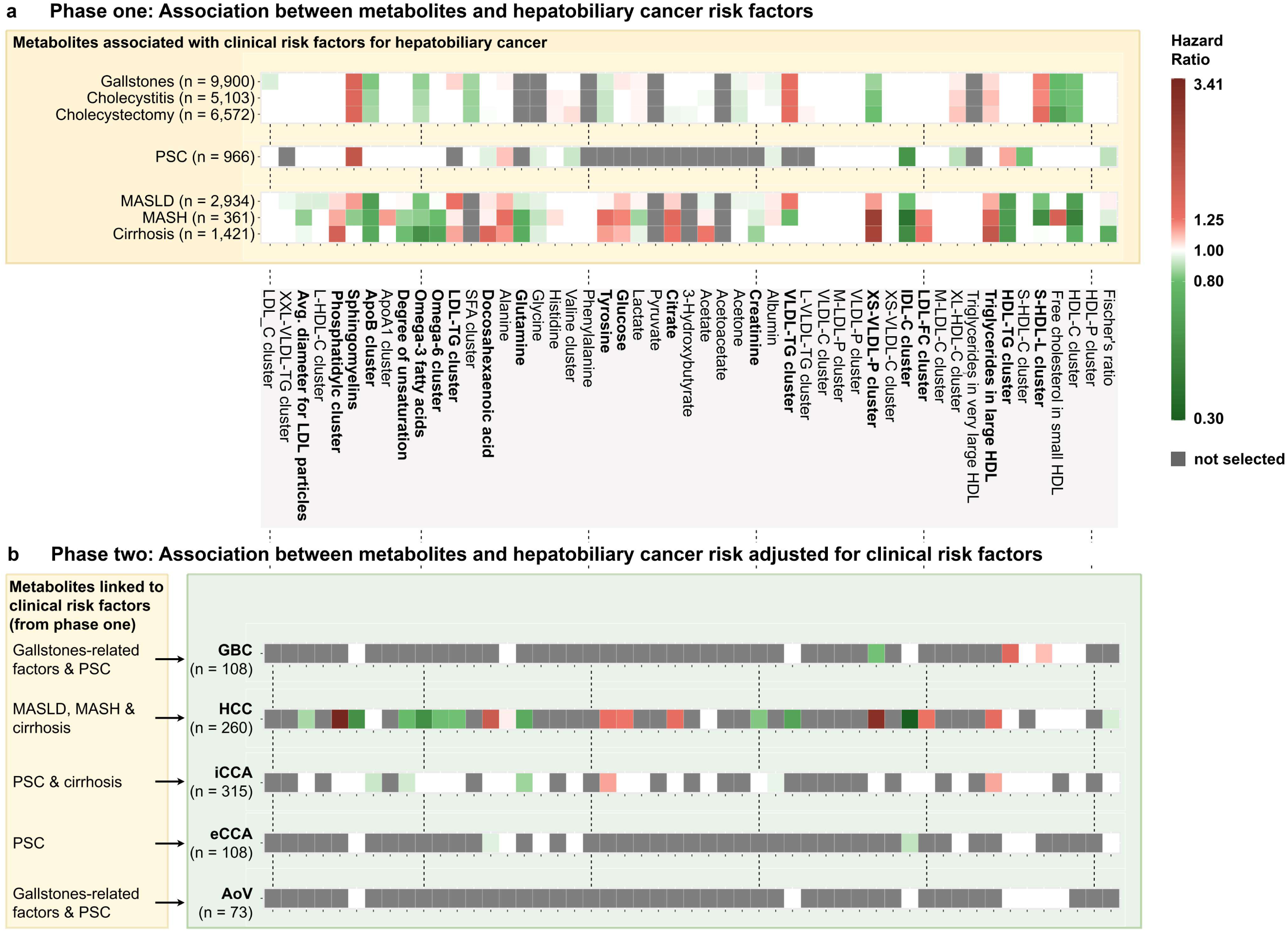
Associations between metabolites and the hepatobiliary cancer risk factors, and between metabolites and the risk of hepatobiliary cancer among 273,190 UK Biobank participants. **a,** Results from multivariate Cox regression analysis to identify metabolites associated with hepatobiliary cancer risk factors (phase one), evaluated by bootstraping and data-shared lasso penalty in three risk factor subsets: 1. gallstone-related clinical conditions (gallstone detection, cholecystitis, cholecystectomy), 2. PSC and 3. the sequence of metabolic liver disease (MASLD, MASH, cirrhosis). White entries show no identified associations (i.e. the corresponding metabolite was selected in less than 50% of bootstraps), while green and red entries show negative and positive associations, respectively (i.e. the metabolite was selected in more than 50% of bootstraps). The darker the colour, the higher the median of the bootstrap hazard ratio distribution. Grey entries show metabolites that did not show individual associations with hepatobiliary cancer risk factors. The x-axis represents the 51 metabolomic features investigated, names correspond to cluster surrogated or single metabolites. **b,** Results from univariate Cox regression analysis to identify metabolites associated with hepatobiliary cancer types (phase two), evaluated by bootstrapping and lasso penalty. For each cancer type a different list of metabolites was tested for association, depending on the results from phase one: only metabolites with the largest absolute values of the median of the bootstrap hazard ratio distribution were chosen, and the number of metabolites examined was limited to the number of cancer cases divided by ten. White, green and red entries and the x-axis have the same interpretation as for phase one. Grey entries correspond to metabolites that were not considered in phase two. AoV, Ampulla of Vater cancer; eCCA, extrahepatic cholangiocarcinoma; GBC, gallbladder cancer; iCCA, intrahepatic cholangiocarcinoma; HCC, hepatocellular carcinoma; MASH, Metabolic dysfunction-associated steatohepatitis; MASLD, Metabolic dysfunction-associated steatotic liver disease; PSC, primary sclerosing cholangitis.

### Discovery phase two: Metabolites associated with the risk of hepatobiliary cancers

In phase two, we investigated whether risk factor-related metabolites were independently associated with the risk of hepatobiliary cancers (**Figure 2b, Suppl Tables S14–S18)**. We observed the strongest and most numerous association signals for HCC, particularly with phosphatidylcholines, IDL-C, and the XS-VLDL-P cluster. For GBC, associations independent of the risk factors included XS-VLDL-P and HDL-TG clusters. For iCCA, tyrosine and triglycerides in large HDL showed positive associations, and plasma glutamine levels showed a protective effect. eCCA association signals were limited to IDL-C and docosahexaenoic acid, and no association was found for AoV. Overall, association directions were largely concordant with the risk-factor associations identified in phase one.

In sensitivity analyses, most associations between plasma metabolites and cancer risk remained after excluding the first two and seven years of follow-up to rule out reverse causality, and after accounting for additional potential confounders (**Suppl Figure S4**). Exceptions were the attenuation of the association between the Omega-6 cluster and tyrosine with HCC risk. In contrast, the association between the phosphatidylcholines cluster and the XS-VLDL-P cluster with HCC risk strengthened after excluding cases diagnosed in the first seven years after blood sampling, supporting their potential role as long-term risk indicators rather than markers of subclinical disease. Sex-stratified analyses (**Suppl Figure S5**) revealed considerable heterogeneity for some metabolites: the association between the phosphatidylcholines cluster and HCC risk was more than twice as strong in men, whereas the association between triglycerides in large HDL and HCC risk was notably stronger in women.

### Validation results

Most associations between plasma metabolites and hepatobiliary cancer risk showed directional consistency in the discovery and validation datasets, with slightly attenuated validation HR estimates (**Figure 3**). The strongest and most consistent replication signals were found for HCC: several lipids and amino acids displayed HRs in the same direction and of similar magnitude in the discovery and validation datasets. For iCCA, associations with unsaturation measures, the amino acid tyrosine, triglycerides in large HDL and the ApoB cluster were consistent with the discovery estimates, albeit with wider confidence intervals. For GBC, only the association with HDL-TG cluster metabolites remained directionally consistent. For eCCA, the associations found in the discovery dataset with the IDL-C cluster and Docosahecaenoic acid could not be validated.

**Figure 3.**
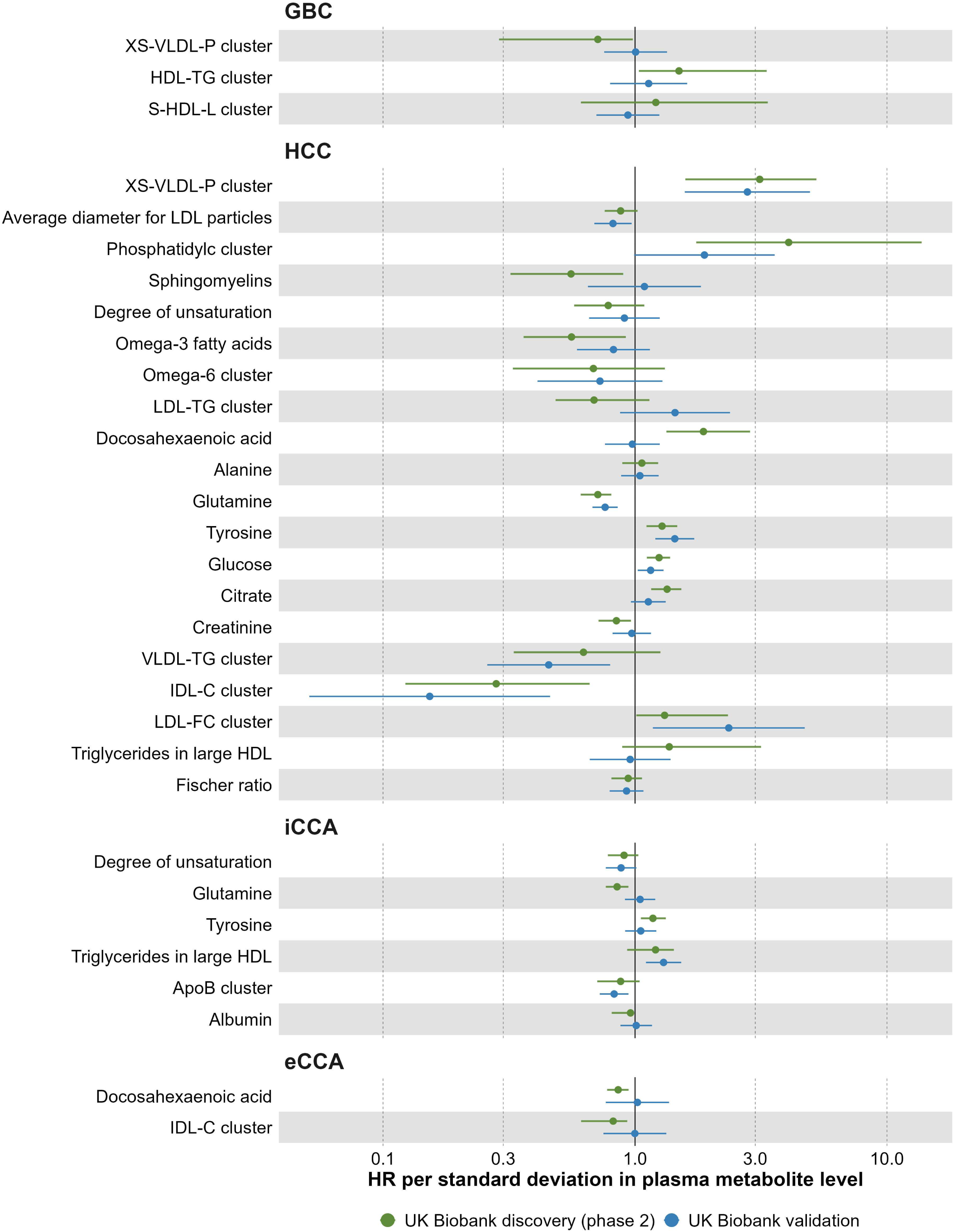
Associations between plasma metabolite levels and the risk of hepatobiliary cancer in the UK Biobank discovery and validation cohorts. Forest plots show hazard ratios for metabolites robustly associated with each hepatobiliary cancer type in discovery phase two (batches 1–2; green) and in the independent validation cohort (batch 3; blue). For discovery, point estimates and 95% percentile intervals correspond to the median and 2.5th/97.5th percentiles of hazard ratios across bootstrap samples in which the metabolite was selected. Validation hazard ratios and 95% confidence intervals per standard deviation are derived from unpenalized Cox models in batch 3. AoV, ampulla of Vater cancer; eCCA, extrahepatic cholangiocarcinoma; GBC, gallbladder cancer; iCCA, intrahepatic cholangiocarcinoma; HCC, hepatocellular carcinoma.

Dichotomization of UK Biobank participants (batch 3) based on metabolomic risk scores (top 10% vs. bottom 90%, **Figure 4**) clearly distinguished individuals at high risk for HCC. For HCC, individuals with a PGS and a metabolomic risk score in the top-deciles reached a cumulative risk of 1.5% at the age of 80. For iCCA, risk-curve separation was more modest than for HCC, but individuals in the joint top-decile of the PGS and metabolomic score still showed the highest cumulative risks, reaching approximately 0.6% by age 80, whereas those with a high PGS but lower metabolomic score did not exceed the reference PGS group. Risk differences for GBC, eCCA, and AoV did not reach statistical significance (overlapping 95% confidence bands). When restricting analyses to participants with diagnosed clinical risk factors (**Suppl. Figure S6**), the separation of risk curves remained broadly similar for HCC, eCCA, and AoV, and became more pronounced for GBC. For iCCA, however, the pattern differed: individuals with a top-decile PGS but metabolomic scores in the lower 90% showed slightly lower cumulative risk than the reference PGS group, while those with top-decile metabolomic scores continued to show the highest risk.

**Figure 4.**
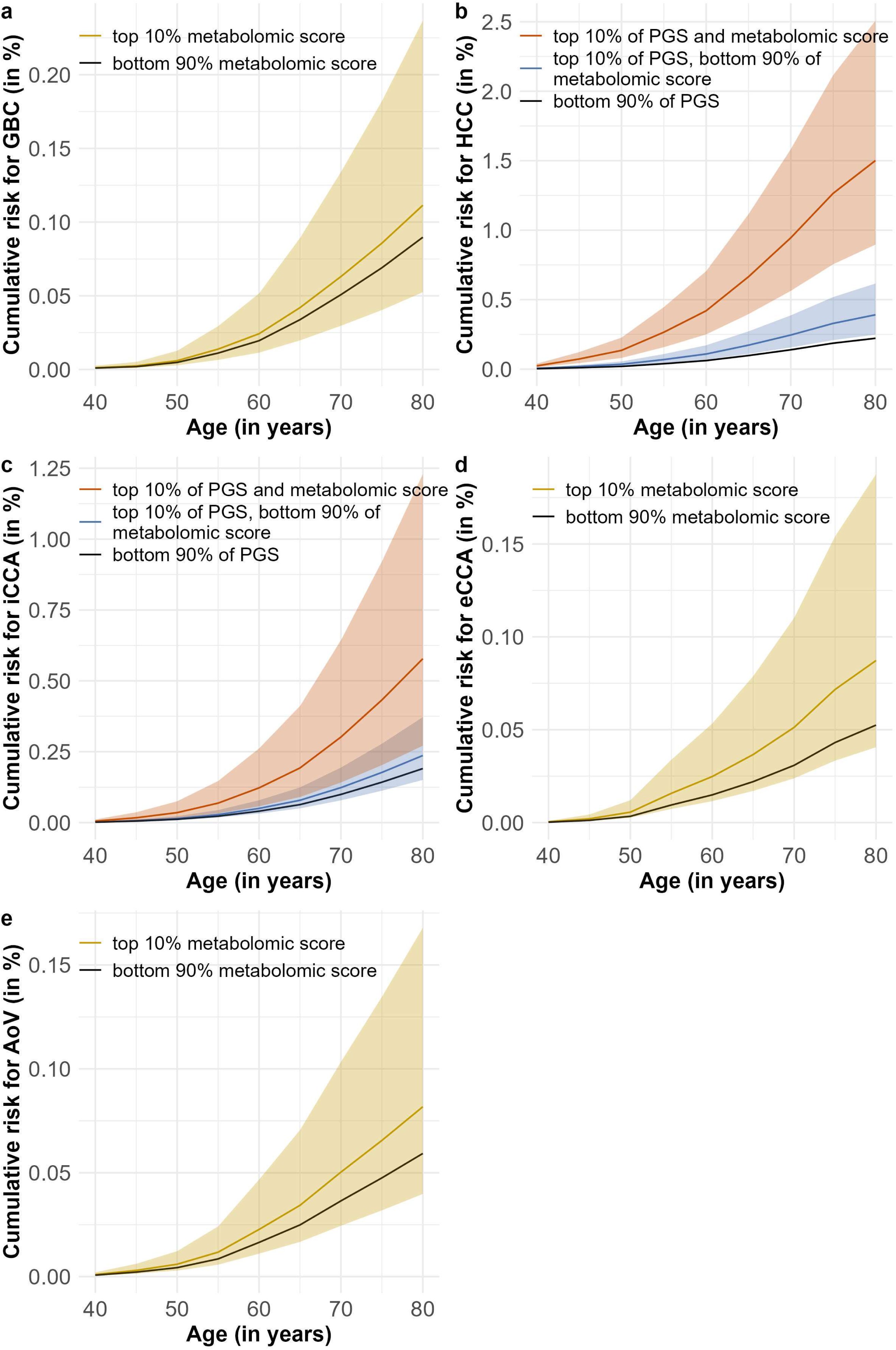
Absolute cumulative risk of hepatobiliary cancers according to metabolomic and genetic risk scores in the validation dataset (UK Biobank batch 3). Panels show age-specific absolute cumulative risk (in %) for five hepatobiliary cancer types based on metabolomic risk scores trained in the discovery study, and for hepatocellular carcinoma (HCC) and intrahepatic cholangiocarcinoma (iCCA) additionally based on polygenic risk scores (PGS). Curves were derived from multivariable Cox models applied in the independent validation cohort, with absolute risks estimated using a competing-risks framework incorporating UK population incidence and mortality rates. **a, d, e,** For gallbladder cancer (GBC), extrahepatic cholangiocarcinoma (eCCA), and ampulla of Vater cancer (AoV), individuals in the top 10% of the metabolomic score are compared with the remaining 90%. **b, c,** For HCC and iCCA, three groups are shown: (1) top-decile PGS and top-decile metabolomic risk score, (2) top-decile PGS and bottom 90% metabolomic risk score, (3) lower 90% of PGS (reference). Shaded areas represent 95% confidence intervals.

## DISCUSSION

In this study, our primary objective was to identify circulating metabolites that reflect biological processes linking established clinical risk factors (MASLD, MASH, cirrhosis, PSC, and gallstone-related conditions) to the subsequent development of specific types of hepatobiliary cancer. Because clinical precursor conditions often share metabolic signatures, and plasma metabolites build correlation clusters, a simple univariate screening would have favoured redundant features and risked conflating shared background biology with signals relevant to specific disease pathways. We therefore applied a risk-factor-based, two-phased metabolomic discovery based on penalized Cox regression, prioritizing those metabolites with non-redundant contributions and reducing false-positive findings driven by correlation structure alone. This design naturally led us to evaluate independent associations with the occurrence of incident cancer by adjusting for clinical risk factors as time-dependent variables, allowing us to distinguish metabolites that merely reflect known causal pathways from those that may provide additional etiologic information beyond routine clinical diagnoses.

In phase one, metabolite associations across the spectrum of metabolic liver disease showed clear severity-related patterns: for many metabolites, associations strengthened from MASLD to MASH and cirrhosis, while for others the most pronounced associations were observed among individuals who were later diagnosed with MASH. In contrast, metabolite associations with gallstone-related conditions were more uniform. Several metabolites overlapped across risk factor groups, occasionally with opposing directions of effect, indicating distinct metabolic processes that may differentially shape cancer susceptibility. In phase two, we assessed the association between several lipid-related clusters (e.g., phosphatidylcholines, IDL-C, XS-VLDL-P) and amino acids (e.g., glutamine, tyrosine) and cancer risk. Since the hazard models included diagnosis of risk factors as time-dependent variables, the identified associations can be interpreted as independent of recorded MASLD, MASH, cirrhosis, PSC, and gallstone-related disease, suggesting that these metabolites capture additional, biologically relevant processes beyond the investigated conditions. The persistence and, in some cases, strengthening of associations after exclusion of the first years of follow-up in sensitivity analyses argues against reverse causality and indicates that the detected metabolic changes arise well before clinical diagnosis, consistent with the long, stepwise evolution from chronic liver disease to HCC and from longstanding gallstone disease to GBC.^44, 45^ Sex-specific association signals (e.g., stronger association between phosphatidylcholines and HCC risk in men, higher HCC risk in women with elevated HDL-TG levels compared to men) confirm the relevance of sex-specific metabolism warranting targeted follow-up. Several identified metabolites, including lipid-related clusters and amino acids such as glutamine and tyrosine, have also been implicated in pathways related to liver disease and hepatobiliary carcinogenesis in prior prospective studies.^20, 21, 31^

Associations were robust to bootstrap resampling and additional multivariable adjustments. In the independent component of the UK Biobank cohort, directional replication was strongest for HCC, with supportive patterns for iCCA and imprecise HR estimates for GBC and eCCA, reflecting the smaller numbers of cases. When translated into absolute risk estimates, individuals in the top decile of the metabolomic score consistently showed the highest cumulative risk, with the largest separation observed for HCC, followed by iCCA. For HCC and iCCA, individuals who were in the top decile of both the metabolomic and polygenic risk scores exhibited the highest cumulative risk curves, indicating that the two scores provide complementary and largely additive risk stratification.

Earlier UK Biobank metabolomic studies on hepatobiliary cancer risk either focused on restricted analyte sets (i.e. fatty acids only), applied pan cancer screens before disease specific testing, or combined biliary cancers as a composite outcome.^20–22^ Our risk-factor informed, hepatobiliary cancer type specific approach complements this literature by prioritizing metabolites rooted in risk factor biology, maintaining cancer type specificity, and identifying additional associations while generally reproducing the direction and magnitude reported previously (details in **Suppl. Figures S7–S8**).

This study has several notable strengths. The prospective design and large sample size provided substantial statistical power to evaluate multiple hepatobiliary cancer types, and the availability of time-updated diagnoses for key clinical risk factors allowed us to more accurately account for disease trajectories preceding cancer onset. The risk factor-guided, two-phase analytic framework offered a biologically informed approach that enhanced specificity beyond conventional metabolite-wide analyses. In addition, the exploitation of an independent set of UK Biobank participants enabled validation of key findings.

We were only partially able to address several limitations. First, although the UK Biobank provided a uniquely large and deeply phenotyped resource, our reliance on routine ICD 10-based diagnoses to define clinical conditions such as MASLD likely resulted in misclassification of individuals with asymptomatic disease as non-cases. Such misclassification would be expected to bias associations in our discovery study, particularly for phase-two analyses that adjust for these clinical diagnoses.

Nevertheless, when we evaluated the predictive performance of risk-factor-based metabolomic scores among participants with documented risk-factor diagnoses, the results supported the robustness of the main findings for all cancers except iCCA. These analyses indicate that despite underdiagnosed clinical risk factors in UK Biobank, the metabolomic scores, except for iCCA, capture cancer-relevant metabolic disturbances not solely driven by coded diagnoses, and that much of the observed risk discrimination likely reflects underlying biological processes present before clinical recognition. For iCCA, however, the pattern among diagnosed patients suggests that the observed metabolite–iCCA associations are primarily driven by undiagnosed PSC and cirrhosis, both of which have strong and direct etiologic links to iCCA.

Second, because many precursor diseases progress silently for years, their true onset likely preceded the recorded clinical diagnosis, introducing another source of potential misclassification. In Cox proportional hazards models, such delays generally bias associations toward the null. However, given the relatively low event rates (<4%) for these clinical conditions, simulations by Oh *et al.* indicate that the magnitude of this underestimation is likely minimal.^46^ For hepatobiliary cancers, the effect is probably even smaller, as event rates were very low (∼0.1%) and the lag between disease onset and diagnosis is typically shorter.^45, 47, 48^ Although such delays may have attenuated weaker metabolite–risk-factor associations, the robustness of the metabolites prioritised in phase one argues against meaningful distortion. However, when interpreting the phase-one associations, differences in effect size across MASLD, MASH, and cirrhosis are better understood as reflecting underlying differences in biological severity already present at blood sampling, rather than as prospective predictors of disease progression.

Third, independent validation was performed in a separate UK Biobank batch, which helps mitigate concerns about technical artefacts, but does not fully address potential cohort-specific biases. Fourth, generalisability may also be limited, as UK Biobank participants are generally healthier, more educated, and less ethnically diverse than the broader population.^49^ This is particularly relevant for cancer risk stratification, where performance could differ substantially in cohorts with different baseline risk profiles, healthcare access, or genetic backgrounds. Finally, because the plasma levels of many metabolites are highly correlated, variable selection in multiple regression models can be shaped by this correlation structure, so non-selection should not be interpreted as evidence of absence. We mitigated this by clustering strongly correlated metabolites and analysing only cluster representatives (PC1 scores), which reduced redundancy while preserving major sources of variation.

Our findings have important implications for refining risk assessment in hepatobiliary cancers. The clearest potential for risk stratification emerged for HCC and iCCA, where metabolomic scores showed meaningful separation of cumulative incidence curves in the validation cohort. For HCC, integrating metabolomic and polygenic risk scores provided complementary information, suggesting added value when both types of markers are considered. In contrast, findings for GBC, eCCA, and AoV were limited by smaller case numbers and wide uncertainty, and should be interpreted as preliminary. Future work should prioritize external validation of these associations, particularly for lipid-related features, assessment of model calibration, and evaluation of whether sex- or aetiology-specific approaches improve prediction. Such evidence will be essential to determine whether metabolomic markers can ultimately contribute to targeted early-detection strategies for hepatobiliary cancers.

In summary, our risk factor-based approach identified metabolomic patterns that precede hepatobiliary cancers and persist after accounting for clinical risk factor diagnoses. The findings point to metabolic processes relevant to cancer susceptibility and add independent, reproducible markers, particularly for HCC and iCCA. Further validation in clinically enriched and more diverse populations, along with refinement of predictive models, is urgently needed to clarify how these metabolites may contribute to more precise risk assessment of hepatobiliary cancers in clinical practice.

## Data Availability

This research was conducted using data from UK Biobank under application number 58030. UK Biobank data are available to bona fide researchers from academic, charitable, government, and commercial institutions following project approval and completion of the required access procedures (https://www.ukbiobank.ac.uk). Access is granted for health‑related research in the public interest in accordance with UK Biobank’s data access policy.

## Disclosures

The authors declare that they have no competing interests.

## Declaration of Generative AI and AI-assisted technologies in the writing process

During the preparation of this work the author Felix Boekstegers used Copilot in order to improve the writing quality. After using this tool, the authors reviewed and edited the content as needed and take full responsibility for the content of the publication.

## IARC disclaimer

Where authors are identified as personnel of the International Agency for Research on Cancer/World Health Organization, the authors alone are responsible for the views expressed in this article and they do not necessarily represent the decisions, policy, or views of the International Agency for Research on Cancer/World Health Organization.

## Author Contributions

Felix Boekstegers, PhD (Conceptualization: Lead; Data curation: Lead; Formal analysis: Lead; Funding acquisition: Lead; Investigation: Lead; Methodology: Lead; Project administration: Lead;

Software: Lead; Validation: Lead; Visualization: Lead; Writing – original draft: Lead; Writing – review & editing: Lead)

Vivian Viallon, PhD (Resources: Equal; Methodology: Supporting; Software: Supporting; Validation: Supporting; Writing – review & editing: Supporting)

Marie Breur, PhD (Methodology: Supporting; Software: Supporting; Writing – review & editing: Supporting)

Cosmin Voican, MD (Resources: Supporting; Writing – review & editing: Supporting) Gabriel Perlemutter, MD (Resources: Supporting; Writing – review & editing: Supporting)

Chrysovalantou Chatziioannou, PhD (Writing – review & editing: Supporting)

Pekka Keski-Rahkonen, PhD (Resources: Supporting; Writing – review & editing: Supporting)

Dominique Scherer, PhD (Resources: Lead; Writing – review & editing: Supporting)

Mazda Jenab, PhD (Conceptualization: Equal; Funding acquisition: Equal; Methodology: Supporting; Resources: Lead; Supervision: Lead; Validation: Supporting; Writing – review & editing: Equal; Supervision: Lead)

Justo Lorenzo Bermjeo, PhD (Conceptualization: Equal; Funding acquisition: Equal; Methodology: Supporting; Resources: Lead; Supervision: Lead; Validation: Supporting; Writing – review & editing: Equal; Lead Supervision: Lead)

## Data Transparency Statement

This research was conducted using data from UK Biobank under application number 58030. UK Biobank data are available to bona fide researchers from academic, charitable, government, and commercial institutions following project approval and completion of the required access procedures (https://www.ukbiobank.ac.uk). Access is granted for health-related research in the public interest in accordance with UK Biobank’s data access policy.

The R and SAS scripts developed to implement the analyses are available on the GitHub platform at: https://github.com/boekstegersf/hepatobiliary_cancer_biomarker_discovery.

## Abbreviations

AoV: ampulla of Vater cancer
BMI: body mass index
eCCA: extrahepatic cholangiocarcinoma
GBC: gallbladder cancer
iCCA: intrahepatic cholangiocarcinoma
HCC: hepatocellular carcinoma
HDL: high-density lipoprotein
HR: hazard ratio
ICD10: the International Classification of Diseases, tenth revision, clinical modification
IDL: intermediate-density lipoprotein
LDL: low-density lipoprotein
MASH: metabolic dysfunction-associated steatohepatitis
MASLD: metabolic dysfunction-associated steatotic liver disease
NMR: nuclear magnetic resonance
OPCS-4: OPCS Classification of Interventions and Procedures version 4
OR: odds ratio
PC: principal component
PSC: primary sclerosing cholangitis
Pval: probability-value
SD: standard deviation
VLDL: very low-density lipoprotein

## Notes

**Grant Support:** Funded by the Deutsche Forschungsgemeinschaft (DFG, German Research Foundation) – project number 514150408 and the European Union’s Horizon 2020 research and innovation program (grant 825741). We also gratefully acknowledge the data storage service SDS@hd supported by the Ministry of Science, Research, and the Arts Baden-Württemberg (MWK) and the German Research Foundation (DFG) through grants INST 35/1314-1 FUGG and INST 35/1503-1 FUGG.

### Competing Interest Statement

The authors have declared no competing interest.

### Funding Statement

Funded by the Deutsche Forschungsgemeinschaft (DFG, German Research Foundation) - project number 514150408 and the European Union's Horizon 2020 research and innovation program (grant 825741). We also gratefully acknowledge the data storage service SDS@hd supported by the Ministry of Science, Research, and the Arts Baden-Wuerttemberg (MWK) and the German Research Foundation (DFG) through grants INST 35/1314-1 FUGG and INST 35/1503-1 FUGG.

### Author Declarations

YES. This study used human data obtained from the UK Biobank resource (Application 58030). UK Biobank provides anonymized human participant data to approved researchers following completion of required ethical and access procedures. This research involved only secondary analysis of these de‑identified human data. No direct contact with participants occurred.

