## Supplementary Tables for "Risk Factor–Based Metabolomic Profiling Reveals Plasma Biomarkers of Hepatobiliary Cancer"

**Supplementary Table S1.** Description of the targeted metabolite data from UK Biobank and EPIC.

| Study | Measurement period | Number of Samples | Sample type | Volume | Laboratory | Kit used |
|---|---|---|---|---|---|---|
| **Discovery: UK Biobank (Phase 1 and 2)** | Phase 1: June 2019 – April 2020 Phase 2: April 2020 – June 2022 | ca. 274,000 | EDTA plasma (aliquot 3) | ≥ 85 µL | Nightingale Health's clinical laboratory in Finland | Nightingale Health's high-throughput 1H-NMR metabolomics platform[1] |
| **Independent Validation: UK Biobank (Phase 3)** | Phase 3: September 2022 – November 2023 | ca. 217,000 | EDTA plasma (aliquot 3) | ≥ 85 µL | Nightingale Health's clinical laboratory in Finland | Nightingale Health's high-throughput 1H-NMR metabolomics platform[1] |

**Supplementary Table S2.** Endpoint definitions.

| Abbreviation | Description | Definition |
|---|---|---|
| **Gallstones** | Calculus of gallbladder | ICD-10 Codes K80.0 (Calculus of gallbladder with acute cholecystitis), K80.1 (Calculus of gallbladder with other cholecystitis), K80.2 (Calculus of gallbladder without cholecystitis) |
| **Cholecystitis** | Calculus of gallbladder with cholecystitis | ICD-10 Codes K80.0-1 or overlapping K80.2 and K81 (Cholecystitis) |
| **Cholecystectomy** | Calculus of gallbladder with cholecystectomy | ICD-10 Codes K80.0-2 and OPCS Classification of Interventions and Procedures version 4 (OPCS-4) J18 |
| **MASLD** | Metabolic dysfunction-associated steatotic liver disease | ICD-10 Code K76.0 (Hepatic steatosis) and no ICD-10 Code K70 (Alcoholic liver disease); along with at least one of the five metabolic syndrome criteria and a low alcohol consumption (less than 20 or 30 grams of alcohol consumption per day in women or men respectively), in accordance with the nomenclature agreed on by the by the multi-society Delphi consensus statement in 2023[1] |
| **MASH** | Metabolic dysfunction-associated steatohepatitis | ICD-10 code K75.8 (Other specified inflammatory liver diseases) |
| **Cirrhosis** | Cirrhosis of liver | ICD-10 codes K70.3 (Alcoholic cirrhosis), K71.7 (Toxic liver disease with cirrhosis), K74 (Liver fibrosis/cirrhosis) |
| **PSC** | Primary sclerosing cholangitis | ICD-10 code K83.0 (Other diseases of biliary tract: Cholangitis) |
| **GBC** | Gallbladder cancer | ICD-10 code C23 (Malignant neoplasm of gallbladder) |
| **HCC** | Hepatocellular carcinoma | ICD-10 code C22.0 (Liver cell carcinoma) |
| **eCCA** | Extrahepatic cholangiocarcinoma | ICD-10 code C24.0 (Malignant neoplasm of extrahepatic bile duct) |
| **iCCA** | Intrahepatic cholangiocarcinoma | ICD-10 code C22.1 (Malignant neoplasm of intrahepatic bile duct carcinoma) |
| **AoV** | Cancer of Ampulla of Vater | ICD-10 code C24.1 (Malignant neoplasm of Ampulla of Vater) |

[1]Lazarus JV, Newsome PN, Francque SM, et al. Reply: A multi-society Delphi consensus statement on new fatty liver disease nomenclature. Hepatology 2024;79:E93-E94.

**Supplementary Table S3.** Details on circulating metabolites provided with UK Biobank and the Fischer's ratio.

| Abbreviation | Description | Subgroup | Group |
|---|---|---|---|
| Ala | Alanine | Amino acids | Amino acids |
| Gln | Glutamine | Amino acids | Amino acids |
| Gly | Glycine | Amino acids | Amino acids |
| His | Histidine | Amino acids | Amino acids |
| Ile | Isoleucine | Branched-chain amino acids | Amino acids |
| Leu | Leucine | Branched-chain amino acids | Amino acids |
| Total BCAA | Total concentration of branched-chain amino acids (leucine + isoleucine + valine) | Branched-chain amino acids | Amino acids |
| Val | Valine | Branched-chain amino acids | Amino acids |
| Phe | Phenylalanine | Aromatic amino acids | Amino acids |
| Tyr | Tyrosine | Aromatic amino acids | Amino acids |
| ApoA1 | Apolipoprotein A1 | Apolipoproteins | Apolipoproteins |
| ApoB | Apolipoprotein B | Apolipoproteins | Apolipoproteins |
| Clinical LDL-C | Clinical LDL cholesterol | Cholesterol | Cholesterol |
| HDL-C | HDL cholesterol | Cholesterol | Cholesterol |
| LDL-C | LDL cholesterol | Cholesterol | Cholesterol |
| non-HDL-C | Total cholesterol minus HDL-C | Cholesterol | Cholesterol |
| Remnant-C | Remnant cholesterol (non-HDL, non-LDL -cholesterol) | Cholesterol | Cholesterol |
| Total-C | Total cholesterol | Cholesterol | Cholesterol |
| VLDL-C | VLDL cholesterol | Cholesterol | Cholesterol |
| HDL-CE | Cholesteryl esters in HDL | Cholesteryl esters | Cholesteryl esters |
| LDL-CE | Cholesteryl esters in LDL | Cholesteryl esters | Cholesteryl esters |
| Total-CE | Total esterified cholesterol | Cholesteryl esters | Cholesteryl esters |
| VLDL-CE | Cholesteryl esters in VLDL | Cholesteryl esters | Cholesteryl esters |
| DHA | Docosahexaenoic acid | Fatty acids | Fatty acids |
| LA | Linoleic acid | Fatty acids | Fatty acids |
| MUFA | Monounsaturated fatty acids | Fatty acids | Fatty acids |
| Omega-3 | Omega-3 fatty acids | Fatty acids | Fatty acids |
| Omega-6 | Omega-6 fatty acids | Fatty acids | Fatty acids |
| PUFA | Polyunsaturated fatty acids | Fatty acids | Fatty acids |
| SFA | Saturated fatty acids | Fatty acids | Fatty acids |
| Total-FA | Total fatty acids | Fatty acids | Fatty acids |
| Unsaturation | Degree of unsaturation | Fatty acids | Fatty acids |
| Albumin | Albumin | Fluid balance | Fluid balance |
| Creatinine | Creatinine | Fluid balance | Fluid balance |
| HDL-FC | Free cholesterol in HDL | Free cholesterol | Free cholesterol |
| LDL-FC | Free cholesterol in LDL | Free cholesterol | Free cholesterol |
| Total-FC | Total free cholesterol | Free cholesterol | Free cholesterol |
| VLDL-FC | Free cholesterol in VLDL | Free cholesterol | Free cholesterol |
| Citrate | Citrate | Glycolysis related metabolites | Glycolysis related metabolites |
| Glucose | Glucose | Glycolysis related metabolites | Glycolysis related metabolites |
| Lactate | Lactate | Glycolysis related metabolites | Glycolysis related metabolites |
| Pyruvate | Pyruvate | Glycolysis related metabolites | Glycolysis related metabolites |
| GlycA | Glycoprotein acetyls | Inflammation | Inflammation |
| Acetate | Acetate | Ketone bodies | Ketone bodies |
| Acetoacetate | Acetoacetate | Ketone bodies | Ketone bodies |
| Acetone | Acetone | Ketone bodies | Ketone bodies |
| bOHbutyrate | 3-Hydroxybutyrate | Ketone bodies | Ketone bodies |

**Supplementary Table S3 (cont).** Details on circulating metabolites provided with UK Biobank and the Fischer's ratio.

| Abbreviation | Description | Subgroup | Group |
|---|---|---|---|
| HDL-P | Concentration of HDL particles | Lipoprotein particle concentrations | Lipoprotein particle concentrations |
| LDL-P | Concentration of LDL particles | Lipoprotein particle concentrations | Lipoprotein particle concentrations |
| Total-P | Total concentration of lipoprotein particles | Lipoprotein particle concentrations | Lipoprotein particle concentrations |
| VLDL-P | Concentration of VLDL particles | Lipoprotein particle concentrations | Lipoprotein particle concentrations |
| HDL size | Average diameter for HDL particles | Lipoprotein particle sizes | Lipoprotein particle sizes |
| LDL size | Average diameter for LDL particles | Lipoprotein particle sizes | Lipoprotein particle sizes |
| VLDL size | Average diameter for VLDL particles | Lipoprotein particle sizes | Lipoprotein particle sizes |
| XXL-VLDL-C | Cholesterol in chylomicrons and extremely large VLDL | Chylomicrons and extremely large VLDL | Lipoprotein subclasses |
| XXL-VLDL-CE | Cholesteryl esters in chylomicrons and extremely large VLDL | Chylomicrons and extremely large VLDL | Lipoprotein subclasses |
| XXL-VLDL-FC | Free cholesterol in chylomicrons and extremely large VLDL | Chylomicrons and extremely large VLDL | Lipoprotein subclasses |
| XXL-VLDL-L | Total lipids in chylomicrons and extremely large VLDL | Chylomicrons and extremely large VLDL | Lipoprotein subclasses |
| XXL-VLDL-P | Concentration of chylomicrons and extremely large VLDL particles | Chylomicrons and extremely large VLDL | Lipoprotein subclasses |
| XXL-VLDL-PL | Phospholipids in chylomicrons and extremely large VLDL | Chylomicrons and extremely large VLDL | Lipoprotein subclasses |
| XXL-VLDL-TG | Triglycerides in chylomicrons and extremely large VLDL | Chylomicrons and extremely large VLDL | Lipoprotein subclasses |
| XL-VLDL-C | Cholesterol in very large VLDL | Very large VLDL | Lipoprotein subclasses |
| XL-VLDL-CE | Cholesteryl esters in very large VLDL | Very large VLDL | Lipoprotein subclasses |
| XL-VLDL-FC | Free cholesterol in very large VLDL | Very large VLDL | Lipoprotein subclasses |
| XL-VLDL-L | Total lipids in very large VLDL | Very large VLDL | Lipoprotein subclasses |
| XL-VLDL-P | Concentration of very large VLDL particles | Very large VLDL | Lipoprotein subclasses |
| XL-VLDL-PL | Phospholipids in very large VLDL | Very large VLDL | Lipoprotein subclasses |
| XL-VLDL-TG | Triglycerides in very large VLDL | Very large VLDL | Lipoprotein subclasses |
| L-VLDL-C | Cholesterol in large VLDL | Large VLDL | Lipoprotein subclasses |
| L-VLDL-CE | Cholesteryl esters in large VLDL | Large VLDL | Lipoprotein subclasses |
| L-VLDL-FC | Free cholesterol in large VLDL | Large VLDL | Lipoprotein subclasses |
| L-VLDL-L | Total lipids in large VLDL | Large VLDL | Lipoprotein subclasses |
| L-VLDL-P | Concentration of large VLDL particles | Large VLDL | Lipoprotein subclasses |
| L-VLDL-PL | Phospholipids in large VLDL | Large VLDL | Lipoprotein subclasses |
| L-VLDL-TG | Triglycerides in large VLDL | Large VLDL | Lipoprotein subclasses |
| M-VLDL-C | Cholesterol in medium VLDL | Medium VLDL | Lipoprotein subclasses |
| M-VLDL-CE | Cholesteryl esters in medium VLDL | Medium VLDL | Lipoprotein subclasses |
| M-VLDL-FC | Free cholesterol in medium VLDL | Medium VLDL | Lipoprotein subclasses |
| M-VLDL-L | Total lipids in medium VLDL | Medium VLDL | Lipoprotein subclasses |
| M-VLDL-P | Concentration of medium VLDL particles | Medium VLDL | Lipoprotein subclasses |
| M-VLDL-PL | Phospholipids in medium VLDL | Medium VLDL | Lipoprotein subclasses |
| M-VLDL-TG | Triglycerides in medium VLDL | Medium VLDL | Lipoprotein subclasses |
| S-VLDL-C | Cholesterol in small VLDL | Small VLDL | Lipoprotein subclasses |
| S-VLDL-CE | Cholesteryl esters in small VLDL | Small VLDL | Lipoprotein subclasses |
| S-VLDL-FC | Free cholesterol in small VLDL | Small VLDL | Lipoprotein subclasses |
| S-VLDL-L | Total lipids in small VLDL | Small VLDL | Lipoprotein subclasses |
| S-VLDL-P | Concentration of small VLDL particles | Small VLDL | Lipoprotein subclasses |

**Supplementary Table S3 (cont).** Details on circulating metabolites provided with UK Biobank and the Fischer's ratio.

| Abbreviation | Description | Subgroup | Group |
|---|---|---|---|
| S-VLDL-PL | Phospholipids in small VLDL | Small VLDL | Lipoprotein subclasses |
| S-VLDL-TG | Triglycerides in small VLDL | Small VLDL | Lipoprotein subclasses |
| XS-VLDL-C | Cholesterol in very small VLDL | Very small VLDL | Lipoprotein subclasses |
| XS-VLDL-CE | Cholesteryl esters in very small VLDL | Very small VLDL | Lipoprotein subclasses |
| XS-VLDL-FC | Free cholesterol in very small VLDL | Very small VLDL | Lipoprotein subclasses |
| XS-VLDL-L | Total lipids in very small VLDL | Very small VLDL | Lipoprotein subclasses |
| XS-VLDL-P | Concentration of very small VLDL particles | Very small VLDL | Lipoprotein subclasses |
| XS-VLDL-PL | Phospholipids in very small VLDL | Very small VLDL | Lipoprotein subclasses |
| XS-VLDL-TG | Triglycerides in very small VLDL | Very small VLDL | Lipoprotein subclasses |
| IDL-C | Cholesterol in IDL | IDL | Lipoprotein subclasses |
| IDL-CE | Cholesteryl esters in IDL | IDL | Lipoprotein subclasses |
| IDL-FC | Free cholesterol in IDL | IDL | Lipoprotein subclasses |
| IDL-L | Total lipids in IDL | IDL | Lipoprotein subclasses |
| IDL-P | Concentration of IDL particles | IDL | Lipoprotein subclasses |
| IDL-PL | Phospholipids in IDL | IDL | Lipoprotein subclasses |
| IDL-TG | Triglycerides in IDL | IDL | Lipoprotein subclasses |
| L-LDL-C | Cholesterol in large LDL | Large LDL | Lipoprotein subclasses |
| L-LDL-CE | Cholesteryl esters in large LDL | Large LDL | Lipoprotein subclasses |
| L-LDL-FC | Free cholesterol in large LDL | Large LDL | Lipoprotein subclasses |
| L-LDL-L | Total lipids in large LDL | Large LDL | Lipoprotein subclasses |
| L-LDL-P | Concentration of large LDL particles | Large LDL | Lipoprotein subclasses |
| L-LDL-PL | Phospholipids in large LDL | Large LDL | Lipoprotein subclasses |
| L-LDL-TG | Triglycerides in large LDL | Large LDL | Lipoprotein subclasses |
| M-LDL-C | Cholesterol in medium LDL | Medium LDL | Lipoprotein subclasses |
| M-LDL-CE | Cholesteryl esters in medium LDL | Medium LDL | Lipoprotein subclasses |
| M-LDL-FC | Free cholesterol in medium LDL | Medium LDL | Lipoprotein subclasses |
| M-LDL-L | Total lipids in medium LDL | Medium LDL | Lipoprotein subclasses |
| M-LDL-P | Concentration of medium LDL particles | Medium LDL | Lipoprotein subclasses |
| M-LDL-PL | Phospholipids in medium LDL | Medium LDL | Lipoprotein subclasses |
| M-LDL-TG | Triglycerides in medium LDL | Medium LDL | Lipoprotein subclasses |
| S-LDL-C | Cholesterol in small LDL | Small LDL | Lipoprotein subclasses |
| S-LDL-CE | Cholesteryl esters in small LDL | Small LDL | Lipoprotein subclasses |
| S-LDL-FC | Free cholesterol in small LDL | Small LDL | Lipoprotein subclasses |
| S-LDL-L | Total lipids in small LDL | Small LDL | Lipoprotein subclasses |
| S-LDL-P | Concentration of small LDL particles | Small LDL | Lipoprotein subclasses |
| S-LDL-PL | Phospholipids in small LDL | Small LDL | Lipoprotein subclasses |
| S-LDL-TG | Triglycerides in small LDL | Small LDL | Lipoprotein subclasses |
| XL-HDL-C | Cholesterol in very large HDL | Very large HDL | Lipoprotein subclasses |
| XL-HDL-CE | Cholesteryl esters in very large HDL | Very large HDL | Lipoprotein subclasses |
| XL-HDL-FC | Free cholesterol in very large HDL | Very large HDL | Lipoprotein subclasses |
| XL-HDL-L | Total lipids in very large HDL | Very large HDL | Lipoprotein subclasses |
| XL-HDL-P | Concentration of very large HDL particles | Very large HDL | Lipoprotein subclasses |
| XL-HDL-PL | Phospholipids in very large HDL | Very large HDL | Lipoprotein subclasses |
| XL-HDL-TG | Triglycerides in very large HDL | Very large HDL | Lipoprotein subclasses |
| L-HDL-C | Cholesterol in large HDL | Large HDL | Lipoprotein subclasses |
| L-HDL-CE | Cholesteryl esters in large HDL | Large HDL | Lipoprotein subclasses |
| L-HDL-FC | Free cholesterol in large HDL | Large HDL | Lipoprotein subclasses |
| L-HDL-L | Total lipids in large HDL | Large HDL | Lipoprotein subclasses |

**Supplementary Table S3 (cont).** Details on circulating metabolites provided with UK Biobank and the Fischer's ratio.

| Abbreviation | Description | Subgroup | Group |
|---|---|---|---|
| L-HDL-P | Concentration of large HDL particles | Large HDL | Lipoprotein subclasses |
| L-HDL-PL | Phospholipids in large HDL | Large HDL | Lipoprotein subclasses |
| L-HDL-TG | Triglycerides in large HDL | Large HDL | Lipoprotein subclasses |
| M-HDL-C | Cholesterol in medium HDL | Medium HDL | Lipoprotein subclasses |
| M-HDL-CE | Cholesteryl esters in medium HDL | Medium HDL | Lipoprotein subclasses |
| M-HDL-FC | Free cholesterol in medium HDL | Medium HDL | Lipoprotein subclasses |
| M-HDL-L | Total lipids in medium HDL | Medium HDL | Lipoprotein subclasses |
| M-HDL-P | Concentration of medium HDL particles | Medium HDL | Lipoprotein subclasses |
| M-HDL-PL | Phospholipids in medium HDL | Medium HDL | Lipoprotein subclasses |
| M-HDL-TG | Triglycerides in medium HDL | Medium HDL | Lipoprotein subclasses |
| S-HDL-C | Cholesterol in small HDL | Small HDL | Lipoprotein subclasses |
| S-HDL-CE | Cholesteryl esters in small HDL | Small HDL | Lipoprotein subclasses |
| S-HDL-FC | Free cholesterol in small HDL | Small HDL | Lipoprotein subclasses |
| S-HDL-L | Total lipids in small HDL | Small HDL | Lipoprotein subclasses |
| S-HDL-P | Concentration of small HDL particles | Small HDL | Lipoprotein subclasses |
| S-HDL-PL | Phospholipids in small HDL | Small HDL | Lipoprotein subclasses |
| S-HDL-TG | Triglycerides in small HDL | Small HDL | Lipoprotein subclasses |
| Cholines | Total cholines | Other lipids | Other lipids |
| Phosphatidylc | Phosphatidylcholines | Other lipids | Other lipids |
| Phosphoglyc | Phosphoglycerides | Other lipids | Other lipids |
| Sphingomyelins | Sphingomyelins | Other lipids | Other lipids |
| HDL-PL | Phospholipids in HDL | Phospholipids | Phospholipids |
| LDL-PL | Phospholipids in LDL | Phospholipids | Phospholipids |
| Total-PL | Total phospholipids in lipoprotein particles | Phospholipids | Phospholipids |
| VLDL-PL | Phospholipids in VLDL | Phospholipids | Phospholipids |
| HDL-L | Total lipids in HDL | Total lipids | Total lipids |
| LDL-L | Total lipids in LDL | Total lipids | Total lipids |
| Total-L | Total lipids in lipoprotein particles | Total lipids | Total lipids |
| VLDL-L | Total lipids in VLDL | Total lipids | Total lipids |
| HDL-TG | Triglycerides in HDL | Triglycerides | Triglycerides |
| LDL-TG | Triglycerides in LDL | Triglycerides | Triglycerides |
| Total-TG | Total triglycerides | Triglycerides | Triglycerides |
| VLDL-TG | Triglycerides in VLDL | Triglycerides | Triglycerides |
| FR | Fischer’s ratio[1] | Amino acids | Amino acids |

[1]Fisher’s ratio is defined as sum of branched-chain amino acids (Leu, Ile and Val) divided by the sum of aromatic amino acids (Tyr, Phe).

**Supplementary Table S4.** Metabolites Associated with any of the Gallstones-related Clinical Conditions (Gallstones, Cholecystitis and Cholecystectomy) in a Univariate Cox Model (Pre-Phase 1).

| | Univariate Cox regression results for gallstones[1] | | | Univariate Cox regression results for cholecystitis[1] | | | Univariate Cox regression results for cholecystecomy[1] | | | |
|---|---|---|---|---|---|---|---|---|---|---|
| **Metabolite** | **Est** | **Se** | **Pval-BH** | **Est** | **Se** | **Pval-BH** | **Est** | **Se** | **Pval-BH** | **Included for phase 1?[2]** |
| LDL_C cluster | -0.092 | 0.009 | < 0.00001 | -0.075 | 0.013 | < 0.00001 | -0.031 | 0.013 | 0.02 | yes |
| XXL-VLDL-TG cluster | 0.109 | 0.011 | < 0.00001 | 0.114 | 0.015 | < 0.00001 | 0.148 | 0.014 | < 0.00001 | yes |
| Average diameter for LDL particles | -0.068 | 0.010 | < 0.00001 | -0.062 | 0.014 | 0.00003 | -0.063 | 0.013 | < 0.00001 | yes |
| L-HDL-C cluster | -0.162 | 0.010 | < 0.00001 | -0.170 | 0.014 | < 0.00001 | -0.184 | 0.012 | < 0.00001 | yes |
| Phosphatidylc cluster | -0.150 | 0.010 | < 0.00001 | -0.141 | 0.015 | < 0.00001 | -0.114 | 0.013 | < 0.00001 | yes |
| Sphingomyelins | -0.159 | 0.011 | < 0.00001 | -0.147 | 0.015 | < 0.00001 | -0.117 | 0.013 | < 0.00001 | yes |
| ApoB cluster | -0.060 | 0.010 | < 0.00001 | -0.039 | 0.014 | 0.008 | 0.010 | 0.013 | 0.47 | yes |
| Cluster 8 (ApoA1 cluster | -0.160 | 0.008 | < 0.00001 | -0.163 | 0.011 | < 0.00001 | -0.163 | 0.010 | < 0.00001 | yes |
| Degree of unsaturation | -0.174 | 0.010 | < 0.00001 | -0.146 | 0.015 | < 0.00001 | -0.144 | 0.013 | < 0.00001 | yes |
| Omega-3 fatty acids | -0.148 | 0.010 | < 0.00001 | -0.127 | 0.014 | < 0.00001 | -0.102 | 0.013 | < 0.00001 | yes |
| Omega-6 cluster | -0.094 | 0.010 | < 0.00001 | -0.073 | 0.014 | < 0.00001 | -0.033 | 0.013 | 0.02 | yes |
| LDL-TG cluster | 0.026 | 0.010 | 0.02 | 0.020 | 0.014 | 0.19 | 0.043 | 0.013 | 0.001 | yes |
| SFA cluster | -0.045 | 0.010 | 0.00004 | -0.043 | 0.015 | 0.005 | -0.006 | 0.013 | 0.68 | yes |
| Docosahexaenoic acid | -0.168 | 0.009 | < 0.00001 | -0.153 | 0.012 | < 0.00001 | -0.140 | 0.011 | < 0.00001 | yes |
| Alanine | 0.026 | 0.010 | 0.02 | 0.026 | 0.014 | 0.09 | 0.030 | 0.013 | 0.02 | yes |
| Glutamine | 0.006 | 0.010 | 0.63 | 0.019 | 0.014 | 0.21 | 0.026 | 0.013 | 0.06 | no |
| Glycine | -0.011 | 0.009 | 0.31 | 0.001 | 0.014 | 0.93 | -0.011 | 0.012 | 0.41 | no |
| Histidine | -0.023 | 0.010 | 0.03 | 0.003 | 0.014 | 0.83 | 0.016 | 0.012 | 0.25 | yes |
| Valine cluster | -0.001 | 0.010 | 0.96 | -0.005 | 0.015 | 0.74 | 0.030 | 0.013 | 0.02 | yes |
| Phenylalanine | 0.013 | 0.010 | 0.22 | 0.014 | 0.014 | 0.38 | 0.009 | 0.012 | 0.50 | no |
| Tyrosine | -0.037 | 0.010 | 0.0005 | -0.051 | 0.014 | 0.0007 | -0.043 | 0.013 | 0.001 | yes |
| Glucose | 0.019 | 0.010 | 0.08 | -0.035 | 0.015 | 0.02 | -0.022 | 0.013 | 0.12 | yes |
| Lactate | 0.022 | 0.010 | 0.04 | 0.034 | 0.014 | 0.02 | 0.030 | 0.012 | 0.02 | yes |
| Pyruvate | 0.008 | 0.010 | 0.50 | 0.015 | 0.014 | 0.34 | 0.017 | 0.013 | 0.22 | no |
| Citrate | 0.003 | 0.010 | 0.79 | -0.013 | 0.014 | 0.41 | -0.033 | 0.013 | 0.01 | yes |
| 3-Hydroxybutyrate | -0.017 | 0.010 | 0.10 | -0.030 | 0.013 | 0.03 | -0.033 | 0.011 | 0.004 | yes |
| Acetate | -0.020 | 0.009 | 0.04 | -0.035 | 0.012 | 0.006 | -0.025 | 0.011 | 0.03 | yes |
| Acetoacetate | -0.016 | 0.010 | 0.13 | -0.018 | 0.014 | 0.21 | -0.023 | 0.012 | 0.06 | no |
| Acetone | -0.049 | 0.010 | < 0.00001 | -0.078 | 0.015 | < 0.00001 | -0.082 | 0.013 | < 0.00001 | yes |
| Creatinine | 0.048 | 0.012 | 0.0001 | 0.026 | 0.017 | 0.15 | 0.039 | 0.015 | 0.01 | yes |
| Albumin | -0.038 | 0.003 | < 0.00001 | -0.036 | 0.005 | < 0.00001 | -0.031 | 0.006 | < 0.00001 | yes |
| VLDL-TG cluster | 0.083 | 0.010 | < 0.00001 | 0.083 | 0.015 | < 0.00001 | 0.115 | 0.013 | < 0.00001 | yes |
| L-VLDL-TG cluster | 0.077 | 0.011 | < 0.00001 | 0.087 | 0.015 | < 0.00001 | 0.141 | 0.014 | < 0.00001 | yes |
| VLDL-C cluster | -0.002 | 0.010 | 0.89 | 0.016 | 0.014 | 0.32 | 0.067 | 0.013 | < 0.00001 | yes |
| M-LDL-P cluster | -0.035 | 0.010 | 0.0009 | -0.015 | 0.014 | 0.32 | 0.038 | 0.013 | 0.005 | yes |
| VLDL-P cluster | 0.027 | 0.010 | 0.01 | 0.041 | 0.014 | 0.007 | 0.088 | 0.013 | < 0.00001 | yes |
| XS-VLDL-P cluster | -0.025 | 0.010 | 0.02 | -0.021 | 0.014 | 0.18 | 0.012 | 0.013 | 0.38 | yes |
| XS-VLDL-C cluster | -0.073 | 0.010 | < 0.00001 | -0.060 | 0.014 | 0.00006 | -0.024 | 0.013 | 0.08 | yes |
| IDL-C cluster | -0.129 | 0.010 | < 0.00001 | -0.108 | 0.014 | < 0.00001 | -0.068 | 0.013 | < 0.00001 | yes |
| LDL-FC cluster | -0.046 | 0.004 | < 0.00001 | -0.044 | 0.006 | < 0.00001 | -0.034 | 0.008 | 0.00001 | yes |
| M-LDL-C cluster | -0.049 | 0.007 | < 0.00001 | -0.040 | 0.011 | 0.0003 | 0.006 | 0.013 | 0.68 | yes |
| XL-HDL-C cluster | -0.100 | 0.010 | < 0.00001 | -0.102 | 0.013 | < 0.00001 | -0.116 | 0.012 | < 0.00001 | yes |
| Triglycerides in very large HDL | 0.006 | 0.010 | 0.61 | 0.003 | 0.015 | 0.87 | 0.006 | 0.013 | 0.68 | no |
| Triglycerides in large HDL | -0.016 | 0.011 | 0.16 | -0.022 | 0.015 | 0.17 | -0.031 | 0.013 | 0.02 | yes |
| HDL-TG cluster | 0.042 | 0.010 | 0.00008 | 0.038 | 0.014 | 0.01 | 0.050 | 0.013 | 0.0001 | yes |

| | Univariate Cox regression results for gallstones[1] | | | Univariate Cox regression results for cholecystitis[1] | | | Univariate Cox regression results for cholecystecomy[1] | | | |
|---|---|---|---|---|---|---|---|---|---|---|
| **Metabolite** | **Est** | **Se** | **Pval-BH** | **Est** | **Se** | **Pval-BH** | **Est** | **Se** | **Pval-BH** | **Included for phase 1?[2]** |
| S-HDL-C cluster | -0.111 | 0.008 | < 0.00001 | -0.098 | 0.012 | < 0.00001 | -0.062 | 0.012 | < 0.00001 | yes |
| S-HDL-L cluster | -0.096 | 0.010 | < 0.00001 | -0.087 | 0.014 | < 0.00001 | -0.054 | 0.013 | 0.00004 | yes |
| Free cholesterol in small HDL | -0.137 | 0.010 | < 0.00001 | -0.126 | 0.014 | < 0.00001 | -0.097 | 0.013 | < 0.00001 | yes |
| HDL-C cluster | -0.225 | 0.011 | < 0.00001 | -0.225 | 0.016 | < 0.00001 | -0.231 | 0.014 | < 0.00001 | yes |
| HDL-P cluster | -0.196 | 0.011 | < 0.00001 | -0.186 | 0.015 | < 0.00001 | -0.165 | 0.013 | < 0.00001 | yes |
| Fischer's ratio | 0.018 | 0.011 | 0.12 | 0.022 | 0.015 | 0.16 | 0.065 | 0.013 | < 0.00001 | yes |

[1] Adjusted by age at blood collection, sex, fasting time, and menopause and use of exogenous hormones for women
[2] There was at least one metabolite-risk factor association with Benjamini-Hochberg adjusted p-value < 0.05
**Abbreviations:** Est, Estimate; Pval-BH, Benjamini-Hochberg adjusted P-value; Se, Standard error;

**Supplementary Table S5.** Metabolites Associated with Primary Sclerosing Cholangitis (PSC) in a Univariate Cox Model (Pre-Phase 1).

| | Univariate Cox regression results for PSC[1] | | | |
|---|---|---|---|---|
| **Metabolite** | **Est** | **Se** | **Pval-BH** | **Included for phase 1?[2]** |
| LDL_C cluster | -0.151 | 0.016 | < 0.00001 | yes |
| XXL-VLDL-TG cluster | 0.034 | 0.035 | 0.38 | no |
| Average diameter for LDL particles | -0.107 | 0.032 | 0.001 | yes |
| L-HDL-C cluster | -0.090 | 0.033 | 0.01 | yes |
| Phosphatidylc cluster | -0.175 | 0.033 | < 0.00001 | yes |
| Sphingomyelins | -0.206 | 0.033 | < 0.00001 | yes |
| ApoB cluster | -0.206 | 0.030 | < 0.00001 | yes |
| Cluster 8 (ApoA1 cluster | -0.144 | 0.028 | < 0.00001 | yes |
| Degree of unsaturation | -0.180 | 0.033 | < 0.00001 | yes |
| Omega-3 fatty acids | -0.124 | 0.033 | 0.0002 | yes |
| Omega-6 cluster | -0.196 | 0.032 | < 0.00001 | yes |
| LDL-TG cluster | 0.023 | 0.033 | 0.52 | no |
| SFA cluster | -0.078 | 0.033 | 0.02 | yes |
| Docosahexaenoic acid | -0.165 | 0.028 | < 0.00001 | yes |
| Alanine | 0.075 | 0.033 | 0.03 | yes |
| Glutamine | -0.066 | 0.032 | 0.06 | no |
| Glycine | -0.047 | 0.022 | 0.04 | yes |
| Histidine | -0.071 | 0.033 | 0.04 | yes |
| Valine cluster | -0.097 | 0.034 | 0.008 | yes |
| Phenylalanine | 0.033 | 0.033 | 0.36 | no |
| Tyrosine | 0.027 | 0.033 | 0.46 | no |
| Glucose | 0.044 | 0.032 | 0.21 | no |
| Lactate | 0.053 | 0.033 | 0.13 | no |
| Pyruvate | 0.028 | 0.033 | 0.45 | no |
| Citrate | 0.036 | 0.033 | 0.33 | no |
| 3-Hydroxybutyrate | 0.009 | 0.036 | 0.83 | no |
| Acetate | -0.014 | 0.030 | 0.68 | no |
| Acetoacetate | 0.001 | 0.035 | 0.98 | no |
| Acetone | -0.035 | 0.033 | 0.34 | no |
| Creatinine | 0.018 | 0.038 | 0.67 | no |
| Albumin | -0.043 | 0.007 | < 0.00001 | yes |
| VLDL-TG cluster | 0.051 | 0.033 | 0.16 | no |
| L-VLDL-TG cluster | -0.042 | 0.033 | 0.24 | no |
| VLDL-C cluster | -0.129 | 0.031 | 0.00006 | yes |
| M-LDL-P cluster | -0.177 | 0.031 | < 0.00001 | yes |
| VLDL-P cluster | -0.082 | 0.032 | 0.02 | yes |
| XS-VLDL-P cluster | -0.071 | 0.032 | 0.04 | yes |
| XS-VLDL-C cluster | -0.152 | 0.032 | < 0.00001 | yes |
| IDL-C cluster | -0.243 | 0.031 | < 0.00001 | yes |
| LDL-FC cluster | -0.056 | 0.008 | < 0.00001 | yes |
| M-LDL-C cluster | -0.076 | 0.010 | < 0.00001 | yes |
| XL-HDL-C cluster | -0.074 | 0.030 | 0.02 | yes |
| Triglycerides in very large HDL | 0.053 | 0.033 | 0.14 | no |
| Triglycerides in large HDL | 0.090 | 0.035 | 0.01 | yes |
| HDL-TG cluster | 0.088 | 0.034 | 0.01 | yes |
| S-HDL-C cluster | -0.226 | 0.020 | < 0.00001 | yes |

| | Univariate Cox regression results for PSC[1] | | | |
|---|---|---|---|---|
| **Metabolite** | **Est** | **Se** | **Pval-BH** | **Included for phase 1?[2]** |
| S-HDL-L cluster | -0.165 | 0.032 | < 0.00001 | yes |
| Free cholesterol in small HDL | -0.197 | 0.032 | < 0.00001 | yes |
| HDL-C cluster | -0.174 | 0.036 | < 0.00001 | yes |
| HDL-P cluster | -0.225 | 0.033 | < 0.00001 | yes |
| Fischer's ratio | -0.151 | 0.033 | 0.00001 | yes |

[1] Adjusted by age at blood collection, sex, fasting time, and menopause and use of exogenous hormones for women

[2] The metabolite-risk factor association with Benjamini-Hochberg adjusted p-value < 0.05

**Abbreviations:** Est, Estimate; Pval-BH, Benjamini-Hochberg adjusted P-value; Se, Standard error;

**Supplementary Table S6.** Metabolites Associated with any of the Spectrum of Metabolic Liver Disease (MASLD, MASH, Cirrhosis) in a Univariate Cox Model (Pre-Phase 1).

| | Univariate Cox regression results for MASLD[1] | | | Univariate Cox regression results for MASH[1] | | | Univariate Cox regression results for cirrhosis[1] | | | |
|---|---|---|---|---|---|---|---|---|---|---|
| **Metabolite** | **Est** | **Se** | **Pval-BH** | **Est** | **Se** | **Pval-BH** | **Est** | **Se** | **Pval-BH** | **Included for phase 1?[2]** |
| LDL_C cluster | -0.155 | 0.009 | < 0.00001 | -0.200 | 0.014 | < 0.00001 | -0.183 | 0.008 | < 0.00001 | yes |
| XXL-VLDL-TG cluster | 0.157 | 0.021 | < 0.00001 | 0.040 | 0.056 | 0.52 | -0.092 | 0.027 | 0.0009 | yes |
| Average diameter for LDL particles | -0.220 | 0.018 | < 0.00001 | -0.349 | 0.048 | < 0.00001 | -0.041 | 0.027 | 0.16 | yes |
| L-HDL-C cluster | -0.235 | 0.014 | < 0.00001 | -0.018 | 0.059 | 0.79 | 0.488 | 0.032 | < 0.00001 | yes |
| Phosphatidylc cluster | -0.221 | 0.018 | < 0.00001 | -0.221 | 0.052 | 0.00004 | 0.012 | 0.028 | 0.71 | yes |
| Sphingomyelins | -0.317 | 0.019 | < 0.00001 | -0.495 | 0.052 | < 0.00001 | -0.161 | 0.028 | < 0.00001 | yes |
| ApoB cluster | -0.185 | 0.018 | < 0.00001 | -0.437 | 0.036 | < 0.00001 | -0.365 | 0.021 | < 0.00001 | yes |
| Cluster 8 (ApoA1 cluster | -0.187 | 0.011 | < 0.00001 | -0.107 | 0.052 | 0.05 | 0.095 | 0.029 | 0.002 | yes |
| Degree of unsaturation | -0.284 | 0.019 | < 0.00001 | -0.498 | 0.051 | < 0.00001 | -0.493 | 0.026 | < 0.00001 | yes |
| Omega-3 fatty acids | -0.107 | 0.019 | < 0.00001 | -0.234 | 0.049 | < 0.00001 | -0.338 | 0.021 | < 0.00001 | yes |
| Omega-6 cluster | -0.183 | 0.019 | < 0.00001 | -0.395 | 0.047 | < 0.00001 | -0.282 | 0.026 | < 0.00001 | yes |
| LDL-TG cluster | 0.132 | 0.019 | < 0.00001 | 0.154 | 0.053 | 0.005 | 0.211 | 0.026 | < 0.00001 | yes |
| SFA cluster | -0.012 | 0.019 | 0.57 | -0.081 | 0.054 | 0.16 | -0.036 | 0.027 | 0.22 | no |
| Docosahexaenoic acid | -0.183 | 0.015 | < 0.00001 | -0.251 | 0.032 | < 0.00001 | -0.232 | 0.018 | < 0.00001 | yes |
| Alanine | 0.215 | 0.019 | < 0.00001 | 0.410 | 0.054 | < 0.00001 | 0.097 | 0.027 | 0.0007 | yes |
| Glutamine | -0.048 | 0.019 | 0.01 | -0.295 | 0.047 | < 0.00001 | -0.372 | 0.023 | < 0.00001 | yes |
| Glycine | -0.033 | 0.015 | 0.04 | -0.079 | 0.029 | 0.009 | -0.047 | 0.018 | 0.01 | yes |
| Histidine | -0.034 | 0.019 | 0.09 | 0.037 | 0.053 | 0.52 | -0.136 | 0.027 | < 0.00001 | yes |
| Valine cluster | 0.123 | 0.019 | < 0.00001 | 0.106 | 0.055 | 0.07 | -0.312 | 0.029 | < 0.00001 | yes |
| Phenylalanine | 0.077 | 0.019 | 0.00006 | 0.274 | 0.049 | < 0.00001 | 0.225 | 0.026 | < 0.00001 | yes |
| Tyrosine | 0.069 | 0.019 | 0.0004 | 0.497 | 0.052 | < 0.00001 | 0.454 | 0.026 | < 0.00001 | yes |
| Glucose | 0.176 | 0.017 | < 0.00001 | 0.468 | 0.035 | < 0.00001 | 0.263 | 0.022 | < 0.00001 | yes |
| Lactate | 0.097 | 0.019 | < 0.00001 | -0.029 | 0.053 | 0.62 | -0.001 | 0.027 | 0.98 | yes |
| Pyruvate | 0.013 | 0.019 | 0.52 | -0.007 | 0.053 | 0.90 | 0.009 | 0.027 | 0.75 | no |
| Citrate | 0.096 | 0.019 | < 0.00001 | 0.448 | 0.051 | < 0.00001 | 0.343 | 0.027 | < 0.00001 | yes |
| 3-Hydroxybutyrate | 0.018 | 0.019 | 0.40 | 0.047 | 0.061 | 0.48 | 0.045 | 0.032 | 0.19 | no |
| Acetate | -0.044 | 0.015 | 0.005 | 0.131 | 0.070 | 0.08 | 0.491 | 0.026 | < 0.00001 | yes |
| Acetoacetate | -0.007 | 0.018 | 0.74 | 0.017 | 0.057 | 0.79 | -0.008 | 0.027 | 0.79 | no |
| Acetone | -0.076 | 0.019 | 0.0002 | -0.025 | 0.053 | 0.68 | 0.186 | 0.023 | < 0.00001 | yes |
| Creatinine | 0.082 | 0.022 | 0.0003 | -0.007 | 0.063 | 0.92 | -0.259 | 0.032 | < 0.00001 | yes |
| Albumin | -0.028 | 0.010 | 0.009 | -0.043 | 0.012 | 0.0004 | -0.048 | 0.004 | < 0.00001 | yes |
| VLDL-TG cluster | 0.182 | 0.019 | < 0.00001 | 0.094 | 0.054 | 0.10 | -0.020 | 0.027 | 0.51 | yes |
| L-VLDL-TG cluster | 0.094 | 0.020 | < 0.00001 | -0.131 | 0.051 | 0.01 | -0.258 | 0.024 | < 0.00001 | yes |
| VLDL-C cluster | -0.072 | 0.018 | 0.0002 | -0.309 | 0.047 | < 0.00001 | -0.271 | 0.024 | < 0.00001 | yes |
| M-LDL-P cluster | -0.130 | 0.018 | < 0.00001 | -0.412 | 0.046 | < 0.00001 | -0.373 | 0.023 | < 0.00001 | yes |
| VLDL-P cluster | 0.019 | 0.019 | 0.37 | -0.192 | 0.051 | 0.0003 | -0.225 | 0.026 | < 0.00001 | yes |
| XS-VLDL-P cluster | -0.064 | 0.019 | 0.001 | -0.073 | 0.053 | 0.21 | 0.163 | 0.027 | < 0.00001 | yes |
| XS-VLDL-C cluster | -0.207 | 0.019 | < 0.00001 | -0.267 | 0.051 | < 0.00001 | 0.055 | 0.027 | 0.06 | yes |
| IDL-C cluster | -0.304 | 0.018 | < 0.00001 | -0.516 | 0.048 | < 0.00001 | -0.247 | 0.026 | < 0.00001 | yes |
| LDL-FC cluster | -0.059 | 0.004 | < 0.00001 | -0.071 | 0.007 | < 0.00001 | -0.066 | 0.004 | < 0.00001 | yes |
| M-LDL-C cluster | -0.070 | 0.007 | < 0.00001 | -0.095 | 0.009 | < 0.00001 | -0.093 | 0.005 | < 0.00001 | yes |
| XL-HDL-C cluster | -0.162 | 0.015 | < 0.00001 | -0.047 | 0.054 | 0.42 | 0.449 | 0.033 | < 0.00001 | yes |
| Triglycerides in very large HDL | 0.117 | 0.019 | < 0.00001 | 0.325 | 0.054 | < 0.00001 | 0.431 | 0.027 | < 0.00001 | yes |
| Triglycerides in large HDL | 0.071 | 0.020 | 0.0004 | 0.448 | 0.057 | < 0.00001 | 0.638 | 0.029 | < 0.00001 | yes |
| HDL-TG cluster | 0.131 | 0.019 | < 0.00001 | 0.301 | 0.055 | < 0.00001 | 0.324 | 0.028 | < 0.00001 | yes |
| S-HDL-C cluster | -0.147 | 0.008 | < 0.00001 | -0.190 | 0.012 | < 0.00001 | -0.217 | 0.007 | < 0.00001 | yes |

| | Univariate Cox regression results for MASLD[1] | | | Univariate Cox regression results for MASH[1] | | | Univariate Cox regression results for cirrhosis[1] | | | |
|---|---|---|---|---|---|---|---|---|---|
| **Metabolite** | **Est** | **Se** | **Pval-BH** | **Est** | **Se** | **Pval-BH** | **Est** | **Se** | **Pval-BH** | **Included for phase 1?[2]** |
| S-HDL-L cluster | -0.150 | 0.019 | < 0.00001 | -0.290 | 0.051 | < 0.00001 | -0.346 | 0.025 | < 0.00001 | yes |
| Free cholesterol in small HDL | -0.181 | 0.019 | < 0.00001 | -0.300 | 0.051 | < 0.00001 | -0.248 | 0.026 | < 0.00001 | yes |
| HDL-C cluster | -0.368 | 0.021 | < 0.00001 | -0.215 | 0.060 | 0.0005 | 0.148 | 0.030 | < 0.00001 | yes |
| HDL-P cluster | -0.320 | 0.019 | < 0.00001 | -0.381 | 0.052 | < 0.00001 | -0.218 | 0.028 | < 0.00001 | yes |
| Fischer's ratio | 0.062 | 0.020 | 0.003 | -0.364 | 0.044 | < 0.00001 | -0.507 | 0.012 | < 0.00001 | yes |

[1] Adjusted by age at blood collection, sex, fasting time, and menopause and use of exogenous hormones for women
[2] There was at least one metabolite-risk factor association with Benjamini-Hochberg adjusted p-value < 0.05
**Abbreviations:** Est, Estimate; MASH, Metabolic dysfunction-associated steatohepatitis; MASLD, Metabolic dysfunction-associated steatotic liver disease; Pval-BH, Benjamini-Hochberg adjusted P-value; Se, Standard error;

**Supplementary Table S7.** Variables Robustly Associated with Gallstones in a Cox Model with Data Shared Lasso Selection among Gallstones-related Clinical Conditions (Gallstone, Cholecystitis and Cholecystectomy) and Bootstrap Resampling.

| Variable | Bootstrap Selection (%) | Bootstrap Median HR | HR 95% Range (Selected Bootstraps) | Final HR (95% CI) |
|---|---|---|---|---|
| **Unpenalized adjustment variables** | | | | |
| Female Sex and no current hormone use or menopause | 100 (fixed) | **1.83** | 1.67-2.00 | 1.84 (1.69-2.01) |
| Female Sex and current hormone use or menopause | 100 (fixed) | **1.73** | 1.64-1.84 | 1.75 (1.65-1.84) |
| Age at blood sampling | 100 (fixed) | **1.35** | 1.30-1.41 | 1.35 (1.29-1.41) |
| Fasting time | 100 (fixed) | **1.05** | 1.03-1.07 | 1.05 (1.03-1.07) |
| **Variables selected via Lasso penalty** | | | | |
| Sphingomyelins | 95 | **1.56** | 1.39-1.77 | 1.56 (1.44-1.70) |
| VLDL-TG cluster | 100 | **1.32** | 1.11-1.61 | 1.28 (1.18-1.38) |
| Free cholesterol in small HDL | 90 | **0.79** | 0.58-0.96 | 0.68 (0.58-0.80) |
| HDL-C cluster | 99 | 0.81 | 0.63-1.06 | 0.79 (0.73-0.86) |
| S-HDL-L cluster | 61 | **1.22** | 1.12-2.04 | 1.24 (1.11-1.39) |
| ApoB cluster | 95 | 0.83 | 0.57-1.03 | 0.91 (0.82-1.00) |
| Omega-3 fatty acids | 100 | **0.86** | 0.80-0.92 | 0.86 (0.84-0.88) |
| XS-VLDL-P cluster | 88 | **0.87** | 0.58-0.97 | 0.86 (0.79-0.92) |
| SFA cluster | 85 | **0.87** | 0.68-0.94 | 0.84 (0.78-0.90) |
| LDL-TG cluster | 58 | **1.07** | 1.01-1.37 | 1.17 (1.06-1.29) |
| Triglycerides in large HDL | 83 | **1.06** | 1.01-1.31 | 1.06 (1.00-1.11) |
| Tyrosine | 99 | **0.95** | 0.90-0.98 | 0.95 (0.93-0.97) |
| LDL_C cluster | 62 | **0.95** | 0.44-0.99 | 0.94 (0.89-0.99) |
| Glucose | 96 | **1.04** | 1.02-1.06 | 1.04 (1.01-1.06) |
| XL-HDL-C cluster | 62 | 1.03 | 0.99-1.14 | 1.04 (0.99-1.09) |
| Albumin | 99 | **0.97** | 0.94-0.98 | 0.97 (0.96-0.98) |
| Acetone | 82 | **0.98** | 0.95-0.99 | 0.97 (0.95-1.00) |
| Alanine | 60 | **1.02** | 1.01-1.05 | 1.02 (1.00-1.05) |
| Lactate | 72 | **1.02** | 1.01-1.05 | 1.01 (0.99-1.04) |
| Creatinine | 64 | 1.02 | 0.99-1.05 | 1.02 (0.99-1.04) |
| Docosahexaenoic acid | 55 | 1.00 | 0.93-1.08 | |
| Phosphatidylc cluster | 55 | 1.00 | 0.70-1.12 | |
| Valine cluster | 51 | 1.00 | 0.97-1.07 | |
| Average diameter for LDL particles | 51 | 1.00 | 0.98-1.06 | |
| HDL-P cluster | 46 | 1.00 | 0.62-1.14 | |
| Fischer ratio | 42 | 1.00 | 0.94-1.03 | |
| Acetate | 41 | 1.00 | 0.97-1.02 | |
| LDL-FC cluster | 40 | 1.00 | 0.95-1.63 | |
| Degree of unsaturation | 40 | 1.00 | 0.91-1.07 | |
| L-HDL-C cluster | 39 | 1.00 | 0.89-1.08 | |
| 3-Hydroxybutyrate | 36 | 1.00 | 0.97-1.03 | |
| Histidine | 35 | 1.00 | 0.97-1.03 | |
| M-LDL-C cluster | 35 | 1.00 | 0.87-1.41 | |
| XXL-VLDL-TG cluster | 35 | 1.00 | 0.90-1.05 | |
| VLDL-C cluster | 34 | 1.00 | 1.16-2.26 | |
| Citrate | 32 | 1.00 | 0.97-1.04 | |
| Cluster 8 (ApoA1 cluster | 32 | 1.00 | 0.87-1.40 | |
| S-HDL-C cluster | 31 | 1.00 | 0.66-1.14 | |
| Omega-6 cluster | 27 | 1.00 | 0.90-1.21 | |
| HDL-TG cluster | 27 | 1.00 | 0.67-1.05 | |

| Variable | Bootstrap Selection (%) | Bootstrap Median HR | HR 95% Range (Selected Bootstraps) | Final HR (95% CI) |
|---|---|---|---|---|
| IDL-C cluster | 22 | 1.00 | 0.85-1.94 | |
| L-VLDL-TG cluster | 20 | 1.00 | 0.87-1.13 | |
| XS-VLDL-C cluster | 14 | 1.00 | 0.78-1.50 | |
| M-LDL-P cluster | 11 | 1.00 | 0.78-1.46 | |
| VLDL-P cluster | 3 | 1.00 | 0.48-1.15 | |

**Bootstrap Selection (%):** Proportion of 1,000 bootstrap samples in which the variable was selected using data shared Lasso-penalized Cox regression ($\lambda = \lambda_{min}$ selected via 5-fold cross-validation). For unpenalized adjustment variables, selection frequency is reported as 100 (fixed) to indicate forced inclusion.
**Bootstrap Median HR:** Median hazard ratio across all bootstraps, obtained from an unpenalized Cox regression refit using only the variables selected in each bootstrap. For penalized variables, HR = 1 was imputed when the variable was not selected. For unpenalized variables, the HR was calculated in every sample. **Bold** indicates median HRs with consistent directionality and bootstrap support (selected HR 95% percentile range excludes 1), not formal statistical significance.
**HR 95% Range (Selected Bootstraps):** 2.5th and 97.5th percentiles of HRs from unpenalized Cox refits in bootstrap samples where the variable was selected. For unpenalized variables, this reflects all bootstraps.
**Final HR (95%CI):** Variables with a bootstrap median hazard ratio ≠ 1 were included in the final model, which was refit using unpenalized Cox regression. Confidence intervals are shown for completeness but may underestimate uncertainty for penalized variables, as they do not account for variability due to selection.
**Abbreviations:** CI, confidence interval; HR, Hazard Ratio;

**Supplementary Table S8.** Variables Robustly Associated with Cholecystitis in a Cox Model with Data Shared Lasso Selection among Gallstones-related Clinical Conditions (Gallstone, Cholecystitis and Cholecystectomy) and Bootstrap Resampling.

| Variable | Bootstrap Selection (%) | Bootstrap Median HR | HR 95% Range (Selected Bootstraps) | Final HR (95% CI) |
|---|---|---|---|---|
| **Unpenalized adjustment variables** | | | | |
| Female Sex and no current hormone use or menopause | 100 (fixed) | **1.88** | 1.66-2.13 | 1.89 (1.69-2.12) |
| Female Sex and current hormone use or menopause | 100 (fixed) | **1.67** | 1.54-1.82 | 1.68 (1.57-1.81) |
| Age at blood sampling | 100 (fixed) | **1.29** | 1.22-1.36 | 1.28 (1.21-1.36) |
| Fasting time | 100 (fixed) | **1.04** | 1.01-1.07 | 1.04 (1.01-1.07) |
| **Variables selected via Lasso penalty** | | | | |
| Sphingomyelins | 95 | **1.51** | 1.32-1.77 | 1.48 (1.33-1.66) |
| VLDL-TG cluster | 100 | **1.35** | 1.10-1.78 | 1.35 (1.23-1.48) |
| Free cholesterol in small HDL | 90 | **0.80** | 0.54-0.99 | 0.77 (0.65-0.90) |
| HDL-C cluster | 99 | 0.81 | 0.64-1.33 | 0.75 (0.66-0.84) |
| XS-VLDL-P cluster | 90 | **0.82** | 0.46-0.94 | 0.84 (0.77-0.92) |
| S-HDL-L cluster | 62 | **1.21** | 1.07-2.25 | 1.18 (1.04-1.33) |
| SFA cluster | 85 | **0.86** | 0.59-0.93 | 0.87 (0.81-0.93) |
| ApoB cluster | 84 | 0.87 | 0.50-1.07 | 0.90 (0.81-1.00) |
| Omega-3 fatty acids | 100 | **0.89** | 0.83-1.00 | 0.88 (0.85-0.91) |
| Triglycerides in large HDL | 79 | **1.11** | 1.04-1.50 | 1.10 (1.02-1.18) |
| Tyrosine | 99 | **0.93** | 0.85-0.97 | 0.93 (0.90-0.96) |
| XL-HDL-C cluster | 63 | **1.07** | 1.03-1.26 | 1.07 (0.99-1.15) |
| Acetone | 92 | **0.96** | 0.92-0.98 | 0.95 (0.92-0.98) |
| Histidine | 73 | **1.03** | 1.01-1.06 | 1.03 (1.00-1.06) |
| Albumin | 71 | **0.98** | 0.94-0.99 | 0.98 (0.95-1.00) |
| Acetate | 63 | **0.98** | 0.96-0.99 | 0.98 (0.96-1.01) |
| Lactate | 69 | **1.02** | 1.00-1.06 | 1.02 (0.99-1.05) |
| Valine cluster | 53 | 1.01 | 0.98-1.14 | 1.02 (0.98-1.06) |
| Docosahexaenoic acid | 56 | 1.00 | 0.90-1.04 | |
| Phosphatidylc cluster | 54 | 1.00 | 0.75-1.31 | |
| LDL_C cluster | 52 | 1.00 | 0.50-1.17 | |
| Average diameter for LDL particles | 48 | 1.00 | 0.95-1.07 | |
| Alanine | 48 | 1.00 | 0.97-1.06 | |
| 3-Hydroxybutyrate | 47 | 1.00 | 0.95-1.03 | |
| Glucose | 46 | 1.00 | 0.95-1.03 | |
| L-HDL-C cluster | 46 | 1.00 | 0.81-1.01 | |
| Creatinine | 40 | 1.00 | 0.95-1.04 | |
| Fischer ratio | 39 | 1.00 | 0.90-1.03 | |
| LDL-FC cluster | 38 | 1.00 | 0.89-1.90 | |
| M-LDL-C cluster | 38 | 1.00 | 0.81-1.34 | |
| Degree of unsaturation | 35 | 1.00 | 0.89-1.10 | |
| VLDL-C cluster | 33 | 1.00 | 1.11-2.67 | |
| Citrate | 32 | 1.00 | 0.97-1.05 | |
| XXL-VLDL-TG cluster | 31 | 1.00 | 0.89-1.10 | |
| Cluster 8 (ApoA1 cluster | 31 | 1.00 | 0.79-1.38 | |
| HDL-P cluster | 27 | 1.00 | 0.43-1.05 | |
| Omega-6 cluster | 27 | 1.00 | 0.90-1.29 | |
| HDL-TG cluster | 26 | 1.00 | 0.60-1.02 | |
| LDL-TG cluster | 24 | 1.00 | 0.92-1.56 | |

| Variable | Bootstrap Selection (%) | Bootstrap Median HR | HR 95% Range (Selected Bootstraps) | Final HR (95% CI) |
|---|---|---|---|---|
| L-VLDL-TG cluster | 21 | 1.00 | 0.81-1.11 | |
| S-HDL-C cluster | 17 | 1.00 | 0.62-1.22 | |
| M-LDL-P cluster | 15 | 1.00 | 0.48-1.15 | |
| XS-VLDL-C cluster | 14 | 1.00 | 0.81-1.79 | |
| IDL-C cluster | 11 | 1.00 | 0.87-2.15 | |
| VLDL-P cluster | 2 | 1.00 | 0.57-1.85 | |

**Bootstrap Selection (%):** Proportion of 1,000 bootstrap samples in which the variable was selected using data shared Lasso-penalized Cox regression ($\lambda = \lambda_{min}$ selected via 5-fold cross-validation). For unpenalized adjustment variables, selection frequency is reported as 100 (fixed) to indicate forced inclusion.
**Bootstrap Median HR:** Median hazard ratio across all bootstraps, obtained from an unpenalized Cox regression refit using only the variables selected in each bootstrap. For penalized variables, HR = 1 was imputed when the variable was not selected. For unpenalized variables, the HR was calculated in every sample. **Bold** indicates median HRs with consistent directionality and bootstrap support (selected HR 95% percentile range excludes 1), not formal statistical significance.
**HR 95% Range (Selected Bootstraps):** 2.5th and 97.5th percentiles of HRs from unpenalized Cox refits in bootstrap samples where the variable was selected. For unpenalized variables, this reflects all bootstraps.
**Final HR (95%CI):** Variables with a bootstrap median hazard ratio ≠ 1 were included in the final model, which was refit using unpenalized Cox regression. Confidence intervals are shown for completeness but may underestimate uncertainty for penalized variables, as they do not account for variability due to selection.
**Abbreviations:** CI, confidence interval; HR, Hazard Ratio;

**Supplementary Table S9.** Variables Robustly Associated with Cholecystectomy in a Cox Model with Data Shared Lasso Selection among Gallstones-related Clinical Conditions (Gallstone, Cholecystitis and Cholecystectomy) and Bootstrap Resampling.

| Variable | Bootstrap Selection (%) | Bootstrap Median HR | HR 95% Range (Selected Bootstraps) | Final HR (95% CI) |
|---|---|---|---|---|
| **Unpenalized adjustment variables** | | | | |
| Female Sex and no current hormone use or menopause | 100 (fixed) | **2.35** | 2.13-2.59 | 2.35 (2.13-2.59) |
| Female Sex and current hormone use or menopause | 100 (fixed) | **2.27** | 2.12-2.45 | 2.28 (2.14-2.44) |
| Age at blood sampling | 100 (fixed) | **1.59** | 1.52-1.66 | 1.60 (1.51-1.68) |
| Fasting time | 100 (fixed) | **1.03** | 1.01-1.06 | 1.03 (1.01-1.06) |
| **Variables selected via Lasso penalty** | | | | |
| Sphingomyelins | 95 | **1.54** | 1.33-1.79 | 1.51 (1.37-1.68) |
| Free cholesterol in small HDL | 90 | **0.67** | 0.47-0.88 | 0.66 (0.56-0.78) |
| S-HDL-L cluster | 64 | **1.36** | 1.15-2.35 | 1.34 (1.19-1.51) |
| VLDL-TG cluster | 100 | **1.35** | 1.09-1.81 | 1.35 (1.20-1.51) |
| XS-VLDL-P cluster | 89 | **0.80** | 0.43-0.93 | 0.82 (0.75-0.89) |
| HDL-C cluster | 99 | 0.81 | 0.55-1.06 | 0.76 (0.68-0.85) |
| SFA cluster | 85 | **0.87** | 0.61-0.94 | 0.87 (0.82-0.93) |
| ApoB cluster | 84 | 0.88 | 0.56-1.10 | 0.96 (0.86-1.07) |
| Omega-3 fatty acids | 100 | **0.90** | 0.85-0.99 | 0.90 (0.84-0.97) |
| XL-HDL-C cluster | 69 | **1.10** | 1.05-1.24 | 1.09 (1.02-1.17) |
| Triglycerides in large HDL | 63 | **1.08** | 1.02-1.43 | 1.06 (0.99-1.13) |
| Tyrosine | 99 | **0.92** | 0.88-0.97 | 0.92 (0.89-0.95) |
| Valine cluster | 81 | **1.06** | 1.02-1.13 | 1.06 (1.02-1.10) |
| Acetone | 95 | **0.95** | 0.92-0.98 | 0.95 (0.92-0.98) |
| Citrate | 68 | **0.98** | 0.95-0.99 | 0.98 (0.95-1.00) |
| L-VLDL-TG cluster | 73 | 1.02 | 0.93-1.17 | 1.02 (0.93-1.12) |
| Histidine | 75 | **1.02** | 1.01-1.05 | 1.02 (1.00-1.05) |
| 3-Hydroxybutyrate | 65 | **0.98** | 0.95-0.99 | 0.98 (0.95-1.01) |
| Lactate | 69 | **1.02** | 1.01-1.05 | 1.02 (1.00-1.05) |
| Docosahexaenoic acid | 56 | 0.99 | 0.90-1.04 | 0.99 (0.92-1.05) |
| S-HDL-C cluster | 66 | 1.00 | 0.74-1.34 | |
| Fischer ratio | 61 | 1.00 | 0.94-1.06 | |
| Phosphatidylc cluster | 55 | 1.00 | 0.69-1.15 | |
| LDL_C cluster | 52 | 1.00 | 0.44-1.08 | |
| Albumin | 51 | 1.00 | 0.96-1.02 | |
| Alanine | 46 | 1.00 | 0.97-1.05 | |
| Average diameter for LDL particles | 45 | 1.00 | 0.97-1.07 | |
| LDL-TG cluster | 45 | 1.00 | 0.92-1.50 | |
| LDL-FC cluster | 42 | 1.00 | 0.89-1.37 | |
| VLDL-C cluster | 40 | 1.00 | 1.11-2.23 | |
| Acetate | 39 | 1.00 | 0.97-1.02 | |
| L-HDL-C cluster | 39 | 1.00 | 0.85-1.11 | |
| M-LDL-C cluster | 38 | 1.00 | 0.92-1.47 | |
| Degree of unsaturation | 38 | 1.00 | 0.90-1.06 | |
| Creatinine | 35 | 1.00 | 0.96-1.04 | |
| XXL-VLDL-TG cluster | 35 | 1.00 | 0.87-1.07 | |
| Omega-6 cluster | 30 | 1.00 | 0.93-1.24 | |
| Cluster 8 (ApoA1 cluster | 29 | 1.00 | 0.82-1.76 | |
| Glucose | 28 | 1.00 | 0.96-1.04 | |

| Variable | Bootstrap Selection (%) | Bootstrap Median HR | HR 95% Range (Selected Bootstraps) | Final HR (95% CI) |
|---|---|---|---|---|
| HDL-TG cluster | 28 | 1.00 | 0.61-1.05 | |
| HDL-P cluster | 26 | 1.00 | 0.61-1.25 | |
| IDL-C cluster | 19 | 1.00 | 0.93-2.16 | |
| XS-VLDL-C cluster | 15 | 1.00 | 0.97-1.84 | |
| M-LDL-P cluster | 10 | 1.00 | 0.63-1.23 | |
| VLDL-P cluster | 3 | 1.00 | 0.55-1.63 | |

**Bootstrap Selection (%):** Proportion of 1,000 bootstrap samples in which the variable was selected using data shared Lasso-penalized Cox regression ($\lambda = \lambda_{min}$ selected via 5-fold cross-validation). For unpenalized adjustment variables, selection frequency is reported as 100 (fixed) to indicate forced inclusion.
**Bootstrap Median HR:** Median hazard ratio across all bootstraps, obtained from an unpenalized Cox regression refit using only the variables selected in each bootstrap. For penalized variables, HR = 1 was imputed when the variable was not selected. For unpenalized variables, the HR was calculated in every sample. **Bold** indicates median HRs with consistent directionality and bootstrap support (selected HR 95% percentile range excludes 1), not formal statistical significance.
**HR 95% Range (Selected Bootstraps):** 2.5th and 97.5th percentiles of HRs from unpenalized Cox refits in bootstrap samples where the variable was selected. For unpenalized variables, this reflects all bootstraps.
**Final HR (95%CI):** Variables with a bootstrap median hazard ratio ≠ 1 were included in the final model, which was refit using unpenalized Cox regression. Confidence intervals are shown for completeness but may underestimate uncertainty for penalized variables, as they do not account for variability due to selection.
**Abbreviations:** CI, confidence interval; HR, Hazard Ratio;

**Supplementary Table S10.** Variables Robustly Associated with Primary Sclerosing Cholangitis (PSC) in a Cox Model with Lasso Selection and Bootstrap Resampling.

| Variable | Bootstrap Selection (%) | Bootstrap Median HR | HR 95% Range (Selected Bootstraps) | Final HR (95% CI) |
|---|---|---|---|---|
| **Unpenalized adjustment variables** | | | | |
| Female Sex and no current hormone use or menopause | 100 (fixed) | **0.67** | 0.44-0.97 | 0.67 (0.46-0.98) |
| Female Sex and current hormone use or menopause | 100 (fixed) | **0.69** | 0.57-0.84 | 0.67 (0.57-0.78) |
| Age at blood sampling | 100 (fixed) | 0.98 | 0.86-1.10 | 0.98 (0.85-1.13) |
| Fasting time | 100 (fixed) | **1.08** | 1.02-1.15 | 1.08 (1.02-1.15) |
| **Variables selected via Lasso penalty** | | | | |
| Sphingomyelins | 64 | **2.01** | 1.56-2.91 | 1.83 (1.53-2.19) |
| IDL-C cluster | 84 | 0.60 | 0.29-1.04 | 0.57 (0.49-0.66) |
| S-HDL-C cluster | 99 | **0.81** | 0.31-0.92 | 0.78 (0.74-0.82) |
| HDL-TG cluster | 85 | 1.15 | 0.89-1.67 | 1.17 (1.09-1.25) |
| Alanine | 81 | **1.11** | 1.05-1.21 | 1.10 (1.03-1.18) |
| XL-HDL-C cluster | 75 | **0.91** | 0.79-0.98 | 0.89 (0.83-0.95) |
| Fischer ratio | 88 | **0.91** | 0.83-0.98 | 0.91 (0.85-0.99) |
| Valine cluster | 70 | **0.93** | 0.83-0.97 | 0.91 (0.84-0.99) |
| Glycine | 68 | **0.96** | 0.91-0.98 | 0.95 (0.91-0.99) |
| Docosahexaenoic acid | 55 | 0.96 | 0.77-1.15 | 0.88 (0.82-0.94) |
| Albumin | 85 | **0.96** | 0.86-0.98 | 0.96 (0.94-0.98) |
| Degree of unsaturation | 55 | 1.00 | 0.84-1.16 | |
| ApoB cluster | 50 | 1.00 | 0.23-1.17 | |
| Histidine | 42 | 1.00 | 0.90-1.07 | |
| XS-VLDL-P cluster | 42 | 1.00 | 1.04-3.45 | |
| Average diameter for LDL particles | 41 | 1.00 | 0.89-1.07 | |
| Omega-6 cluster | 39 | 1.00 | 0.72-1.16 | |
| Omega-3 fatty acids | 38 | 1.00 | 0.73-1.22 | |
| L-HDL-C cluster | 25 | 1.00 | 0.90-1.44 | |
| LDL-FC cluster | 24 | 1.00 | 0.98-4.37 | |
| HDL-C cluster | 24 | 1.00 | 0.35-1.07 | |
| Triglycerides in large HDL | 24 | 1.00 | 0.66-1.25 | |
| Free cholesterol in small HDL | 22 | 1.00 | 0.36-1.13 | |
| XS-VLDL-C cluster | 19 | 1.00 | 0.24-1.12 | |
| SFA cluster | 16 | 1.00 | 0.64-1.36 | |
| S-HDL-L cluster | 16 | 1.00 | 0.82-4.92 | |
| VLDL-C cluster | 15 | 1.00 | 0.90-4.37 | |
| LDL_C cluster | 14 | 1.00 | 0.68-2.76 | |
| HDL-P cluster | 14 | 1.00 | 0.60-3.59 | |
| Cluster 8 (ApoA1 cluster | 13 | 1.00 | 0.86-1.71 | |
| M-LDL-C cluster | 9 | 1.00 | 0.33-1.17 | |
| VLDL-P cluster | 7 | 1.00 | 0.26-1.63 | |
| Phosphatidylc cluster | 7 | 1.00 | 0.48-2.18 | |
| M-LDL-P cluster | 3 | 1.00 | 0.33-5.22 | |

**Bootstrap Selection (%):** Proportion of 1,000 bootstrap samples in which the variable was selected using Lasso-penalized Cox regression ($\lambda = \lambda_{min}$ selected via 5-fold cross-validation). For unpenalized adjustment variables, selection frequency is reported as 100 (fixed) to indicate forced inclusion.

**Bootstrap Median HR:** Median hazard ratio across all bootstraps, obtained from an unpenalized Cox regression refit using only the variables selected in each bootstrap. For penalized variables, HR = 1 was imputed when the variable was not selected. For unpenalized variables, the HR was calculated in every sample. **Bold** indicates median HRs with consistent directionality and bootstrap support (selected HR 95% percentile range excludes 1), not formal statistical significance.
**HR 95% Range (Selected Bootstraps):** 2.5th and 97.5th percentiles of HRs from unpenalized Cox refits in bootstrap samples where the variable was selected. For unpenalized variables, this reflects all bootstraps.
**Final HR (95%CI):** Variables with a bootstrap median hazard ratio ≠ 1 were included in the final model, which was refit using unpenalized Cox regression. Confidence intervals are shown for completeness but may underestimate uncertainty for penalized variables, as they do not account for variability due to selection.
CI, confidence interval; HR, Hazard Ratio;

**Supplementary Table S11.** Variables Robustly Associated with MASLD in a Cox Model with Data Shared Lasso Selection among Spectrum of Metabolic Liver Disease (MASLD, MASH, Cirrhosis) and Bootstrap Resampling.

| Variable | Bootstrap Selection (%) | Bootstrap Median HR | HR 95% Range (Selected Bootstraps) | Final HR (95% CI) |
|---|---|---|---|---|
| **Unpenalized adjustment variables** | | | | |
| Female Sex and current hormone use or menopause | 100 (fixed) | **1.76** | 1.54-1.97 | 1.74 (1.57-1.93) |
| Female Sex and no current hormone use or menopause | 100 (fixed) | **1.20** | 1.01-1.44 | 1.20 (1.02-1.41) |
| Age at blood sampling | 100 (fixed) | **0.45** | 0.42-0.49 | 0.45 (0.42-0.49) |
| Fasting time | 100 (fixed) | **1.11** | 1.07-1.14 | 1.11 (1.07-1.14) |
| **Variables selected via Lasso penalty** | | | | |
| HDL-C cluster | 58 | **0.66** | 0.26-0.79 | 0.58 (0.48-0.71) |
| ApoB cluster | 98 | **0.66** | 0.22-0.99 | 0.71 (0.61-0.83) |
| HDL-TG cluster | 60 | **0.66** | 0.45-0.78 | 0.58 (0.46-0.73) |
| LDL-TG cluster | 90 | 1.26 | 0.95-1.73 | 1.28 (1.08-1.52) |
| IDL-C cluster | 95 | 0.80 | 0.52-1.23 | 0.78 (0.63-0.97) |
| VLDL-TG cluster | 83 | 1.24 | 0.91-1.79 | 1.39 (1.07-1.81) |
| S-HDL-L cluster | 70 | 0.82 | 0.49-1.14 | 0.98 (0.88-1.10) |
| Omega-3 fatty acids | 98 | **0.82** | 0.72-0.97 | 0.78 (0.70-0.87) |
| XS-VLDL-P cluster | 98 | 1.17 | 0.88-1.87 | 1.11 (0.96-1.28) |
| Sphingomyelins | 69 | **1.17** | 1.02-1.50 | 1.24 (1.05-1.45) |
| Triglycerides in large HDL | 100 | 1.14 | 0.83-1.59 | 1.34 (1.13-1.60) |
| Alanine | 98 | **1.13** | 1.07-1.19 | 1.12 (1.07-1.18) |
| Glucose | 100 | **1.10** | 1.06-1.15 | 1.11 (1.07-1.15) |
| Citrate | 99 | **1.08** | 1.03-1.13 | 1.07 (1.03-1.12) |
| Glutamine | 99 | **0.94** | 0.90-0.98 | 0.94 (0.90-0.97) |
| Phosphatidylc cluster | 65 | 1.06 | 0.75-1.83 | 1.29 (0.99-1.69) |
| L-HDL-C cluster | 75 | 0.95 | 0.83-1.07 | 1.03 (0.94-1.12) |
| Average diameter for LDL particles | 87 | 0.96 | 0.89-1.06 | 0.95 (0.90-0.99) |
| Docosahexaenoic acid | 96 | 1.04 | 0.96-1.14 | 1.07 (0.99-1.17) |
| Glycine | 91 | **0.96** | 0.94-0.99 | 0.96 (0.93-0.99) |
| Albumin | 63 | **1.03** | 1.01-1.10 | 1.03 (0.99-1.08) |
| Creatinine | 98 | 0.97 | 0.92-1.02 | 0.98 (0.93-1.02) |
| Acetate | 99 | 0.97 | 0.94-1.01 | 0.97 (0.94-1.00) |
| Acetone | 98 | 0.97 | 0.92-1.03 | 0.97 (0.92-1.01) |
| Fischer ratio | 100 | 1.02 | 0.80-1.12 | 1.04 (0.99-1.08) |
| XXL-VLDL-TG cluster | 62 | 0.98 | 0.81-1.05 | 0.91 (0.82-1.01) |
| Lactate | 66 | 1.02 | 0.99-1.09 | 1.04 (1.00-1.09) |
| Tyrosine | 98 | 1.01 | 0.85-1.10 | 1.04 (0.99-1.08) |
| Degree of unsaturation | 100 | 1.01 | 0.83-1.13 | 1.04 (0.95-1.15) |
| Omega-6 cluster | 93 | 0.99 | 0.78-1.24 | 0.89 (0.73-1.08) |
| LDL-FC cluster | 92 | 1.01 | 0.88-1.90 | 1.02 (0.97-1.07) |
| S-HDL-C cluster | 78 | 1.00 | 0.64-1.20 | |
| Phenylalanine | 71 | 1.00 | 0.87-1.09 | |
| L-VLDL-TG cluster | 66 | 1.00 | 0.79-1.23 | |
| XL-HDL-C cluster | 55 | 1.00 | 0.93-1.22 | |
| Valine cluster | 53 | 1.00 | 0.89-1.48 | |
| LDL_C cluster | 51 | 1.00 | 0.71-1.70 | |
| Triglycerides in very large HDL | 51 | 1.00 | 0.71-1.24 | |
| Histidine | 49 | 1.00 | 0.91-0.99 | |
| Free cholesterol in small HDL | 49 | 1.00 | 1.01-2.39 | |

| Variable | Bootstrap Selection (%) | Bootstrap Median HR | HR 95% Range (Selected Bootstraps) | Final HR (95% CI) |
|---|---|---|---|---|
| M-LDL-C cluster | 47 | 1.00 | 0.74-1.73 | |
| Cluster 8 (ApoA1 cluster | 41 | 1.00 | 0.91-2.15 | |
| VLDL-P cluster | 34 | 1.00 | 0.57-1.91 | |
| HDL-P cluster | 31 | 1.00 | 0.44-1.40 | |
| M-LDL-P cluster | 23 | 1.00 | 0.79-2.76 | |
| XS-VLDL-C cluster | 21 | 1.00 | 0.57-1.62 | |
| VLDL-C cluster | 16 | 1.00 | 0.52-1.26 | |

**Bootstrap Selection (%):** Proportion of 1,000 bootstrap samples in which the variable was selected using data shared Lasso-penalized Cox regression ($\lambda = \lambda_{min}$ selected via 5-fold cross-validation). For unpenalized adjustment variables, selection frequency is reported as 100 (fixed) to indicate forced inclusion.
**Bootstrap Median HR:** Median hazard ratio across all bootstraps, obtained from an unpenalized Cox regression refit using only the variables selected in each bootstrap. For penalized variables, HR = 1 was imputed when the variable was not selected. For unpenalized variables, the HR was calculated in every sample. **Bold** indicates median HRs with consistent directionality and bootstrap support (selected HR 95% percentile range excludes 1), not formal statistical significance.
**HR 95% Range (Selected Bootstraps):** 2.5th and 97.5th percentiles of HRs from unpenalized Cox refits in bootstrap samples where the variable was selected. For unpenalized variables, this reflects all bootstraps.
**Final HR (95% CI):** Variables with a bootstrap median hazard ratio ≠ 1 were included in the final model, which was refit using unpenalized Cox regression. Confidence intervals are shown for completeness but may underestimate uncertainty for penalized variables, as they do not account for variability due to selection.
**Abbreviations:** CI, confidence interval; HR, Hazard Ratio; MASH, Metabolic dysfunction-associated steatohepatitis; MASLD, Metabolic dysfunction-associated steatotic liver disease;

**Supplementary Table S12.** Variables Robustly Associated with MASH in a Cox Model with Data Shared Lasso Selection among Spectrum of Metabolic Liver Disease (MASLD, MASH, Cirrhosis) and Bootstrap Resampling.

| Variable | Bootstrap Selection (%) | Bootstrap Median HR | HR 95% Range (Selected Bootstraps) | Final HR (95% CI) |
|---|---|---|---|---|
| **Unpenalized adjustment variables** | | | | |
| Female Sex and no current hormone use or menopause | 100 (fixed) | 0.67 | 0.37-1.14 | 0.66 (0.40-1.12) |
| Female Sex and current hormone use or menopause | 100 (fixed) | 1.15 | 0.84-1.61 | 1.15 (0.87-1.53) |
| Age at blood sampling | 100 (fixed) | **0.57** | 0.47-0.69 | 0.57 (0.46-0.71) |
| Fasting time | 100 (fixed) | **1.26** | 1.18-1.34 | 1.27 (1.18-1.36) |
| **Variables selected via Lasso penalty** | | | | |
| XS-VLDL-P cluster | 87 | **2.72** | 1.20-7.44 | 3.00 (1.89-4.76) |
| IDL-C cluster | 94 | 0.40 | 0.18-1.03 | 0.33 (0.18-0.60) |
| HDL-C cluster | 98 | 0.49 | 0.17-1.46 | 0.41 (0.19-0.89) |
| S-HDL-L cluster | 81 | 0.56 | 0.24-1.12 | 0.52 (0.29-0.93) |
| Free cholesterol in small HDL | 73 | **1.63** | 1.02-3.49 | 2.03 (1.09-3.79) |
| HDL-TG cluster | 89 | 0.63 | 0.38-1.22 | 0.57 (0.32-1.00) |
| Triglycerides in large HDL | 86 | 1.55 | 0.96-2.74 | 1.55 (0.96-2.49) |
| Citrate | 99 | **1.42** | 1.27-1.60 | 1.42 (1.27-1.58) |
| ApoB cluster | 98 | 0.71 | 0.19-1.66 | 0.61 (0.33-1.12) |
| Glutamine | 97 | **0.72** | 0.65-0.80 | 0.71 (0.65-0.78) |
| Tyrosine | 72 | **1.32** | 1.04-1.69 | 1.29 (1.15-1.45) |
| Alanine | 99 | **1.31** | 1.15-1.52 | 1.33 (1.17-1.51) |
| VLDL-TG cluster | 59 | 0.80 | 0.31-1.06 | 0.69 (0.39-1.23) |
| Omega-6 cluster | 68 | 0.80 | 0.36-1.13 | 0.69 (0.40-1.21) |
| Glucose | 100 | **1.22** | 1.09-1.34 | 1.19 (1.10-1.29) |
| LDL-FC cluster | 65 | 1.21 | 0.97-4.75 | 1.44 (1.05-1.97) |
| Omega-3 fatty acids | 96 | 0.85 | 0.60-1.12 | 0.83 (0.61-1.11) |
| Sphingomyelins | 84 | 0.85 | 0.50-1.27 | 0.84 (0.55-1.28) |
| Cluster 8 (ApoA1 cluster | 64 | 1.17 | 0.99-2.85 | 1.38 (0.62-3.07) |
| Average diameter for LDL particles | 92 | 0.86 | 0.73-1.04 | 0.85 (0.75-0.95) |
| Degree of unsaturation | 72 | 0.87 | 0.61-1.09 | 0.86 (0.65-1.12) |
| Phosphatidylc cluster | 51 | **1.15** | 1.17-8.68 | 2.89 (1.18-7.08) |
| Lactate | 84 | **0.87** | 0.77-0.97 | 0.86 (0.77-0.97) |
| Glycine | 92 | 0.92 | 0.87-1.00 | 0.91 (0.86-0.97) |
| Histidine | 89 | 1.05 | 0.95-1.17 | 1.05 (0.94-1.17) |
| Creatinine | 69 | 0.95 | 0.79-1.04 | 0.93 (0.83-1.05) |
| Fischer ratio | 56 | 0.96 | 0.54-1.11 | 0.87 (0.78-0.97) |
| Albumin | 89 | 1.05 | 0.95-1.22 | 1.08 (0.96-1.21) |
| LDL-TG cluster | 97 | 1.03 | 0.56-1.88 | 0.84 (0.53-1.31) |
| Acetate | 76 | 1.03 | 0.95-1.24 | 1.06 (0.94-1.19) |
| Docosahexaenoic acid | 63 | 1.02 | 0.91-1.51 | 1.08 (0.87-1.34) |
| L-HDL-C cluster | 73 | 1.00 | 0.81-1.62 | |
| S-HDL-C cluster | 69 | 1.00 | 0.60-1.88 | |
| XXL-VLDL-TG cluster | 66 | 1.00 | 0.73-1.24 | |
| Acetone | 65 | 1.00 | 0.87-1.11 | |
| Valine cluster | 61 | 1.00 | 0.73-1.80 | |
| L-VLDL-TG cluster | 59 | 1.00 | 0.73-1.49 | |
| XL-HDL-C cluster | 53 | 1.00 | 0.81-1.13 | |
| HDL-P cluster | 52 | 1.00 | 0.58-3.81 | |
| M-LDL-C cluster | 49 | 1.00 | 0.22-1.79 | |

| Variable | Bootstrap Selection (%) | Bootstrap Median HR | HR 95% Range (Selected Bootstraps) | Final HR (95% CI) |
|---|---|---|---|---|
| Phenylalanine | 48 | 1.00 | 0.71-1.22 | |
| VLDL-C cluster | 39 | 1.00 | 0.34-2.29 | |
| LDL_C cluster | 34 | 1.00 | 0.29-3.06 | |
| Triglycerides in very large HDL | 33 | 1.00 | 0.58-1.86 | |
| XS-VLDL-C cluster | 31 | 1.00 | 0.32-3.11 | |
| M-LDL-P cluster | 20 | 1.00 | 1.32-7.91 | |
| VLDL-P cluster | 12 | 1.00 | 0.30-2.67 | |

**Bootstrap Selection (%):** Proportion of 1,000 bootstrap samples in which the variable was selected using data shared Lasso-penalized Cox regression ($\lambda = \lambda_{min}$ selected via 5-fold cross-validation). For unpenalized adjustment variables, selection frequency is reported as 100 (fixed) to indicate forced inclusion.
**Bootstrap Median HR:** Median hazard ratio across all bootstraps, obtained from an unpenalized Cox regression refit using only the variables selected in each bootstrap. For penalized variables, HR = 1 was imputed when the variable was not selected. For unpenalized variables, the HR was calculated in every sample. **Bold** indicates median HRs with consistent directionality and bootstrap support (selected HR 95% percentile range excludes 1), not formal statistical significance.
**HR 95% Range (Selected Bootstraps):** 2.5th and 97.5th percentiles of HRs from unpenalized Cox refits in bootstrap samples where the variable was selected. For unpenalized variables, this reflects all bootstraps.
**Final HR (95% CI):** Variables with a bootstrap median hazard ratio ≠ 1 were included in the final model, which was refit using unpenalized Cox regression. Confidence intervals are shown for completeness but may underestimate uncertainty for penalized variables, as they do not account for variability due to selection.
**Abbreviations:** CI, confidence interval; HR, Hazard Ratio; MASH, Metabolic dysfunction-associated steatohepatitis; MASLD, Metabolic dysfunction-associated steatotic liver disease;

**Supplementary Table S13.** Variables Robustly Associated with Cirrhosis in a Cox Model with Data Shared Lasso Selection among Spectrum of Metabolic Liver Disease (MASLD, MASH, Cirrhosis) and Bootstrap Resampling.

| Variable | Bootstrap Selection (%) | Bootstrap Median HR | HR 95% Range (Selected Bootstraps) | Final HR (95% CI) |
|---|---|---|---|---|
| **Unpenalized adjustment variables** | | | | |
| Female Sex and no current hormone use or menopause | 100 (fixed) | **0.19** | 0.13-0.32 | 0.18 (0.13-0.25) |
| Female Sex and current hormone use or menopause | 100 (fixed) | **0.44** | 0.36-0.68 | 0.44 (0.38-0.51) |
| Age at blood sampling | 100 (fixed) | **0.58** | 0.52-0.68 | 0.59 (0.52-0.66) |
| Fasting time | 100 (fixed) | **1.18** | 1.12-1.25 | 1.18 (1.14-1.22) |
| **Variables selected via Lasso penalty** | | | | |
| XS-VLDL-P cluster | 88 | **2.50** | 1.64-4.13 | 2.76 (2.18-3.49) |
| Triglycerides in large HDL | 97 | **1.97** | 1.21-3.73 | 2.16 (1.72-2.71) |
| IDL-C cluster | 95 | 0.54 | 0.27-1.58 | 0.40 (0.32-0.51) |
| Omega-3 fatty acids | 95 | **0.58** | 0.43-0.91 | 0.48 (0.42-0.55) |
| Phosphatidylc cluster | 52 | **1.68** | 1.64-5.91 | 3.74 (2.54-5.52) |
| HDL-TG cluster | 60 | **0.62** | 0.33-0.78 | 0.44 (0.35-0.55) |
| Docosahexaenoic acid | 66 | **1.54** | 1.38-2.05 | 1.66 (1.46-1.89) |
| Omega-6 cluster | 91 | 0.69 | 0.43-1.04 | 0.51 (0.41-0.63) |
| ApoB cluster | 98 | 0.71 | 0.21-1.06 | 0.65 (0.50-0.84) |
| Fischer ratio | 95 | **0.71** | 0.65-0.82 | 0.73 (0.70-0.77) |
| Glutamine | 98 | **0.73** | 0.68-0.78 | 0.72 (0.69-0.75) |
| HDL-C cluster | 57 | 0.73 | 0.21-1.05 | 0.60 (0.49-0.73) |
| Citrate | 99 | **1.26** | 1.15-1.39 | 1.26 (1.19-1.33) |
| Acetate | 58 | **1.24** | 1.20-1.45 | 1.32 (1.25-1.40) |
| LDL-FC cluster | 70 | **1.24** | 1.06-2.67 | 1.35 (1.10-1.65) |
| Degree of unsaturation | 88 | **0.81** | 0.63-1.00 | 0.90 (0.80-1.01) |
| Alanine | 98 | **1.19** | 1.02-1.34 | 1.19 (1.12-1.27) |
| Tyrosine | 98 | **1.17** | 1.08-1.35 | 1.15 (1.08-1.22) |
| LDL-TG cluster | 88 | 1.16 | 0.82-2.11 | 0.98 (0.83-1.15) |
| Creatinine | 76 | **0.87** | 0.76-0.93 | 0.86 (0.81-0.92) |
| Glucose | 100 | **1.13** | 1.06-1.20 | 1.11 (1.06-1.16) |
| Glycine | 92 | 0.96 | 0.92-1.00 | 0.95 (0.91-0.99) |
| Lactate | 74 | 0.97 | 0.89-1.02 | 0.97 (0.92-1.03) |
| Average diameter for LDL particles | 94 | 0.98 | 0.89-1.12 | 0.96 (0.90-1.02) |
| S-HDL-L cluster | 72 | 0.99 | 0.39-1.36 | 0.92 (0.81-1.04) |
| S-HDL-C cluster | 72 | 1.00 | 0.73-1.82 | |
| XXL-VLDL-TG cluster | 68 | 1.00 | 0.80-1.37 | |
| Histidine | 58 | 1.00 | 0.93-1.06 | |
| Phenylalanine | 49 | 1.00 | 0.84-1.03 | |
| Albumin | 49 | 1.00 | 0.93-1.06 | |
| Sphingomyelins | 46 | 1.00 | 1.03-1.85 | |
| L-VLDL-TG cluster | 45 | 1.00 | 0.65-1.25 | |
| XL-HDL-C cluster | 43 | 1.00 | 0.83-1.11 | |
| Triglycerides in very large HDL | 37 | 1.00 | 0.42-0.94 | |
| Acetone | 36 | 1.00 | 1.07-1.29 | |
| VLDL-TG cluster | 35 | 1.00 | 0.33-1.01 | |
| Cluster 8 (ApoA1 cluster | 35 | 1.00 | 0.89-2.12 | |
| L-HDL-C cluster | 30 | 1.00 | 0.92-1.74 | |
| M-LDL-C cluster | 29 | 1.00 | 0.45-1.28 | |
| LDL_C cluster | 28 | 1.00 | 0.32-1.43 | |

| Variable | Bootstrap Selection (%) | Bootstrap Median HR | HR 95% Range (Selected Bootstraps) | Final HR (95% CI) |
|---|---|---|---|---|
| M-LDL-P cluster | 21 | 1.00 | 0.83-4.42 | |
| HDL-P cluster | 20 | 1.00 | 0.59-3.65 | |
| Free cholesterol in small HDL | 19 | 1.00 | 0.39-1.21 | |
| Valine cluster | 11 | 1.00 | 0.64-1.30 | |
| XS-VLDL-C cluster | 11 | 1.00 | 0.42-2.29 | |
| VLDL-P cluster | 8 | 1.00 | 0.34-1.42 | |
| VLDL-C cluster | 8 | 1.00 | 0.58-2.66 | |

**Bootstrap Selection (%):** Proportion of 1,000 bootstrap samples in which the variable was selected using data shared Lasso-penalized Cox regression ($\lambda = \lambda_{min}$ selected via 5-fold cross-validation). For unpenalized adjustment variables, selection frequency is reported as 100 (fixed) to indicate forced inclusion.
**Bootstrap Median HR:** Median hazard ratio across all bootstraps, obtained from an unpenalized Cox regression refit using only the variables selected in each bootstrap. For penalized variables, HR = 1 was imputed when the variable was not selected. For unpenalized variables, the HR was calculated in every sample. **Bold** indicates median HRs with consistent directionality and bootstrap support (selected HR 95% percentile range excludes 1), not formal statistical significance.
**HR 95% Range (Selected Bootstraps):** 2.5th and 97.5th percentiles of HRs from unpenalized Cox refits in bootstrap samples where the variable was selected. For unpenalized variables, this reflects all bootstraps.
**Final HR (95% CI):** Variables with a bootstrap median hazard ratio ≠ 1 were included in the final model, which was refit using unpenalized Cox regression. Confidence intervals are shown for completeness but may underestimate uncertainty for penalized variables, as they do not account for variability due to selection.
**Abbreviations:** CI, confidence interval; HR, Hazard Ratio; MASH, Metabolic dysfunction-associated steatohepatitis; MASLD, Metabolic dysfunction-associated steatotic liver disease;

**Supplementary Table S14.** Variables Robustly Associated with Gallbladder Cancer in a Cox Model with Lasso Selection and Bootstrap Resampling.

| Variable | Bootstrap Selection (%) | Bootstrap Median HR | HR 95% Range (Selected Bootstraps) | Final HR (95% CI) |
|---|---|---|---|---|
| **Unpenalized adjustment variables** | | | | |
| Female sex | 100 (fixed) | **2.35** | 1.33-4.00 | 2.18 (1.41-3.37) |
| Age at blood sampling | 100 (fixed) | **1.67** | 1.22-2.38 | 1.64 (1.09-2.47) |
| Fasting time | 100 (fixed) | 1.20 | 0.97-1.36 | 1.20 (1.03-1.41) |
| **Unpenalized time-dependent covariates** | | | | |
| Gallstones | 100 (fixed) | **3.33** | 1.64-6.21 | 3.47 (2.00-6.04) |
| PSC | 100 (fixed) | **24.34** | 6.51-58.12 | 23.66 (9.61-58.23) |
| **Variables selected via Lasso penalty** | | | | |
| HDL-TG cluster | 92 | **1.46** | 1.04-3.33 | 1.57 (1.17-2.10) |
| XS-VLDL-P cluster | 63 | **0.81** | 0.29-0.98 | 0.74 (0.59-0.92) |
| S-HDL-L cluster | 71 | 1.11 | 0.61-3.36 | 1.07 (0.85-1.36) |
| Alanine | 62 | 1.00 | 0.81-1.36 | |
| Sphingomyelins | 46 | 1.00 | 0.41-1.56 | |
| HDL-C cluster | 33 | 1.00 | 0.58-1.67 | |
| VLDL-TG cluster | 32 | 1.00 | 0.49-1.94 | |
| IDL-C cluster | 28 | 1.00 | 0.72-5.17 | |
| S-HDL-C cluster | 27 | 1.00 | 0.83-2.79 | |
| Free cholesterol in small HDL | 18 | 1.00 | 0.12-0.99 | |

**Bootstrap Selection (%):** Proportion of 1,000 bootstrap samples in which the variable was selected using Lasso-penalized Cox regression ($\lambda = \lambda_{min}$ selected via 5-fold cross-validation). For unpenalized variables, selection frequency is reported as 100 (fixed) to indicate forced inclusion.
**Bootstrap Median HR:** Median hazard ratio across all bootstraps, obtained from an unpenalized Cox regression refit using only the variables selected in each bootstrap. For penalized variables, HR = 1 was imputed when the variable was not selected. For unpenalized variables, the HR was calculated in every sample. **Bold** indicates median HRs with consistent directionality and bootstrap support (selected HR 95% percentile range excludes 1), not formal statistical significance.
**HR 95% Range (Selected Bootstraps):** 2.5th and 97.5th percentiles of HRs from unpenalized Cox refits in bootstrap samples where the variable was selected. For unpenalized variables, this reflects all bootstraps.
**Final HR (95% CI):** Variables with a bootstrap median hazard ratio ≠ 1 were included in the final model, which was refit using unpenalized Cox regression. Confidence intervals are shown for completeness but may underestimate uncertainty for penalized variables, as they do not account for variability due to selection.
**Abbreviations:** CI, confidence interval; HR, Hazard Ratio; PSC, primary sclerosing cholangitis;

**Supplementary Table S15.** Variables Robustly Associated with Hepatocellular Carcinoma in a Cox Model with Lasso Selection and Bootstrap Resampling.

| Variable | Bootstrap Selection (%) | Bootstrap Median HR | HR 95% Range (Selected Bootstraps) | Final HR (95% CI) |
|---|---|---|---|---|
| **Unpenalized adjustment variables** | | | | |
| Female sex | 100 (fixed) | **0.21** | 0.15-0.30 | 0.22 (0.15-0.31) |
| Age at blood sampling | 100 (fixed) | 1.07 | 0.83-1.38 | 1.08 (0.82-1.44) |
| Fasting time | 100 (fixed) | 1.02 | 0.89-1.16 | 1.02 (0.91-1.15) |
| **Unpenalized time-dependent covariates** | | | | |
| MASH | 100 (fixed) | **2.40** | 1.08-4.75 | 2.49 (1.30-4.77) |
| MASLD | 100 (fixed) | 2.06 | 0.92-4.06 | 2.05 (1.15-3.66) |
| Cirrhosis | 100 (fixed) | **20.36** | 10.78-36.25 | 19.78 (12.90-30.33) |
| **Variables selected via Lasso penalty** | | | | |
| Phosphatidylc cluster | 75 | **3.40** | 1.74-15.93 | 3.07 (1.77-5.33) |
| IDL-C cluster | 89 | **0.30** | 0.11-0.63 | 0.27 (0.14-0.53) |
| XS-VLDL-P cluster | 94 | **3.12** | 1.62-5.29 | 3.44 (2.25-5.25) |
| Docosahexaenoic acid | 92 | **1.86** | 1.33-2.96 | 1.84 (1.32-2.55) |
| Omega-3 fatty acids | 96 | **0.56** | 0.35-0.93 | 0.59 (0.43-0.82) |
| Sphingomyelins | 87 | **0.58** | 0.32-0.86 | 0.62 (0.39-0.99) |
| VLDL-TG cluster | 86 | 0.68 | 0.32-1.34 | 0.50 (0.31-0.81) |
| Glutamine | 100 | **0.72** | 0.61-0.81 | 0.73 (0.67-0.79) |
| Triglycerides in large HDL | 94 | 1.36 | 0.90-3.12 | 1.13 (0.83-1.53) |
| Citrate | 100 | **1.35** | 1.17-1.54 | 1.34 (1.17-1.53) |
| Tyrosine | 100 | **1.29** | 1.12-1.47 | 1.29 (1.12-1.48) |
| Degree of unsaturation | 95 | 0.80 | 0.57-1.10 | 0.74 (0.55-0.99) |
| LDL-FC cluster | 83 | **1.25** | 1.01-2.25 | 1.24 (0.92-1.68) |
| Glucose | 100 | **1.24** | 1.11-1.38 | 1.24 (1.14-1.35) |
| Omega-6 cluster | 67 | 0.81 | 0.31-1.26 | 0.90 (0.55-1.48) |
| LDL-TG cluster | 65 | 0.81 | 0.48-1.19 | 0.70 (0.49-1.02) |
| Creatinine | 99 | **0.83** | 0.71-0.96 | 0.84 (0.73-0.96) |
| Average diameter for LDL particles | 89 | 0.89 | 0.75-1.01 | 0.87 (0.75-1.00) |
| Fischer ratio | 93 | 0.95 | 0.81-1.08 | 0.96 (0.85-1.08) |
| Alanine | 78 | 1.03 | 0.88-1.22 | 1.02 (0.89-1.18) |
| Acetate | 83 | 1.00 | 0.88-1.21 | |
| HDL-C cluster | 63 | 1.00 | 0.54-2.27 | |
| S-HDL-L cluster | 57 | 1.00 | 0.78-2.95 | |
| ApoB cluster | 47 | 1.00 | 0.55-2.82 | |
| Free cholesterol in small HDL | 47 | 1.00 | 0.28-1.10 | |
| HDL-TG cluster | 40 | 1.00 | 0.22-0.79 | |

**Bootstrap Selection (%):** Proportion of 1,000 bootstrap samples in which the variable was selected using Lasso-penalized Cox regression ($\lambda = \lambda_{min}$ selected via 5-fold cross-validation). For unpenalized variables, selection frequency is reported as 100 (fixed) to indicate forced inclusion.
**Bootstrap Median HR:** Median hazard ratio across all bootstraps, obtained from an unpenalized Cox regression refit using only the variables selected in each bootstrap. For penalized variables, HR = 1 was imputed when the variable was not selected. For unpenalized variables, the HR was calculated in every sample. **Bold** indicates median HRs with consistent directionality and bootstrap support (selected HR 95% percentile range excludes 1), not formal statistical significance.
**HR 95% Range (Selected Bootstraps):** 2.5th and 97.5th percentiles of HRs from unpenalized Cox refits in bootstrap samples where the variable was selected. For unpenalized variables, this reflects all bootstraps.
**Final HR (95% CI):** Variables with a bootstrap median hazard ratio ≠ 1 were included in the final model, which was refit using unpenalized Cox regression. Confidence intervals are shown for completeness but may underestimate uncertainty for penalized variables, as they do not account for variability due to selection.

**Abbreviations:** CI, confidence interval; HR, Hazard Ratio; MASH, Metabolic dysfunction-associated steatohepatitis; MASLD, Metabolic dysfunction-associated steatotic liver disease;

**Supplementary Table S16.** Variables Robustly Associated with Intrahepatic Cholangiocarcinoma in a Cox Model with Lasso Selection and Bootstrap Resampling.

| Variable | Bootstrap Selection (%) | Bootstrap Median HR | HR 95% Range (Selected Bootstraps) | Final HR (95% CI) |
|---|---|---|---|---|
| **Unpenalized adjustment variables** | | | | |
| Female sex | 100 (fixed) | 0.84 | 0.59-1.07 | 0.85 (0.66-1.08) |
| Age at blood sampling | 100 (fixed) | **1.35** | 1.08-1.70 | 1.35 (1.06-1.73) |
| Fasting time | 100 (fixed) | 1.08 | 0.95-1.20 | 1.09 (0.98-1.22) |
| **Unpenalized time-dependent covariates** | | | | |
| Cirrhosis | 100 (fixed) | **5.48** | 2.28-10.35 | 5.57 (2.81-11.05) |
| PSC | 100 (fixed) | **18.49** | 8.66-31.97 | 18.55 (10.19-33.78) |
| **Variables selected via Lasso penalty** | | | | |
| Tyrosine | 81 | **1.16** | 1.06-1.33 | 1.20 (1.07-1.34) |
| Triglycerides in large HDL | 72 | 1.16 | 0.93-1.43 | 1.22 (1.07-1.38) |
| Glutamine | 74 | **0.87** | 0.77-0.94 | 0.85 (0.76-0.94) |
| ApoB cluster | 59 | 0.93 | 0.71-1.04 | 0.89 (0.80-0.99) |
| Degree of unsaturation | 69 | 0.94 | 0.78-1.03 | 0.90 (0.81-1.01) |
| Albumin | 60 | **0.97** | 0.81-0.98 | 0.97 (0.93-1.01) |
| Lactate | 45 | 1.00 | 1.06-1.28 | |
| Average diameter for LDL particles | 44 | 1.00 | 0.82-0.98 | |
| Glucose | 38 | 1.00 | 1.03-1.19 | |
| Fischer ratio | 37 | 1.00 | 0.78-0.97 | |
| HDL-TG cluster | 35 | 1.00 | 0.88-1.46 | |
| LDL-FC cluster | 27 | 1.00 | 0.96-1.28 | |
| Acetate | 22 | 1.00 | 0.89-1.21 | |
| Glycine | 17 | 1.00 | 0.92-1.57 | |
| S-HDL-C cluster | 15 | 1.00 | 0.72-1.01 | |
| Creatinine | 15 | 1.00 | 0.77-1.08 | |
| LDL-TG cluster | 12 | 1.00 | 1.02-1.56 | |
| Citrate | 12 | 1.00 | 0.84-1.16 | |
| Alanine | 12 | 1.00 | 0.78-1.13 | |
| HDL-C cluster | 11 | 1.00 | 1.00-1.49 | |
| S-HDL-L cluster | 10 | 1.00 | 0.92-1.44 | |
| Docosahexaenoic acid | 9 | 1.00 | 0.86-1.10 | |
| Valine cluster | 7 | 1.00 | 1.04-1.38 | |
| XL-HDL-C cluster | 7 | 1.00 | 0.85-1.39 | |
| IDL-C cluster | 6 | 1.00 | 0.61-1.06 | |
| Omega-3 fatty acids | 5 | 1.00 | 0.76-1.20 | |
| Omega-6 cluster | 4 | 1.00 | 0.54-1.01 | |
| Sphingomyelins | 2 | 1.00 | 1.09-1.97 | |
| Phosphatidylc cluster | 2 | 1.00 | 0.94-1.83 | |
| XS-VLDL-P cluster | 1 | 1.00 | 0.58-1.49 | |

**Bootstrap Selection (%):** Proportion of 1,000 bootstrap samples in which the variable was selected using Lasso-penalized Cox regression ($\lambda = \lambda_{min}$ selected via 5-fold cross-validation). For unpenalized variables, selection frequency is reported as 100 (fixed) to indicate forced inclusion.
**Bootstrap Median HR:** Median hazard ratio across all bootstraps, obtained from an unpenalized Cox regression refit using only the variables selected in each bootstrap. For penalized variables, HR = 1 was imputed when the variable was not selected. For unpenalized variables, the HR was calculated in every sample. **Bold** indicates median HRs with consistent directionality and bootstrap support (selected HR 95% percentile range excludes 1), not formal statistical significance.
**HR 95% Range (Selected Bootstraps):** 2.5th and 97.5th percentiles of HRs from unpenalized Cox refits in bootstrap samples where the variable was selected. For unpenalized variables, this reflects all bootstraps.

**Final HR (95% CI):** Variables with a bootstrap median hazard ratio ≠ 1 were included in the final model, which was refit using unpenalized Cox regression. Confidence intervals are shown for completeness but may underestimate uncertainty for penalized variables, as they do not account for variability due to selection.
**Abbreviations:** CI, confidence interval; HR, Hazard Ratio; PSC, primary sclerosing cholangitis;

**Supplementary Table S17.** Variables Robustly Associated with Extrahepatic Cholangiocarcinoma in a Cox Model with Lasso Selection and Bootstrap Resampling.

| Variable | Bootstrap Selection (%) | Bootstrap Median HR | HR 95% Range (Selected Bootstraps) | Final HR (95% CI) |
|---|---|---|---|---|
| **Unpenalized adjustment variables** | | | | |
| Female sex | 100 (fixed) | 0.65 | 0.38-1.01 | 0.73 (0.48-1.09) |
| Age at blood sampling | 100 (fixed) | 1.35 | 0.96-1.90 | 1.36 (0.91-2.01) |
| Fasting time | 100 (fixed) | 1.08 | 0.83-1.28 | 1.08 (0.90-1.29) |
| **Unpenalized time-dependent covariates** | | | | |
| PSC | 100 (fixed) | **45.82** | 16.50-88.87 | 44.70 (20.46-97.67) |
| **Variables selected via Lasso penalty** | | | | |
| IDL-C cluster | 53 | **0.92** | 0.61-0.93 | 0.89 (0.73-1.07) |
| Docosahexaenoic acid | 51 | **0.96** | 0.78-0.94 | 0.88 (0.75-1.04) |
| HDL-TG cluster | 26 | 1.00 | 1.08-1.54 | |
| Fischer ratio | 25 | 1.00 | 0.68-1.08 | |
| Glycine | 16 | 1.00 | 1.15-2.40 | |
| Alanine | 15 | 1.00 | 0.73-1.27 | |
| Valine cluster | 13 | 1.00 | 0.71-1.30 | |
| XL-HDL-C cluster | 12 | 1.00 | 0.87-1.72 | |
| S-HDL-C cluster | 12 | 1.00 | 0.90-1.63 | |
| Sphingomyelins | 8 | 1.00 | 0.58-1.71 | |

**Bootstrap Selection (%):** Proportion of 1,000 bootstrap samples in which the variable was selected using Lasso-penalized Cox regression ($\lambda = \lambda_{min}$ selected via 5-fold cross-validation). For unpenalized variables, selection frequency is reported as 100 (fixed) to indicate forced inclusion.
**Bootstrap Median HR:** Median hazard ratio across all bootstraps, obtained from an unpenalized Cox regression refit using only the variables selected in each bootstrap. For penalized variables, HR = 1 was imputed when the variable was not selected. For unpenalized variables, the HR was calculated in every sample. **Bold** indicates median HRs with consistent directionality and bootstrap support (selected HR 95% percentile range excludes 1), not formal statistical significance.
**HR 95% Range (Selected Bootstraps):** 2.5th and 97.5th percentiles of HRs from unpenalized Cox refits in bootstrap samples where the variable was selected. For unpenalized variables, this reflects all bootstraps.
**Final HR (95% CI):** Variables with a bootstrap median hazard ratio ≠ 1 were included in the final model, which was refit using unpenalized Cox regression. Confidence intervals are shown for completeness but may underestimate uncertainty for penalized variables, as they do not account for variability due to selection.
**Abbreviations:** CI, confidence interval; HR, Hazard Ratio; PSC, primary sclerosing cholangitis;

**Supplementary Table S18.** Variables Robustly Associated with Ampulla of Vater Cancer in a Cox Model with Lasso Selection and Bootstrap Resampling.

| Variable | Bootstrap Selection (%) | Bootstrap Median HR | HR 95% Range (Selected Bootstraps) | Final HR (95% CI) |
|---|---|---|---|---|
| **Unpenalized adjustment variables** | | | | |
| Female sex | 100 (fixed) | 0.76 | 0.46-1.27 | 0.75 (0.47-1.19) |
| Age at blood sampling | 100 (fixed) | 1.14 | 0.72-1.76 | 1.14 (0.70-1.85) |
| Fasting time | 100 (fixed) | 0.94 | 0.70-1.14 | 0.95 (0.72-1.23) |
| **Unpenalized time-dependent covariates** | | | | |
| Gallstones | 100 (fixed) | 1.48 | 0.36-3.77 | 1.48 (0.61-3.62) |
| PSC | 100 (fixed) | **49.31** | 11.95-135.6 | 49.97 (19.08-130.9) |
| **Variables selected via Lasso penalty** | | | | |
| VLDL-TG cluster | 24 | 1.00 | 1.06-1.77 | |
| S-HDL-C cluster | 10 | 1.00 | 0.91-1.81 | |
| Sphingomyelins | 9 | 1.00 | 0.44-0.92 | |
| S-HDL-L cluster | 7 | 1.00 | 0.82-2.21 | |
| IDL-C cluster | 4 | 1.00 | 0.77-2.25 | |
| HDL-TG cluster | 2 | 1.00 | 0.44-1.32 | |
| Free cholesterol in small HDL | 1 | 1.00 | 0.29-1.39 | |

**Bootstrap Selection (%):** Proportion of 1,000 bootstrap samples in which the variable was selected using Lasso-penalized Cox regression ($\lambda = \lambda_{min}$ selected via 5-fold cross-validation). For unpenalized variables, selection frequency is reported as 100 (fixed) to indicate forced inclusion.
**Bootstrap Median HR:** Median hazard ratio across all bootstraps, obtained from an unpenalized Cox regression refit using only the variables selected in each bootstrap. For penalized variables, HR = 1 was imputed when the variable was not selected. For unpenalized variables, the HR was calculated in every sample. **Bold** indicates median HRs with consistent directionality and bootstrap support (selected HR 95% percentile range excludes 1), not formal statistical significance.
**HR 95% Range (Selected Bootstraps):** 2.5th and 97.5th percentiles of HRs from unpenalized Cox refits in bootstrap samples where the variable was selected. For unpenalized variables, this reflects all bootstraps.
**Final HR (95% CI):** Variables with a bootstrap median hazard ratio ≠ 1 were included in the final model, which was refit using unpenalized Cox regression. Confidence intervals are shown for completeness but may underestimate uncertainty for penalized variables, as they do not account for variability due to selection.
**Abbreviations:** CI, confidence interval; HR, Hazard Ratio; PSC, primary sclerosing cholangitis;
