## Supplementary Figures for "Risk Factor–Based Metabolomic Profiling Reveals Plasma Biomarkers of Hepatobiliary Cancer"

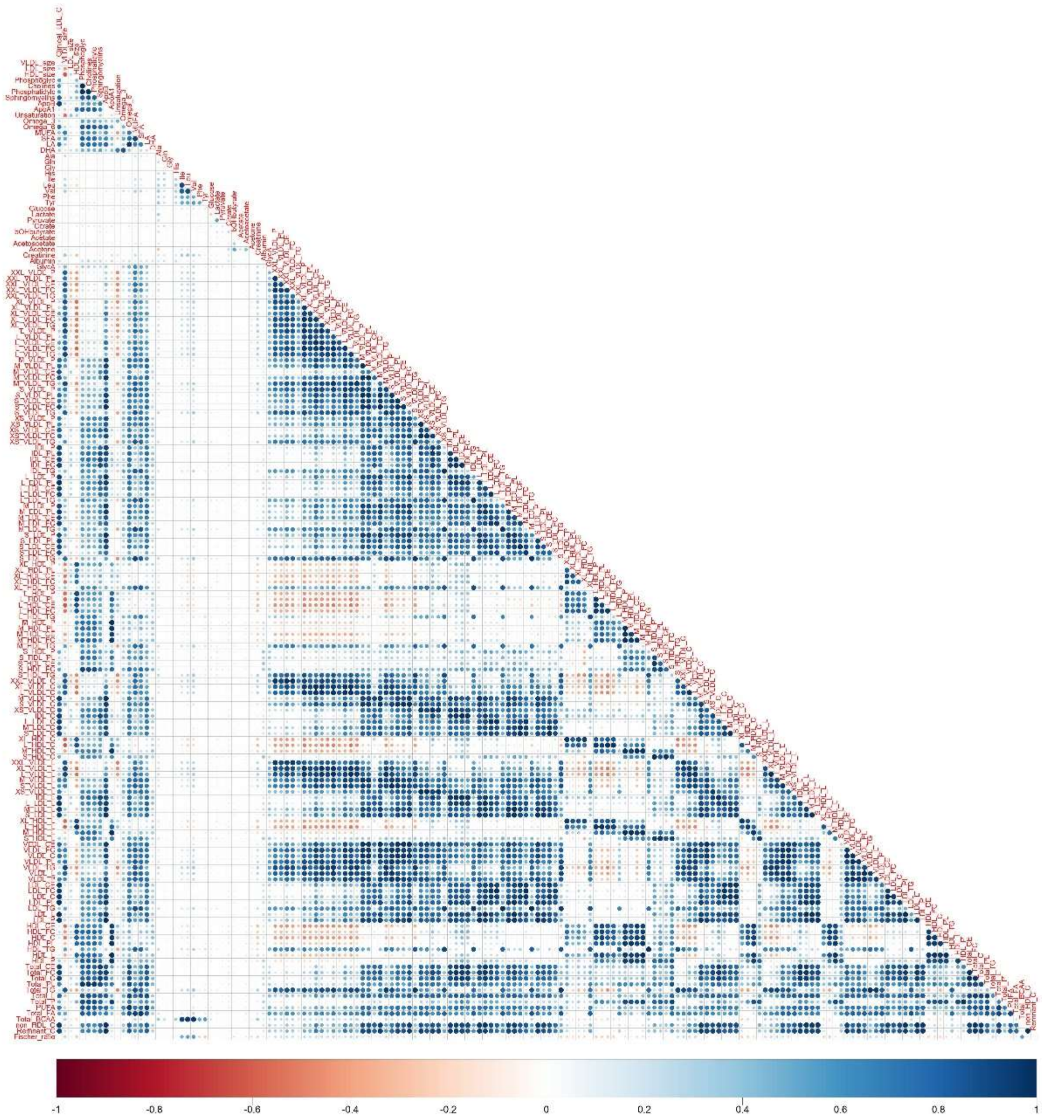

**Figure S1.** Pearson correlation between the 168 circulating metabolites and the Fisher's ratio computed in the 273,190 UK Biobank participants with metabolomics data.

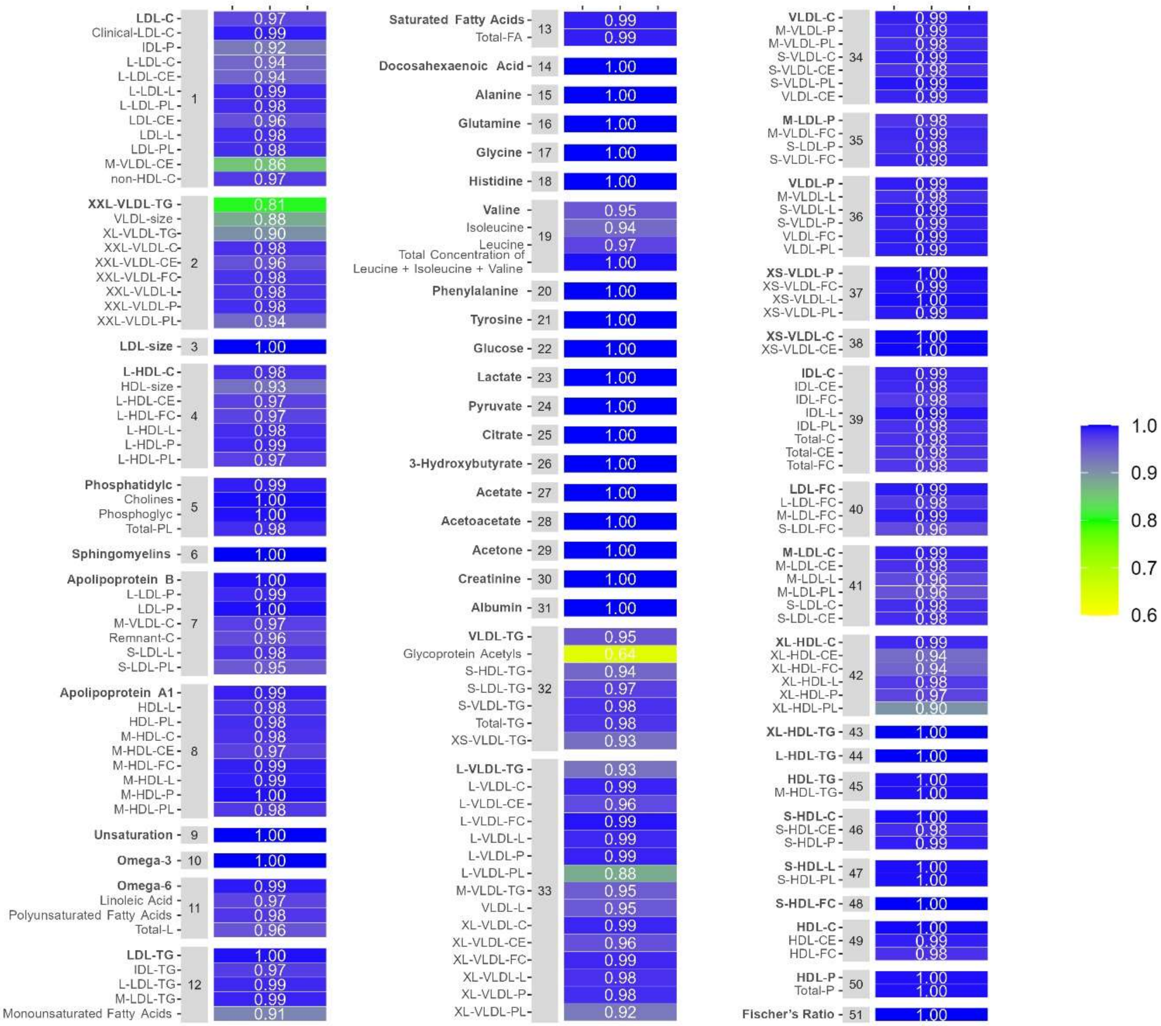

**Figure S2.** Description of the 51 metabolomic features considered included in the main analysis. The 51 metabolomic features investigated consist of 26 metabolites as surrogates of highly correlated metabolite-clusters and 25 single metabolites. For each cluster, the bolded name denotes the selected as the cluster surrogate, representing the first principal component of the metabolites within that cluster. Heat maps display the correlation between each cluster surrogate and the metabolites in that cluster.

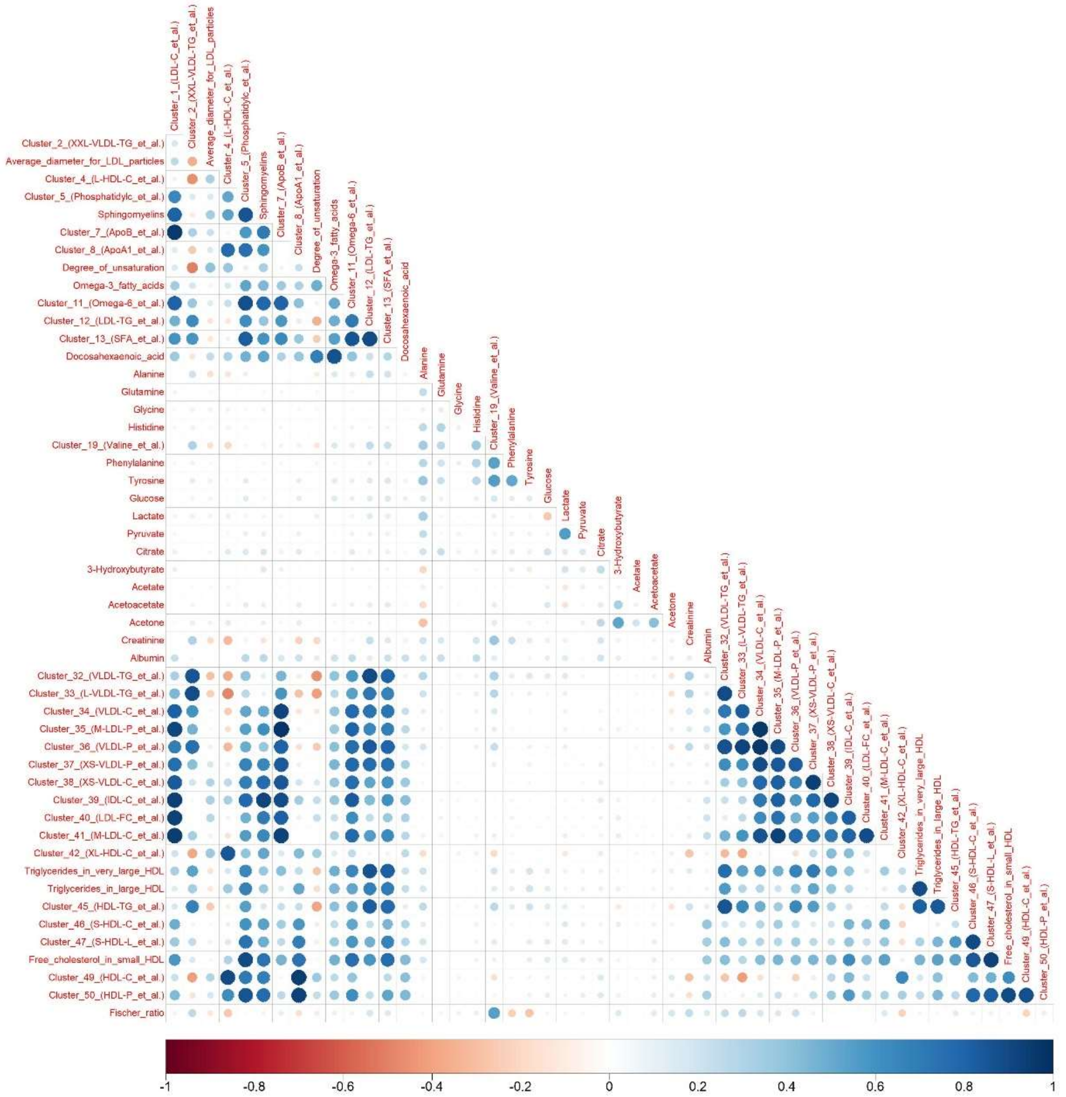


**Figure S3.** Pearson correlation between the 51 metabolite clusters computed in the 273,190 UK Biobank participants with metabolomics data. The 51 clusters correspond to 25 isolated metabolites and 26 representatives of clusters of strongly correlated metabolites, obtained by hierarchical clustering of the 168 circulating metabolites and the Fischer's ratio among the UK Biobank participants.

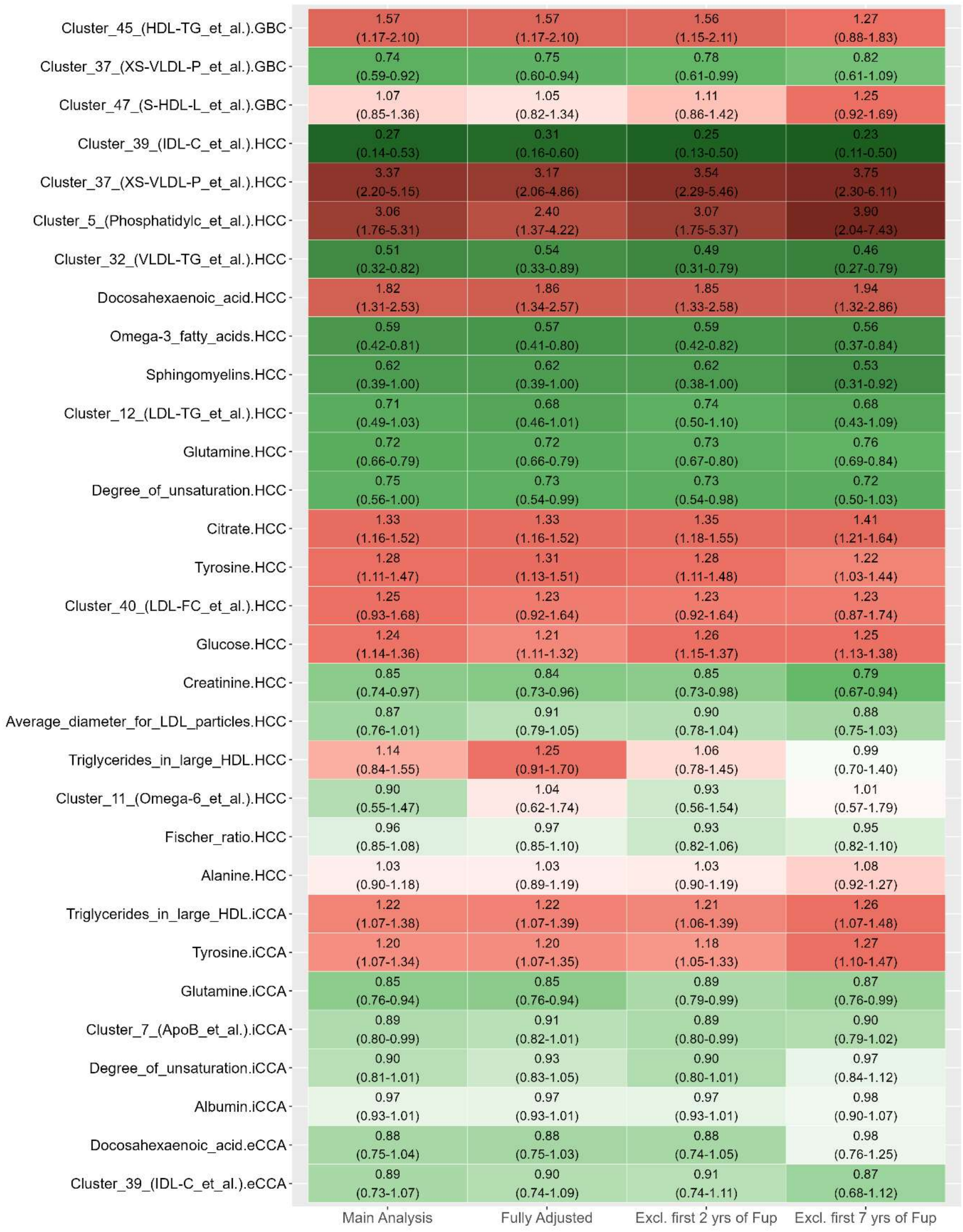


**Figure S4.** Sensitivity analyses of the mutually adjusted hazard ratios for the cancer type-specific associations identified with bootstrapping and lasso penalized Cox regression analyses in phase two. The heatmap shows the hazard ratio estimates and 95% confidence intervals for all robust metabolite-cancer association identified in phase two (bootstrap median hazard ratio $\neq$ 1). Multivariable Cox proportional hazard models were used: The main analyses model (*first column*) was adjusted for age at blood collection, sex, fasting time, and respective time-dependent risk factors (GBC: gallstones, HCC: MASLD, MASH and cirrhosis, iCCA: PSC and cirrhosis, eCCA: PSC). The fully adjusted model (*second column*) was adjusted for variables in the main analyses model plus body mass index, education level, educational level, number of years of smoking and alcohol consumption; From the main analyses model participant's age at blood sampling was shifted forward by two (*third column*) or seven years (*fourth column*); Point estimates and confidence intervals have to be interpreted with caution since they are the result of post-selection inference. eCCA, extrahepatic cholangiocarcinoma; GBC, gallbladder cancer; iCCA, intrahepatic cholangiocarcinoma; HCC, hepatocellular carcinoma.

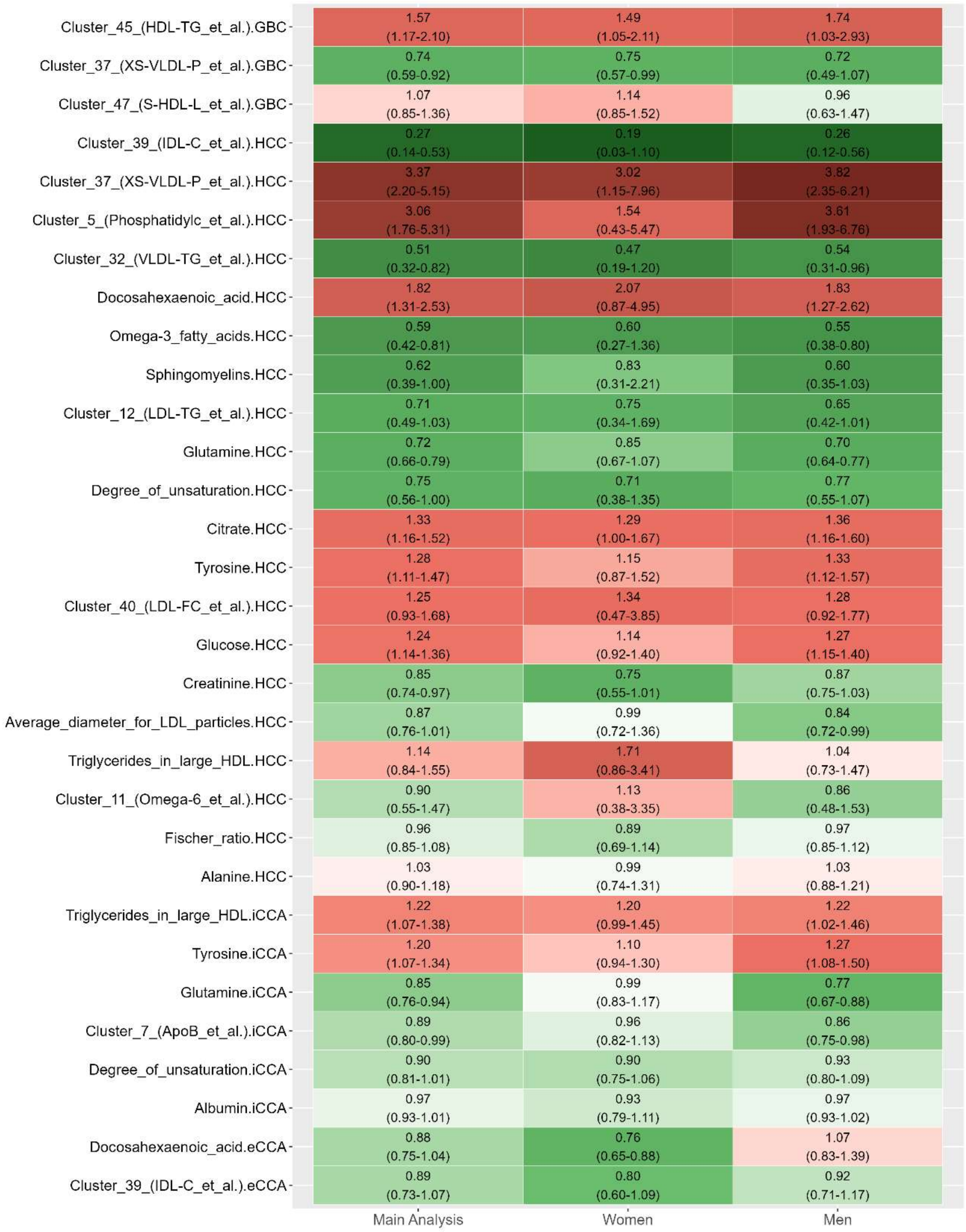


**Figure S5.** Sex-stratified analyses of the mutually adjusted hazard ratios for the cancer type-specific associations identified with bootstrapping and lasso penalized Cox regression analyses in phase two. The heatmap shows the hazard ratio estimates and 95% confidence intervals for all robust metabolite-cancer association identified in phase two (bootstrap median hazard ratio ≠ 1). Multivariable Cox proportional hazard models were used: The main analyses model (*first column*) was adjusted for age at blood collection, sex, fasting time, and menopause and use of exogenous hormones for women. The analyses stratified for women (*second column*) and (men) was adjusted for variables in the main analyses model except for gender; Point estimates and confidence intervals have to be interpreted with caution since they are the result of post-selection inference. eCCA, extrahepatic cholangiocarcinoma; GBC, gallbladder cancer; iCCA, intrahepatic cholangiocarcinoma; HCC, hepatocellular carcinoma.

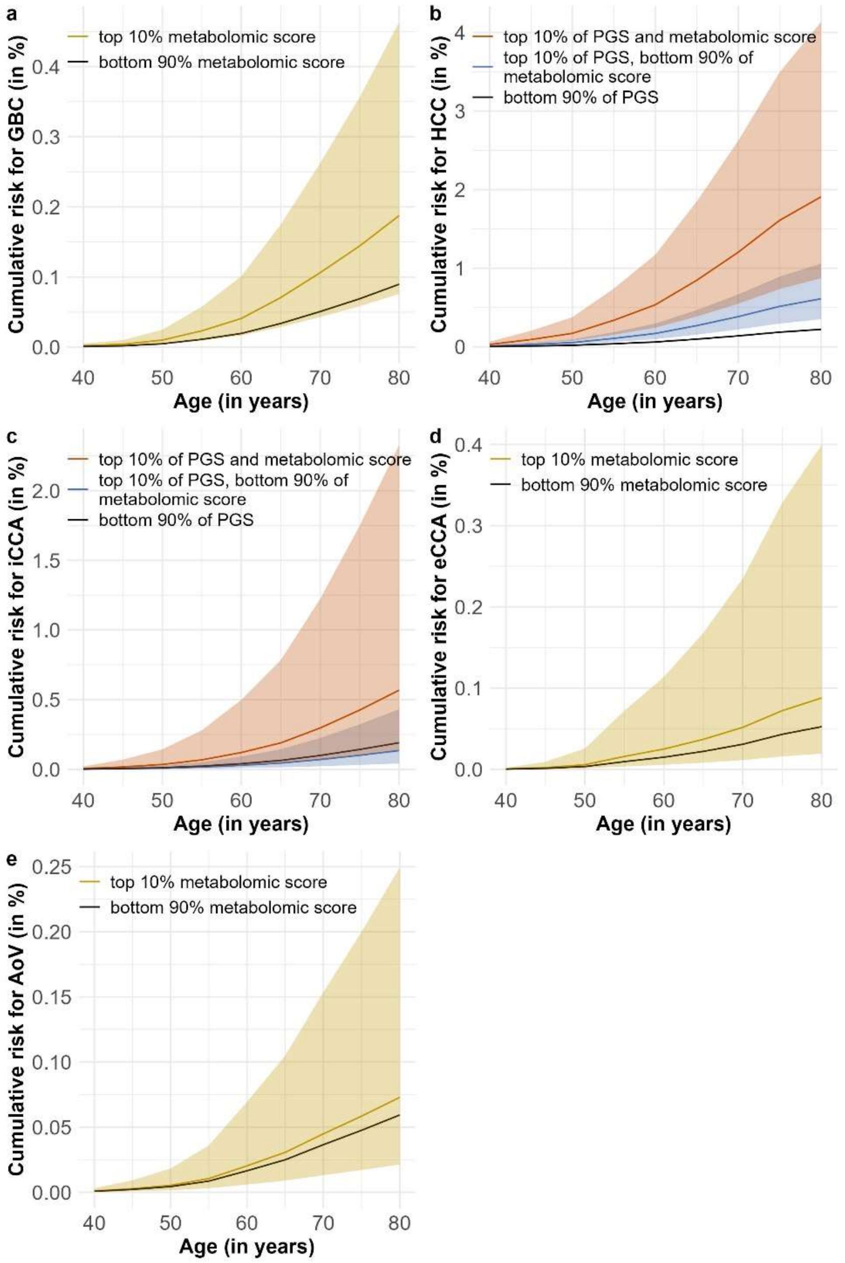


**Figure S6.** Absolute cumulative risk of hepatobiliary cancers by metabolomic and genetic risk groups among participants with diagnosed clinical risk factors in UK Biobank batch 3. Panels show age-specific absolute cumulative risk (in %) for five hepatobiliary cancer types among participants with diagnosed clinical risk factors. Risks are based on metabolomic scores trained in the discovery study and, for hepatocellular carcinoma (HCC) and intrahepatic cholangiocarcinoma (iCCA), additionally on cirrhosis-related polygenic risk scores (PGS). Curves were derived from multivariable Cox models fitted in this subgroup, with absolute risks estimated using a competing-risks framework incorporating UK population incidence and mortality rates.

**a, d, e** For gallbladder cancer (GBC), extrahepatic cholangiocarcinoma (eCCA), and ampulla of Vater cancer (AoV), individuals in the top 10% of the metabolomic score are compared with the remaining 90%.

**b, c** For HCC and iCCA, three mutually exclusive groups are shown:

(1) top-decile PGS and top-decile metabolomic score,

(2) top-decile PGS with metabolomic score in the lower 90%,

(3) lower 90% of PGS (reference).

Shaded areas represent 95% confidence intervals.

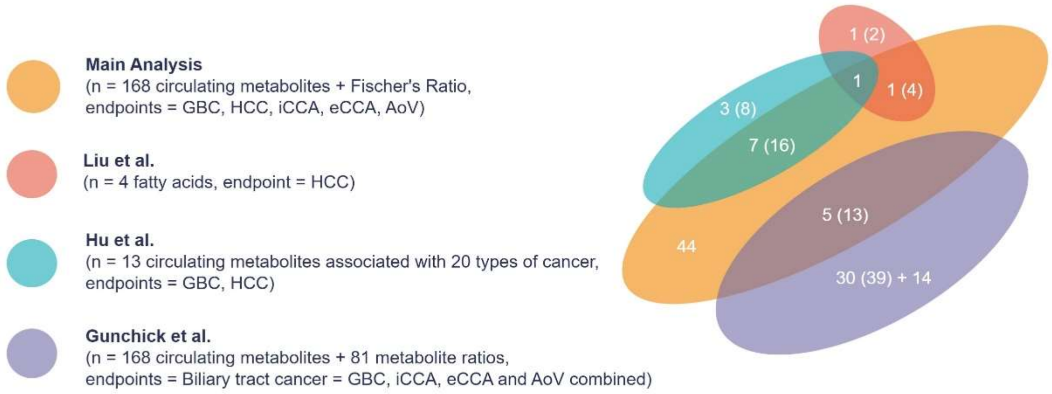


**Figure S7.** Overlap between metabolite-cancer associations identified in our analyses and those reported in previous studies. We included all robust metabolite-cancer associations identified in phase two of our analysis. Liu *et al.* investigated associations between fatty acids and hepatocellular carcinoma (HCC); data were extracted from Supplementary Table S7, Model 3[1]. Hu *et al.* initially identified 13 metabolites associated with overall cancer risk and subsequently evaluated these across 20 cancer types, including HCC and gallbladder cancer (GBC); associations were obtained from Supplementary Table S4[2]. Gunchick *et al.* analyzed biliary tract cancer (BTC) as a composite outcome, including extrahepatic cholangiocarcinoma (eCCA), intrahepatic cholangiocarcinoma (iCCA), GBC, and ampulla of Vater (AoV) cancers, and assessed associations with all circulating metabolites and metabolite ratios; data were extracted from Supplementary Table S4, Model 1[3]. Because our statistical analyses were conducted on 51 cluster representatives for circulating metabolites and the Fischer's ratio, we reported overlapping metabolite-cancer associations in two ways: (1) by individual metabolites, and (2) by all metabolites included within each representative cluster (numbers in brackets). For example, Hu *et al.* analyzed omega-6 fatty acids, which in our study were grouped with three other metabolites in Cluster 11. The reported omega-6–HCC association was the only one uniquely shared between our findings and theirs; therefore, the number of overlapping individual metabolites between our analysis and Hu *et al.* was 1 (or 4 when accounting for all metabolites within the corresponding cluster). To enable comparison with results reported by Gunchick *et al.*, we combined our cancer subtype-specific associations for GBC, iCCA, and eCCA into a single composite outcome. In addition, since we did not analyze metabolite ratios directly, we calculated Pearson correlations between each of our 51 metabolite clusters and the 81 metabolite ratios. For ratios showing strong correlations ($r > 0.6$), we assigned them to the cluster with the highest correlation. The number of non-overlapping metabolite–BTC associations reported by Gunchick *et al.* was as follows: 30 individual metabolite-BTC associations corresponding to ratios strongly correlated with our clusters (39 when considering all metabolites within those correlated clusters), and 14 additional individual metabolite–BTC associations not strongly correlated with any of our clusters.

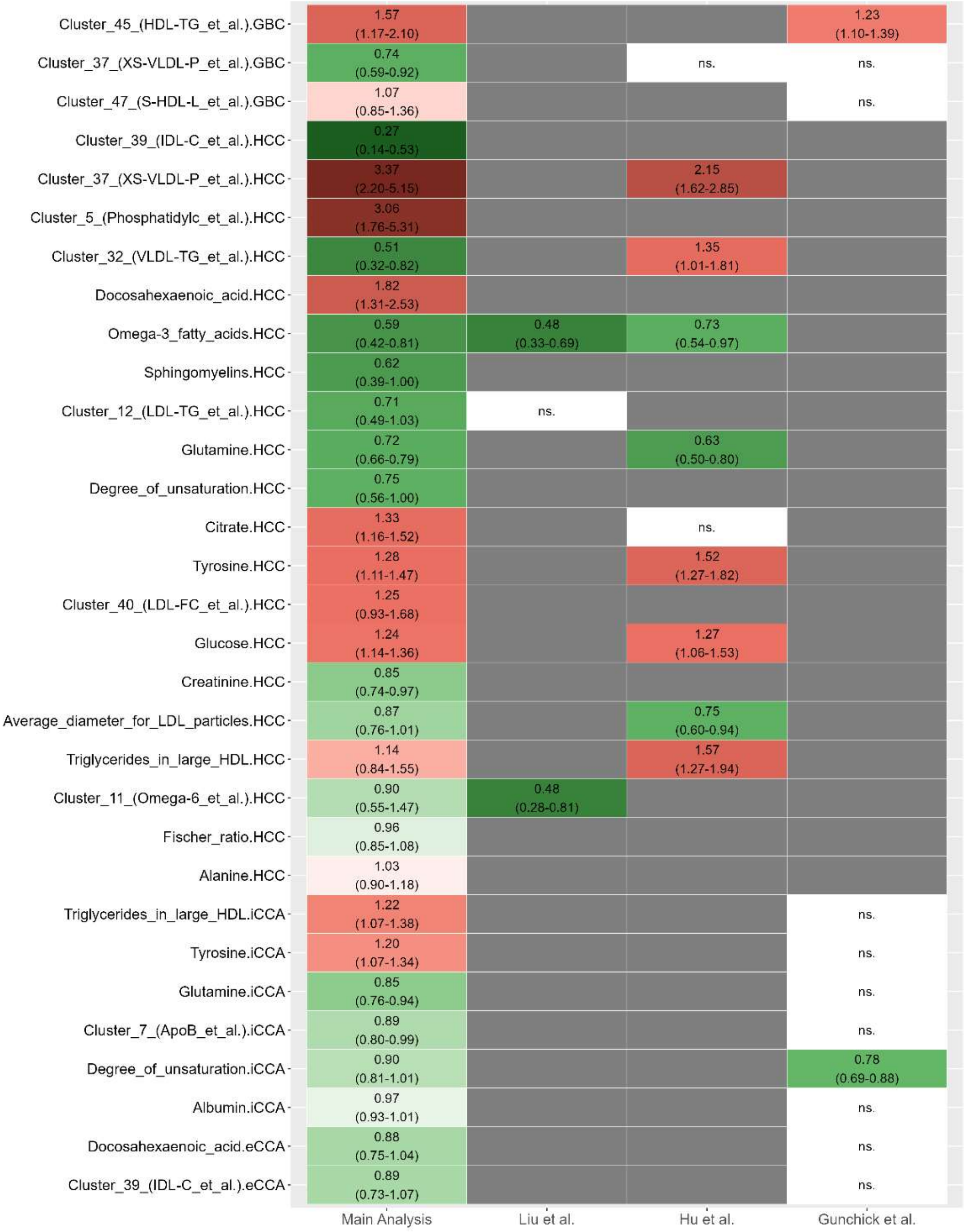


**Figure S8.** Comparison between overlapping metabolite-cancer associations identified in our analyses and those reported in previous studies. The heatmap shows the hazard ratio estimates and 95% confidence intervals for all robust metabolite-cancer association identified in phase two (bootstrap median hazard ratio $\neq 1$). Multivariable Cox proportional hazard models were used: The main analyses model (*first column*) was adjusted for age at blood collection, sex, fasting time, and menopause and use of exogenous hormones for women. The model by Liu *et al.* (*second column*) was adjusted for age, sex, ethnicity, body mass index, waist circumference, Townsend deprivation index, education level, household income, self-reported smoking status, self-reported frequency of alcohol intake, physical activity, diet quality score, baseline hypertension, baseline diabetes, and baseline dyslipidemia, total cholesterol level, triglycerides level, total fatty acids level, serum ALT level, serum AST level, and blood platelet count (Suppl Table S7, Model 3 in Liu *et al.*)[1]; The model by Hu *et al.* (*third column*) was adjusted by age, sex, family history of cancer, education level, average total household income, smoking status, drinking status, body mass index, days of moderate to vigorous physical activity, fruit and vegetable intake, red meat intake, processed meat intake, fish intake, total cholesterol, low-density lipoprotein cholesterol and high-density lipoprotein cholesterol (Suppl Table S4 in Hu *et al.*)[2]; The model by Gunchick *et al.* (*fourth column*) was adjusted by age, sex, education, income, fasting time and statin use (Suppl Table S2, Model 1 in Gunchick *et al.*)[3].
